# Application of wastewater and environmental surveillance for pathogenic agents during the 2024 National Football League (NFL) Draft in Detroit, Michigan (USA)

**DOI:** 10.64898/2026.03.20.26348829

**Authors:** Ryland Corchis-Scott, Ethan Harrop, Qiudi Geng, Mackenzie Beach, John Norton, Mehdi Aloosh, Thomas Reid, Rajesh Seth, Christopher G. Weisener, R. Michael McKay

**Author notes:** Corresponding author: Ryland Corchis-Scott, Great Lakes Institute for Environmental Research, University of Windsor, 401 Sunset Ave., Windsor, ON, Canada N9B 3P4.

## Abstract

Mass gatherings pose a concern for public health because they are associated with dense crowds, increased social interaction, and travel, all of which can facilitate the rapid transmission of infectious diseases. Wastewater and environmental surveillance (WES) were used for pathogen monitoring during the 2024 NFL Annual Player Selection Meeting (the ‘Draft’) in Detroit, MI, an event that drew an estimated 775,000 attendees. Wastewater and environmental samples were queried for respiratory viruses and clinically relevant ARGs. WES did not detect an increase in the concentration of monitored respiratory viruses (SARS-CoV-2, IAV, IBV, and RSV) associated with the 2024 NFL Draft. In contrast, WES detected a transient increase in carbapenemase targets in wastewater, primarily driven by a fourfold increase in *bla_OXA-48_*. Resistome structure in wastewater was dominated by sampling site characteristics rather than changes associated with the event. The Draft weekend coincided with rainfall-driven combined sewer overflow (CSO), potentially allowing the dissemination of antimicrobial resistance genes (ARGs) to the environment. In surface waters receiving wastewater effluent, an increase in detection frequency and normalized concentrations for multiple ARGs were observed following the Draft. WES provided an overview of pathogen prevalence before, during, and after a large-scale gathering, showing how it can warn of emerging health risks in near real time.

## Introduction

Analysis of wastewater can provide information that accurately reflects infectious diseases dynamics^1–3^ by integrating signals from symptomatic, subclinical, and asymptomatic individuals^4–7^. Wastewater and environmental surveillance (WES) is not a new discipline, but it has experienced a renaissance following its successful implementation during the COVID-19 pandemic^8–10^. Increased interest in WES has been driven by the numerous advantages it offers, including correlation with clinical metrics^11–17^, cost-effectiveness^18–21^, independence from test-seeking behaviour or availability of testing infrastructure^22^, scalability from building to city levels^14,23–25^, and timeliness with near real-time data generation^12,26,27^.

An emerging application of WES is infectious diseases surveillance during mass gatherings^28^. Mass gatherings are typically large, planned events that concentrate individuals from diverse geographies into dense settings with high mobility, conditions that facilitate the transmission of infectious diseases^29^. Mass gatherings pose a challenge to epidemiological disease tracking due to travel and increased likelihood of rapid transmission. Disease surveillance before, during and after events allows for rapid response to widespread transmission following disease detection^28,30^. WES offers an effective means by which populations may be monitored surrounding large-scale gatherings such as sporting events^31–33^.

Prior applications of WES to mass gatherings have targeted SARS-CoV-2 to monitor transmission of COVID-19 and include the 2020 Tokyo Olympics^34^, the Two Oceans Marathon in South Africa^35^, and the 2022 FIFA World Cup in Qatar^36^. Recent festival-based monitoring in New Orleans, LA, expanded the application of WES to mass gatherings, where it was implemented to target Mpox transmission^37^. Beyond respiratory viruses, surveillance of antimicrobial resistance (AMR) via antimicrobial resistance genes (ARGs) is gaining traction, given public health relevance and the potential for dissemination facilitated by mass gatherings^38^. Antimicrobial-resistant organisms (AROs) are also targets of expanded WES^39,40^ with surveillance focused on the presence of specific ARGs of public health significance^41^. Monitoring has been established as a critical step in addressing the proliferation of AMR^42^ with efforts having been made to test for the presence and level of ARGs at wastewater treatment facilities^39,43,44^. Exacerbating the threat of AMR are rainfall-driven combined sewer overflows (CSOs) and bypass events that can introduce partially treated effluent into receiving waters, increasing environmental ARG loads.

WES may be implemented through a OneHealth framework that considers the movement of organisms and resistance determinants between humans, infrastructure, and natural systems^45^. Wastewater is a critical connection between these systems and thus surveillance implemented here captures both human population dynamics and pathways for dissemination to the environment. Mass gatherings increase population density and can strain sanitation infrastructure. WES implemented during these events is opportune for advancing knowledge of disease trajectories in human population and ARG dissemination in the environment.

The 2024 NFL Draft was held in Detroit, MI from April 25-27, 2024, with events concentrated in an area with an approximate radius of 1 km near the riverfront in downtown Detroit. This event drew approximately 775,000 individuals to the city of Detroit^46,47^ of which an estimated 30% travelled more than 160 km to attend the event. Visitors to Detroit included people from over 20 different nations and all 50 states, with some individuals coming from as far afield as Australia^48^. Almost all of Detroit’s 45,140 hotel rooms were occupied for the Draft weekend, with spillover absorbed by neighbouring Windsor, ON, a region separated from Detroit by a river and high-traffic international land border.

Here, we used the 2024 NFL Draft as a natural experiment to explore WES utility for mass gatherings across two domains: (1) respiratory pathogens in wastewater; and (2) public health relevant ARGs, with emphasis on carbapenemases, in wastewater and the Detroit River, an environmental reservoir for wastewater treatment plant (WWTP) effluent. We also compared multiplex RT-qPCR with a nanofluidic OpenArray® platform to assess method agreement across matrices. We hypothesized that respiratory signals would not increase immediately given the primarily outdoor nature, late-season timing of the event, and its brevity (in comparison to the incubation of respiratory viruses). We also expected that ARGs could show transient changes, and that heavy rainfall coincident with the Draft could elevate environmental ARG detections due to CSO and/or bypass activity.

## Methods

### Site information

Sampling sites included five wastewater treatment facilities and a surface water transect of the Detroit River with six sample collection sites (Figure 1). The Draft weekend coincided with a wet weather period in the Windsor-Detroit region, with approximately 40 mm of rain falling over a 48 h period (Sunday-Tuesday)^49^. This influx of precipitation contributed to CSO events where partially treated WWTP effluent and diluted raw sewage was discharged into the Detroit River or its tributaries from both sides of the Canda-USA border^50^. Wastewater treatment facilities were identified as potentially impacted by the NFL Draft or as control sites based on proximity (Table 1). Additional site information can be found in S1.

**Figure 1.**
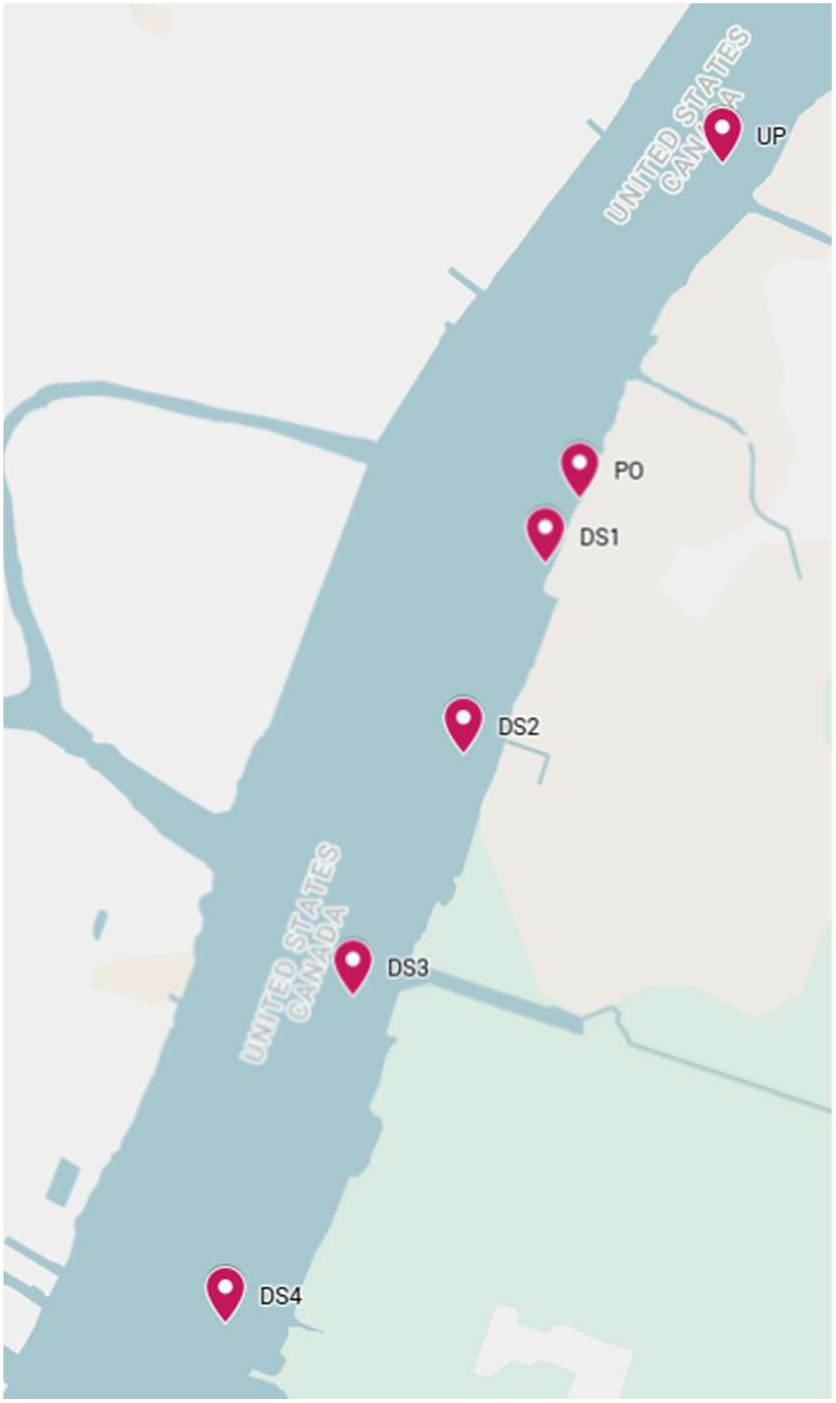
Locations of sampling sites making up the Detroit River transect. Samples were collected on 2024-04-25, prior to the 2024 NFL Draft, and on 2024-04-29, after the event.

**Table 1.** Site characteristics and sample collection modalities.

| Facility Name | Site ID | Location | Site type | Population served | Sample type | Sampling frequency | Role |
| --- | --- | --- | --- | --- | --- | --- | --- |
| Water Resource Recovery Facility (WRRF) | NIEA | Detroit, MI, USA | Interceptor feeding wastewater treatment plant | 1482000 | 1L 24h composite | 1×weekly. Daily sampling during Draft | Impacted site |
|  | OAK | Detroit, MI, USA | Interceptor feeding wastewater treatment plant | 840600 | 1L 24h composite | 1×weekly. Daily sampling during Draft | Impacted site |
|  | JEFF | Detroit, MI, USA | Interceptor feeding wastewater treatment plant | 492000 | 1L 24h composite | 1×weekly. Daily sampling during Draft | Impacted site |
| Anonymous Windsor-Essex Wastewater Treatment Plant | WEWTP | Windsor-Essex, ON, Canada | Wastewater treatment plant | 180000 | 1L 24h composite | 5×weekly. Daily sampling during Draft | Impacted site |
| Leamington Pollution Control Centre | LPCC | Leamington, ON, Canada | Wastewater treatment plant | 18000 | 1L 24h composite | 3×weekly | Control site |
| Chatham Water Pollution Control Plant | CHWW | Chatham, ON, Canada | Wastewater treatment plant | 48000 | 0.5L 24h composite | 3×weekly | Control site |
| Atlantic Avenue Water Pollution Control Plant | TB | Thunder Bay, ON, Canada | Wastewater treatment plant | 97500 | 0.5L 24h composite | 3×weekly | Control site |
| Detroit River Transect | UP | Upstream of Detroit River outfall | Surface water | NA | 2L Grab | Event based | Env sample |
|  | PO | Detroit River outfall | Surface water | NA | 2L Grab | Event based | Env sample |
|  | DS1 | Downstream of Detroit River outfall | Surface water | NA | 2L Grab | Event based | Env sample |
|  | DS2 | Downstream of Detroit River outfall | Surface water | NA | 2L Grab | Event based | Env sample |
|  | DS3 | Downstream of Detroit River outfall | Surface water | NA | 2L Grab | Event based | Env sample |
|  | DS4 | Downstream of Detroit River outfall | Surface water | NA | 2L Grab | Event based | Env sample |

### Sample collection

Untreated wastewater influent was collected from each of the wastewater treatment facilities included in the study at different weekly frequencies (Table 1) with daily sampling between April 24-29 from each of the three interceptors (NIEA, OAK, and JEFF) of the Great Lakes Water Authority Water Resource Recovery Facility (WRRF) as well as the WWTP plant (WEWTP) in Windsor, ON. All samples were transported directly to the laboratory in coolers on ice and processed upon arrival. Surface water grab samples (2 L) were collected in duplicate from each site in the Detroit River transect (Figure 1).

Environmental samples were collected on April 25, prior to the 2024 NFL Draft, and on April 29 following the end of the he event.

### Sample processing

Influent samples collected from WWTPs were concentrated by filtering between 60-120 mL through 0.22 μm Sterivex PES cartridge filters (MilliporeSigma, Burlington, MA). River water samples were likewise concentrated by passing 500-800 mL of water through cartridge filters. Immediately following filtration, the filters were sealed and flash-frozen through immersion in liquid nitrogen, with filters stored at -80°C before nucleic acid extraction. Total nucleic acids were extracted from filters using the AllPrep PowerViral DNA/RNA kit (Qiagen, Germantown, MD) with the addition of 5% (v/v) 2-mercaptoethanol to the lysis buffer. Nucleic acids were eluted in 50 μL of RNAse-free water. Samples were not treated with DNase upon extraction. Extracted nucleic acids were stored in LoBind tubes at -80°C.

### RT-qPCR assays for quantification of respiratory viruses and carbapenemase genes

Respiratory viruses, select carbapenemase genes and Pepper Mild Mottle Virus (PMMoV) were quantified using established RT-qPCR assays. Severe Acute Respiratory Syndrome Coronavirus 2 (SARS-CoV-2), influenza A virus (IAV), influenza B virus (IBV), and respiratory syncytial virus (RSV) were the respiratory viruses measured. PMMoV, a human fecal indicator^51^ and established wastewater surveillance normalizing agent^52^, was used to normalize the wastewater signal for targets measured using RT-qPCR assays. Carbapenemase genes, including *bla_KPC_*, *bla_NDM_*, *bla_OXA-48_*, *bla_VIM_*, and *bla_VIM7_* were measured using two multiplexed assays^44^. All targets were measured in at least technical triplicate. Standard curves were generated using serial dilution of synthetic RNA or DNA controls. Inhibition was assessed using an internal amplification control, and template RNA was diluted to overcome inhibition. Negative controls were included with each RT-qPCR run. Detailed RT-qPCR methods are described in S2.

### Development of the nanofluidic qPCR card using TaqMan® assays in the OpenArray ® platform

Assays for the quantification of clinically important and locally relevant ARGs and genes associated with mobile genetic elements (MGEs) were selected for inclusion on the OpenArray® card (Table 2). The OpenArray® card also included assays for 16S rRNA and crAssphage to allow normalization and act as controls. Assays were selected from the literature and optimized for TaqMan chemistry. The resulting nanofluidic RT-qPCR card was used to analyze samples in technical duplicate in 33nL reactions on a Quant Studio 12K Flex Real-Time PCR System (Thermo Fisher Scientific, Waltham, MA). A 7-point standard curve, created through a serial dilution of pooled gBlocks™ (Integrated DNA Technologies, Coralville, IA), was run on each OpenArray ® plate. Assay performance met established technical standards for RT-qPCR. Full assay design and nanofluidic qPCR methods can be found in S3.1 and S3.2.

**Table 2.** Antimicrobial resistance genes monitored, associated drug resistance, and rationale for inclusion on the Nanofluidic qRT-PCR chip

| Class of antimicrobial | Gene(s) | Rationale for testing |
| --- | --- | --- |
| Aminoglycoside | *aph(3')-Ia*,<br>*aac(6')-Ib-cr* | Often used in pediatric settings<sup>53</sup><br>Renewal in interest with rise in AMR for treatment of MDR infections<sup>54</sup>. Most common resistance mechanism for aminoglycosides is through enzymatic modification with the listed ARGs coding for the most common enzymes<sup>55</sup> |
| Carbapenems | *bla<sub>KPC</sub>*, *bla<sub>NDM</sub>*,<br>*bla<sub>OXA-48</sub>*,<br>*bla<sub>IMP</sub>*, *bla<sub>VIM</sub>* | Rapid spread and ability to infect patients in healthcare settings. Rise in prevalence with few viable treatment options. Often resistant to multiple classes of antibiotic<sup>56-58</sup>. |
| Cephalosporins/extended spectrum β-lactam (ESBL) antibiotics | *bla<sub>CMY-2</sub>*, *bla<sub>SHV</sub>*,<br>*bla<sub>TEM</sub>*, *CTX-M1*, *CTX-M9*, | Beta-lactams are highly common treatments for bacterial infections but resistance levels are high<sup>59</sup>. Some later generation cephalosporins and ESBL are considered last resort antibiotics<sup>60</sup>. Levels of cephalosporin resistance are generally rising<sup>61</sup>. Identified as likely a large threat to human health with emerging concerns regarding environmental spread<sup>62</sup>. |
| Colistin | *mcr-1* | Listed as a reserve antimicrobial agent for last resort use by the World Health Organization<sup>63</sup>. Rapid global spread<sup>42</sup>. |
| Fluoroquinolones | *gyrA83L-Esh*,<br>*QnrB1*, *qnrS* | Widely used therapeutics<sup>64</sup> with fluoroquinolone resistant infections resulting in worse patient outcomes and increase risk of mortality<sup>65</sup>. Resistance is widespread<sup>66-69</sup>. Many fluoroquinolones are on the WHO “Watch” list.<sup>63</sup> |
| Glycopeptides (vancomycin) | *vanA* | Last resort antibiotic used in the treatment of serious gram-positive bacterial infections<sup>70,71</sup>. Recent rapid increases in resistance<sup>72</sup> Vancomycin is on the WHO “Watch” list.<sup>63</sup> |
| Macrolides | *mphA* | Used widely in the treatment of common infections such as pneumonia, sinusitis, and chlamydia. Members include erythromycin, clarithromycin, azithromycin<sup>73</sup>. Associated with mobility<sup>74</sup> *mphA* is plasmid associated and mobile with evidence of transmission through horizontal gene transfer<sup>75</sup>. Commonly prescribed throughout Ontario<sup>76</sup> |
| Methicillin | *mecA* | Primary gene associated with beta-lactam resistance in Methicillin-Resistant Staphylococcus aureus (MRSA)<sup>77</sup> Thought to be mobile and has been shown confer advantages to bacterial exposed to a diversity of antimicrobial agents/chemical environments<sup>78</sup> |
| Tetracycline | *tetA*, *tetM* | Broadly administered in agriculture and human healthcare<sup>62</sup>. Evidence of co-selection with fluoroquinolone resistance genes<sup>79</sup> |
| Sulfonamides | *sul1*, *sul2* | Widely used and well established class of antimicrobial agent<sup>42</sup>. Broad spectrum antibiotics with both human and agricultural applications<sup>80</sup>. |
| Trimethoprim | *dfrA1* | Antimicrobial agent often used in the treatment of urinary tract infections. Widely prescribed in Ontario<sup>76</sup>. |
| Associated with resistance to multiple therapeutics | *cfr*, *int1* | The *cfr* gene is associated with multi-drug resistance<sup>81</sup> while the *int1* gene is a sign of the presence of mobile genetic elements often associated with AMR. Presence of *int1* alone does not indicate AMR. However, it is associated with multiple ARGs and their mobility<sup>82,83</sup> |
| Normalization/Control | 16S rRNA, CrAssphage | crAssphage is a bacteriophage found in association with human waste making it a good fecal indicator and a measure of wastewater concentration that can be used for normalization of wastewater signals<sup>84,85</sup>. A general assay for 16S rRNA was also incorporated to act as a control and for normalization. |

### Statistical methods

Data analysis was performed in R (version 4.5.1)^86^ with data manipulation and visualization relying on the *tidyverse* suite of packages^87^. Multiple different measurement techniques were employed in this investigation, leading to the creation of five distinct datasets. Statistical analyses were conducted separately for each dataset due to differences in sampling frequency, sample matrix, availability of covariate data, data distribution, and response variable. All analyses tested changes in wastewater and environmental occurrence of pathogens associated with the mass gathering of 2024 NFL Draft attendees and the co-occurring CSO event.

### Interrupted time series (ITS) analysis

To test if the 2024 NFL Draft coincided with altered concentrations of respiratory pathogens or carbapenemase genes in wastewater, we fit Generalized Linear Mixed Models (GLMMs) using the *glmmTMB* package. Independent analyses were conducted on “affected” and “control” sites for a quasi-experimental design. Model specification followed a segmented regression framework with a binary “event” variable (modelling immediate event-related changes) and a “post-time” variable (more gradual event-related changes). Models controlled for a variety of covariates (pH, temperature, flow, precipitation, etc.) and temporal autocorrelation was addressed via an autoregressive term for all targets and seasonal harmonics for respiratory virus response variables. A Tweedie distribution with a log link was used for continuous zero-inflated viral pathogen concentration data. A Gamma distribution with a log link was used for individual carbapenemase gene concentration data, while a Tweedie distribution was used for the composite measure of all assayed carbapenemase genes. To account for multiple testing, p-values were adjusted using the Benjamini-Hochberg (BH) False Discovery Rate (FDR) method. Adjustment was performed independently for respiratory viruses and carbapenemase genes and for “control” and “affected” sites. Details can be found in S4.1 and S4.2.

### Multivariate resistome characterization

The Open Array nanofluidic RT-qPCR card was used to measure a subset of samples collected from WWTPs during the month surrounding the 2024 NFL Draft. ARG concentrations were normalized to the concentration of crAssphage and subjected to fourth root transformation. Data was split into the uneven groups (“before”, “during” and “after”) relative to the timing of the 2024 NFL Draft. Characterization of the resistome was carried out with Non-Metric Multidimensional Scaling (NMDS) using Bray-Curtis dissimilarity matrices. A marginal Permutational Multivariate Analysis of Variance (PERMANOVA) was then run on the data to partition sources of variation, and a distance-based redundancy analysis was used for visualization of variance partitioning. A

Permutational Analysis of Multivariate Dispersions (PERMDISP) was used to test if observed differences arose from differences in resistome or within-group variance. A principal component analysis (PCA) was run to identify ARGs driving variability. Finally, JEFF was analyzed alone as it drains the NFL Draft location. A Kruskal-Wallis test was used to determine if concentration differences existed between the “before”, “during” and “after” groupings for the measured ARGs. The p-values were adjusted for multiple comparisons using the Benjamini-Hochberg method. Details can be found in S4.3.

### Environmental detection

To determine if the CSO events altered the occurrence of carbapenemase genes in the Detroit River, detection frequencies were calculated for each carbapenemase gene in the pre- and post- CSO transects. Fisher’s exact test was used to compare the detection frequency. Additionally, a paired Wilcoxon signed-rank test was performed to evaluate concentration changes for PMMoV normalized *bla_KPC_* between the pre- and post CSO samplings.

To determine if the CSO and mass gathering event led to either changes in abundance or detection frequency for additional ARGs, data produced by the Open Array nanofluidic RT-qPCR card were employed. Data were normalized to 16S rRNA concentrations and log-transformed before quantitative analysis. Detection frequencies were also calculated for both the pre- and post- CSO groupings. Changes to detection frequency were assessed using a GLMM with a binomial distribution and logit link function. To prevent complete separation, ARGs that were constitutively detected or undetected in either the pre-CSO or post-CSO grouping were excluded from logistic regression. The main GLMM included ARG identity, sampling group (pre/post CSO) and the interaction of the two as fixed effects. Random intercepts and slopes for groups nested within sites were also included to account for differences between sites. Post-hoc comparisons were conducted using estimated marginal means. A second GLMM was conducted with site and sample grouping as fixed effects and gene as a random intercept to directly investigate site heterogeneity. Quantitative abundance differences between pre- and post-CSO sample groupings were tested using paired Wilcoxon signed rank tests with pairings assigned by site identity and ARG. A sensitivity analysis was performed to identify the influence of potential detection saturation in samples collected at a known WWTP outlet. To account for multiple comparisons, p-values were adjusted using the Benjamini-Hochberg method to control FDR. Model diagnostics were assessed using simulated residuals in the *DHARMa* package. Details can be found in S4.4 and S4.5.

### Method comparison

OpenArray® assays were compared with established RT-qPCR assays using a subset of wastewater and environmental samples that were analyzed by both methods (n=42).

Comparison was limited to assays for *bla_KPC_*, *bla_NDM_*, and *bla _OXA-48_* as they were shared between the OpenArray® and the multiplexed RT-qPCR assays. Qualitative agreement was evaluated using Cohen’s Kappa and detection frequency comparisons. Quantitative agreement was determined through Bland-Altman analysis to calculate 95% limits of agreement (LoA). Additional quantitative measures of method comparison included Passing-Bablok regression, Deming regression, spearman correlation, Pearson correlation and calculation of concordance correlation coefficients (CCC). Details can be found in S4.6.

## Results

### Respiratory pathogen dynamics

ITS analysis showed no event-associated increase in wastewater concentrations of SARS-CoV-2, IAV, IBV, or RSV at monitored sites. IBV showed a post-event decline (β=-0.356, SE=0.143, p= 0.035), consistent with seasonal-tapering rather than event impact (Figure 2). Variability in respiratory virus concentrations in wastewater was driven by dilution and seasonality. Concentrations of SARS-CoV-2 were negatively associated with flow (β=–0.288, SE=0.115, p = 0.035) and positively associated with PMMoV concentration (β = 0.379; SE=0.065, p < 0.001), consistent with dilution. Likewise, IAV concentration in wastewater was negatively predicted by flow (β=-0.467, SE=0.160, p = 0.012), consistent with dilution. However, IAV concentrations were positively predicted by precipitation (β=0.317, SE=0.079, p<0.001). RSV concentrations were positively associated with PMMoV (β= 0.455, SE= 0. 141, p = 0.005) while IAV was a negative predictor (β= -0.318, SE= 0.128 p = 0.035). Precipitation was positively associated with IBV concentration (β=0.345, SE=0.138, p = 0.035) while flow was negatively associated (β=-0.924, SE=0.284, p = 0.005). Finally, IBV was found to be positively associated with IAV concentration (β=0.814, SE=0.244, p = 0.004).

**Figure 2.**
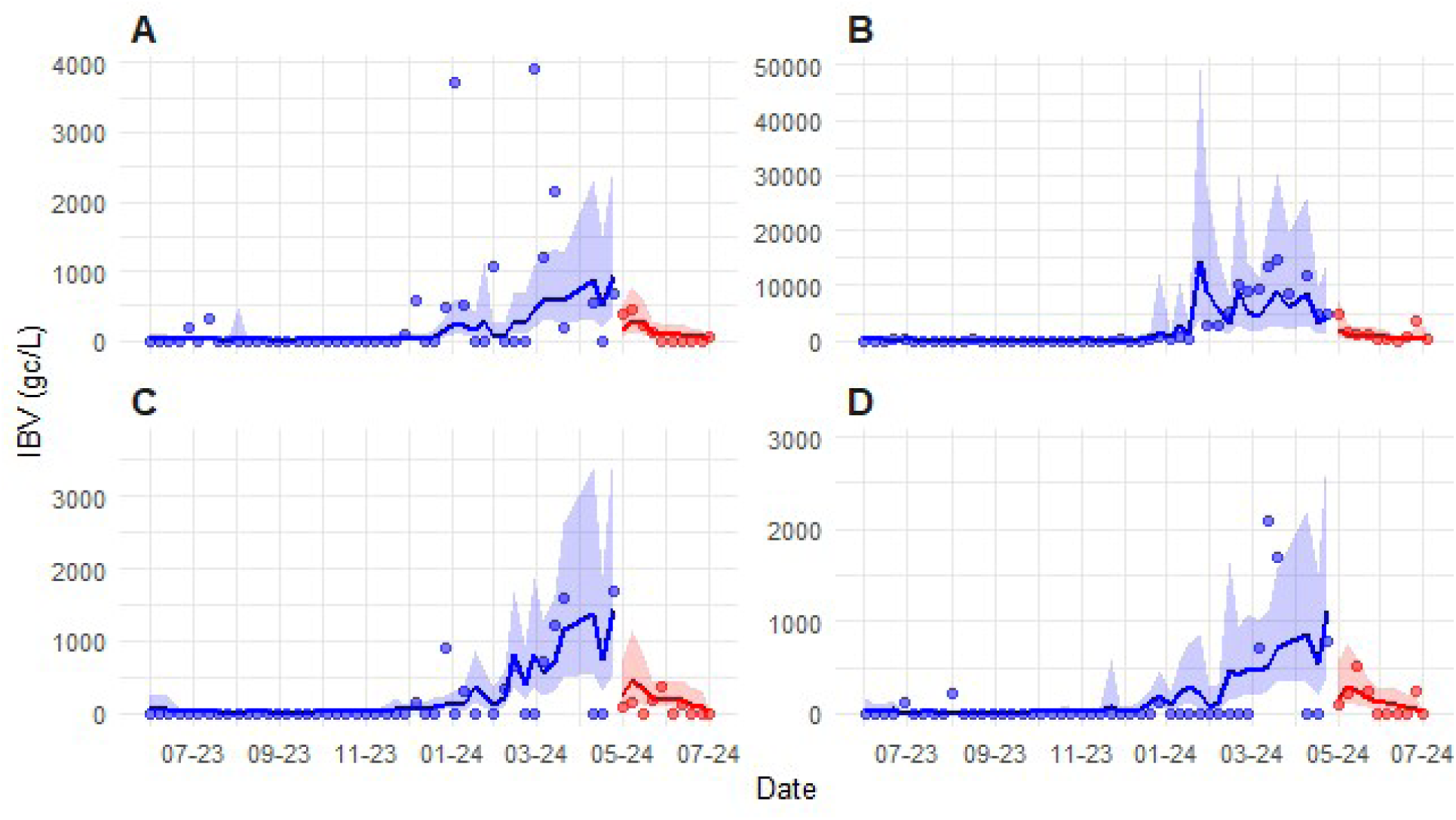
Longitudinal wastewater surveillance data for IBV at the four draft proximal sites (**A -JEFF** , **B-WEWTP, C-NIEA, D-OAK**) both before (blue), and after (red) the 2024 NFL Draft. Points represent IBV concentrations in gene copies per litre. Solid lines and shaded areas denote predicted mean concentrations and 95% confidence intervals.

Parallel analysis at control sites showed no event effects. The GLMM run for SARS-CoV-2 did not detect any stepwise increase in SARS-CoV-2 concentration or change in trend associated with the Draft. However, LRT indicated that the inclusion of “event” and “post time” parameters improved model fit (Table S10). Sensitivity analysis and visualization of the data indicate that this is due to noise in the N2 concentration at TB (Figure 3, Table 3).

**Figure 3.**
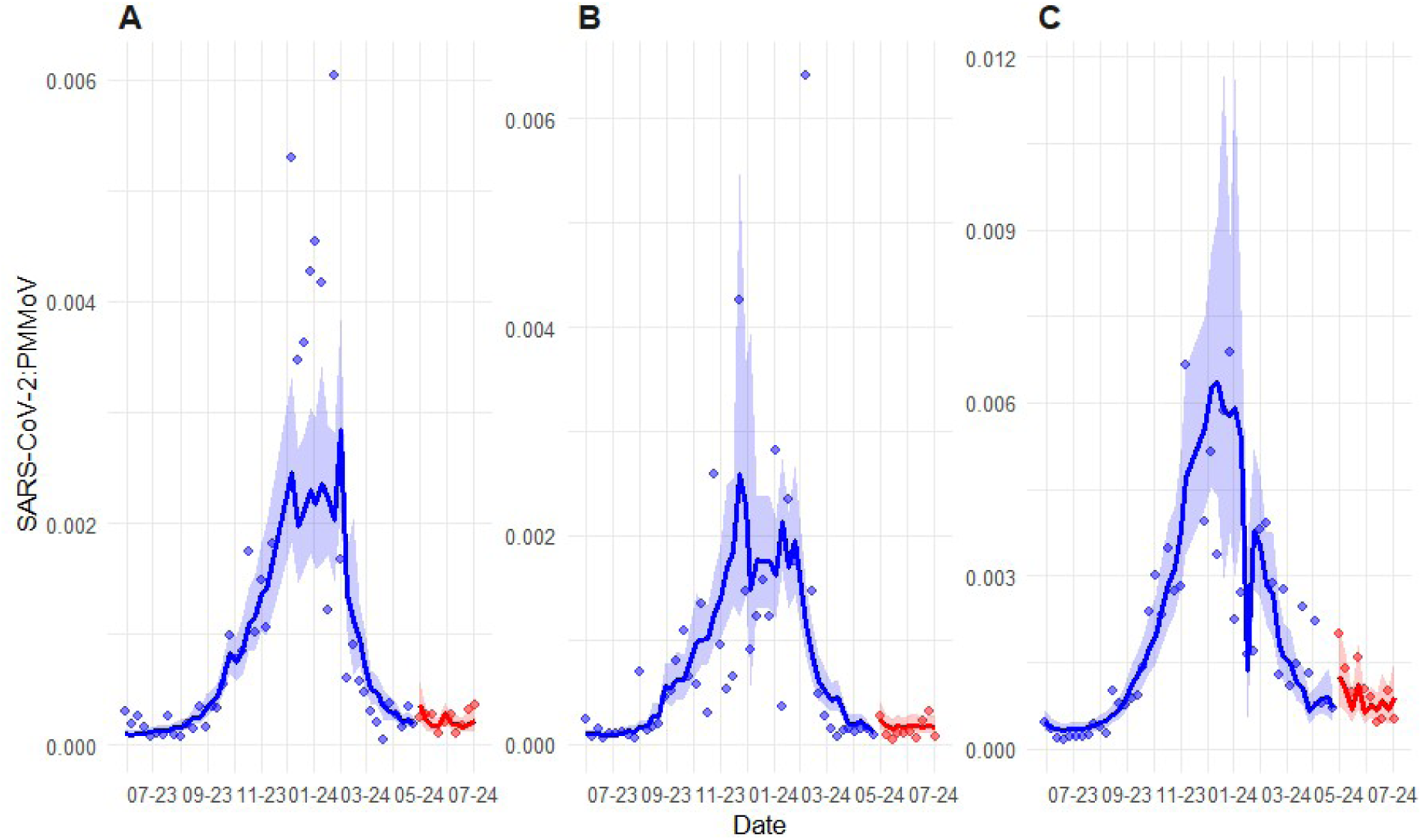
Longitudinal wastewater surveillance data for SARS-CoV-2 at the three control sites (**A -CHWW**, **B-LPCC, C-TB**) both before (blue), and after (red) the 2024 NFL Draft. Points represent mean N1 and N2 gene concentrations normalized to Pepper Mild Mottle Virus (PMMoV) concentration. Solid lines and shaded areas denote predicted mean concentrations and 95% confidence intervals, respectively, derived from a Generalized Linear Mixed Model (GLMM).

**Table 3.** Summary of Interrupted Time Series (ITS) model parameters, Likelihood Ratio Tests (LRT), and diagnostic checks for gamma GLMMS for N1 and N2 gene targets in the control site cluster.

| Metric | Statistic | Target |  |
| --- | --- | --- | --- |
|  |  | N1 | N2 |
| Model Estimates | Event | 0.333 (p=0.272) | 0.393 (p=0.179) |
|  | Post Time | 0.012 (p=0.789) | 0.025 (p=0.572) |
| Model Fit | ΔAIC | -0.5 | -4.0 |
|  | Log-Likelihood | 1122.1 | 1136.4 |
|  | Dispersion | 0.187 | 0.193 |
| LRT | Chi-Square | 4.485 | 8.066 |
|  | p-value | 0.106 | 0.018* |
| DHARMA diagnostics | KS | p=0.309 | P=0.627 |
|  | Dispersion | p=0.892 | P=0.424 |
|  | Quantiles | p=0.074 | P=0.069 |
|  | Outlier | P=0.288 | P=0.288 |

The detection of a marginal, site-specific fluctuation at a control location demonstrates sensitivity to minor background variance in the approach, reinforcing the validity of the null results observed at affected site cluster. This contrast confirms that while the modelling framework can identify localized shifts, no such signal was generated by the mass gathering event.

### Carbapenemase gene concentration in wastewater

ITS analysis indicated a statistically significant increase in the composite measure of carbapenemase gene concentrations concurrent with the 2024 NFL Draft (β=0.625, SE=0.290, p=0.031). An ∼87% increase in combined ARG burden could be observed following the Draft weekend (Figure 4).

**Figure 4.**
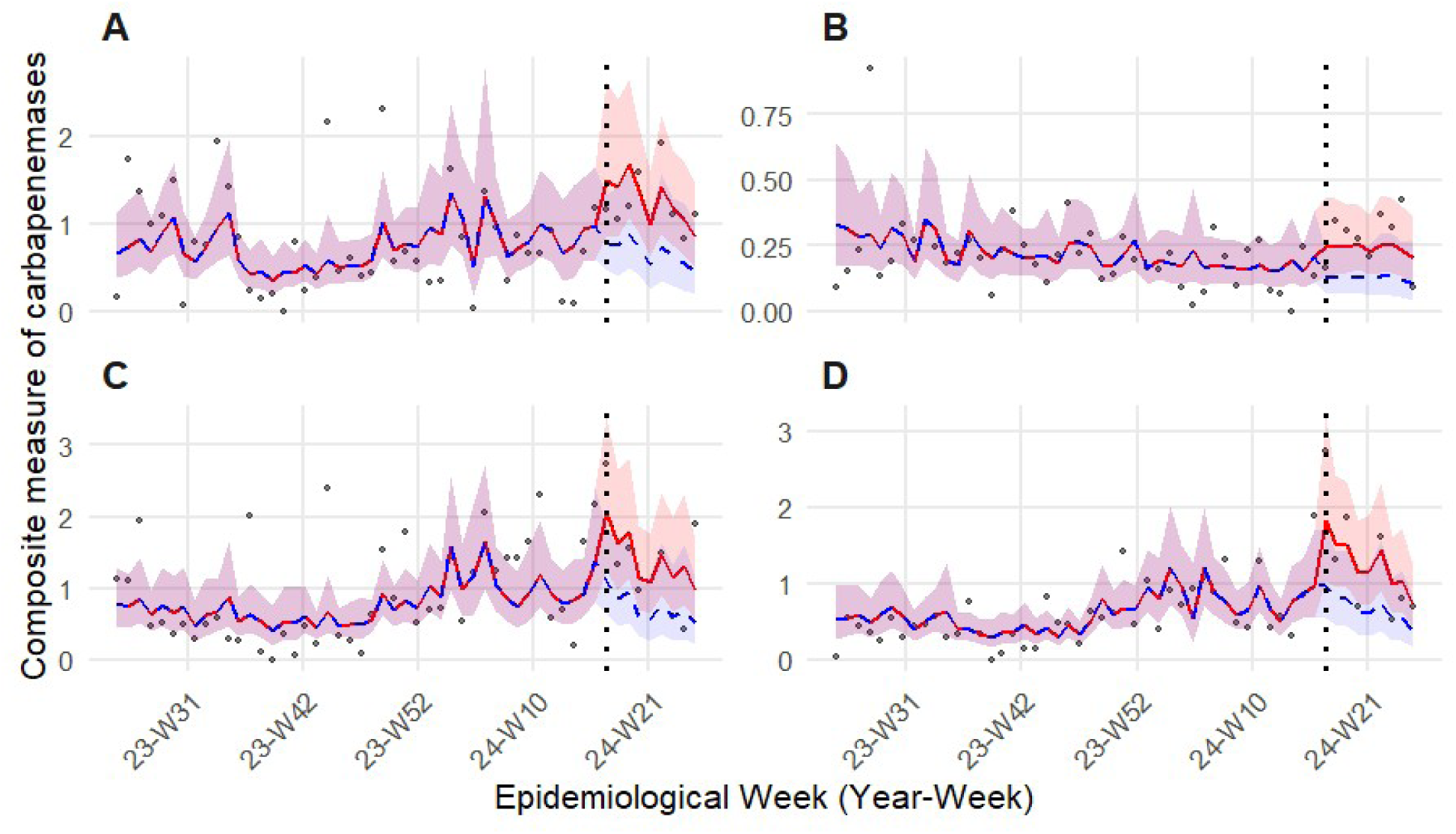
Longitudinal wastewater surveillance data for a composite measure of carbapenemase gene concentrations at the four Draft proximal sites (**A -JEFF** , **B-WEWTP, C-NIEA, D-OAK**) both before, and after the 2024 NFL Draft (vertical dotted line). Points represent a composite carbapenemase index calculated by taking the mean of z-score standardized normalized gene concentrations *for blaKPC*, *blaNDM*, *blaOXA48*, *blaVIM*, and *blaVIM7*. The redline and shading represent model predicted factual fit and 95% confidence intervals. The dashed blue line and shading represent the counter factual scenario (model predictions excluding draft related parameters).

However, the event did not lead to a change in the trend for the ARG composite index as the post-event temporal trend variable was non-significant (p = 0.96). ITS for individual carbapenemase genes revealed that the increase was primarily driven by a spike in normalized wastewater concentration for *bla_OXA-48_* (β=1.38, SE=0.374, p=0.006), which increased by ∼400% following the Draft (Figure 5). A parallel increase in *bla_KPC_* concentration (β=0.447, SE=0.212, p=0.035) did not survive FDR correction (p=0.18). ITS for *bla_NDM_, bla_VIM_*, and *bla_VIM7_* indicate that the 2024 NFL Draft did not significantly impact normalized ARG concentration despite event estimates all indicating post-event increases.

**Figure 5.**
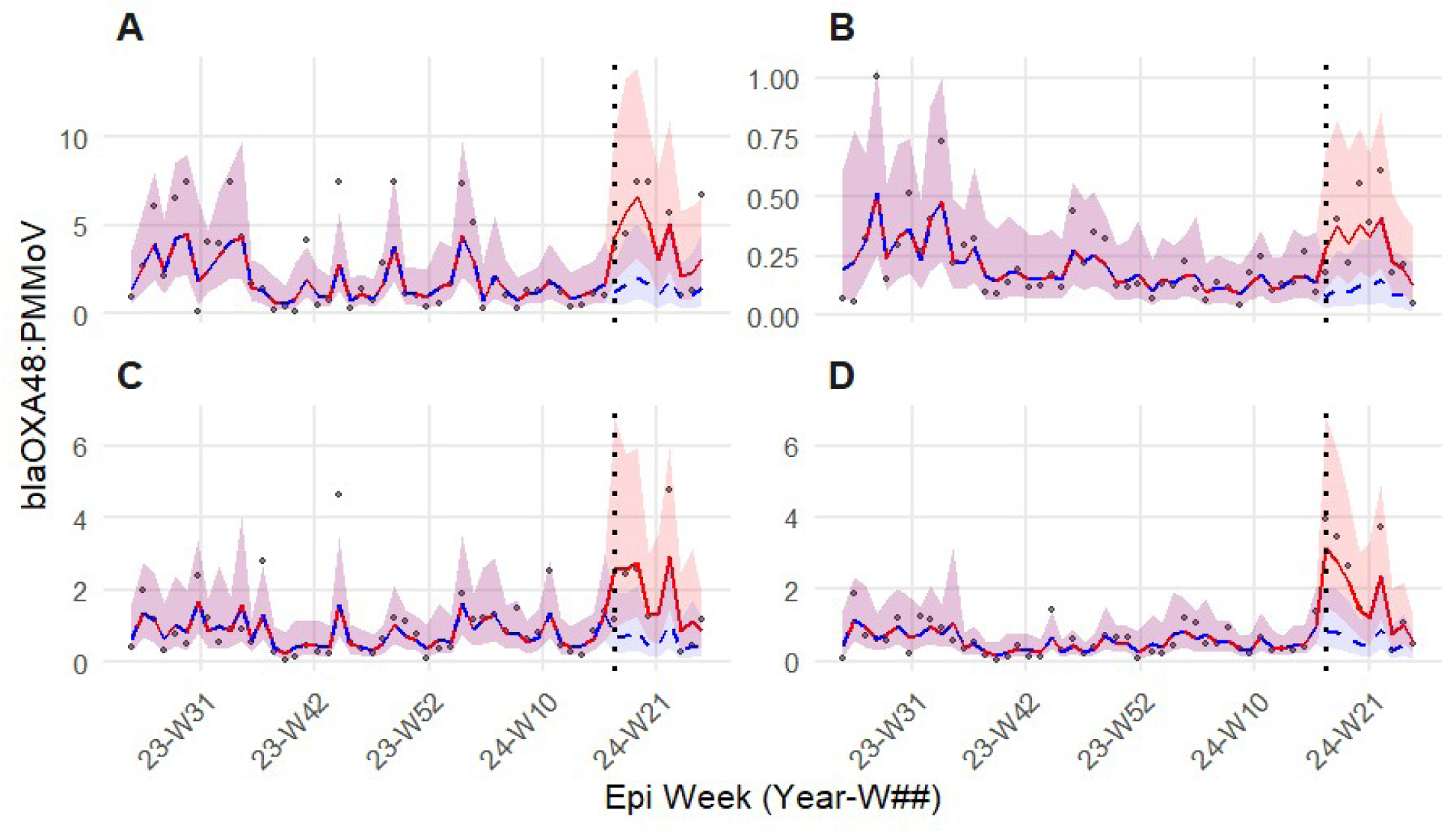
Longitudinal wastewater surveillance data for PMMoV normalized *blaOXA-48* concentrations at the four Draft proximal sites (**A -JEFF** , **B-WEWTP, C-NIEA, D-OAK**) both before, and after the 2024 NFL Draft (vertical dotted line). Points represent normalized wastewater concentrations. The redline and shading represent model predicted factual fit and 95% confidence intervals. The dashed blue line and shading represent the counter factual scenario (model predictions excluding draft related parameters).

Flow was a significant positive predictor of ARG concentration across 4 of 5 gene targets (Table S14). Higher flows were consistently associated with increased normalized ARG concentrations.

### Resistome variability

Characterization of the resistome with NMDS revealed samples collected from the same site clustered regardless of sample collection time in relation to the 2024 NFL Draft stress= 0.068; Figure 6).

**Figure 6.**
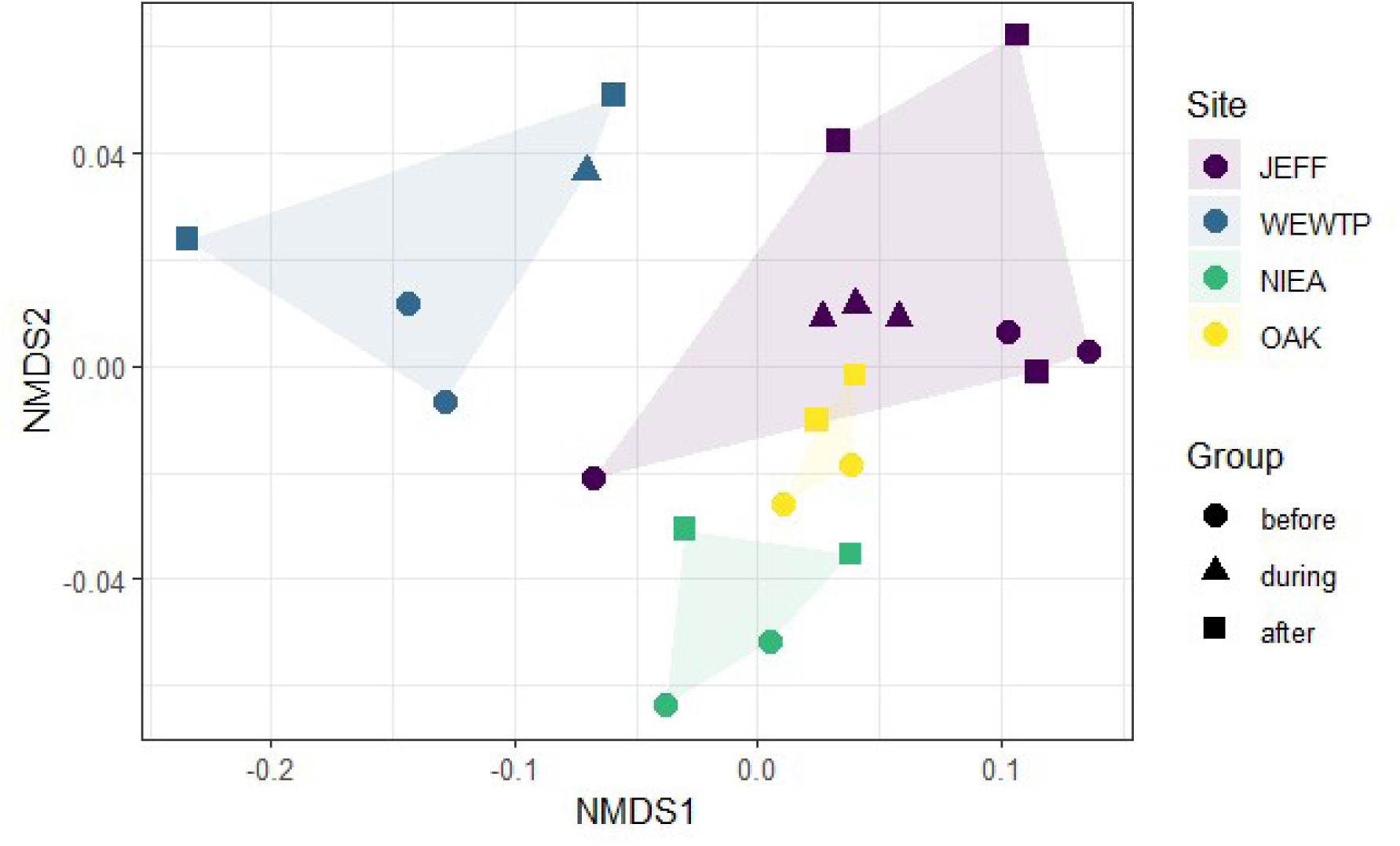
Resistome structure across collection sites and draft periods. Non-metric Multidimensional Scaling (NMDS) ordination based on Bray-Curtis dissimilarities of crAssphage normalized antimicrobial resistance gene concentrations. Shaded areas represent site-specific clusters. Points are colored by site identity and shaped by draft period (before, during, and after). The low stress value (0.068) indicates a good representation of resistome in two-dimensional space.

The marginal PERMANOVA corroborated the NMDS visualization, indicating that site identity explained 61% of variance in resistome (R^2^ = 0.611, F_3,16_= 9.30, p = 0.001) while collection period was statistically insignificant (R^2^ = 0.033, F_2,16_= 0.76, p = 0.531) (Table 4).

**Table 4.** Multivariate analysis of resistome composition. Results of the marginal Permutational Multivariate Analysis of Variance (PERMANOVA) based on Bray-Curtis dissimilarities. Site identity serves as the primary driver of community variance, while the NFL Draft period (Group) did not significantly influence the resistome structure

| Factor | Degrees of Freedom | $R^2$ | F-Statistic | p-value |
| --- | --- | --- | --- | --- |
| Site | 3 | 0.611 | 9.305 | 0.001 |
| Draft period | 2 | 0.033 | 0.760 | 0.531 |
| Residual | 16 | 0.350 | - | - |
| Total | 21 | 1 | - | - |

**Note:** Site dispersion ( $p = 0.236$ ) and Group dispersion ( $p = 0.421$ ) were non-significant, supporting the PERMANOVA findings.

The PERMDISP showed that these differences were not a result of heterogeneity of variance with no difference in dispersion between groups (F_2,19_ = 0.90, p = 0.421) or site identities (F_3,18_ = 1.55, p = 0.236). The first two principal components of the PCA accounted for 78% of the variance. The highest loadings for PC1 were *int1*, *sul1*, *aac(6′)-Ib-cr*, *tet(A)*, *sul2*, *mphA, qnrs*, *dfrA1*, *bla_OXA-48_*, and *bla_TEM_*. The top PC1 loadings had the same sign, indicating the top contributors varied together across collection sites (Figure 7). The Kruskal-Wallis test for JEFF revealed no significant difference in concentration for any ARG in the “before”, “during”, or “after” groups.

**Figure 7.**
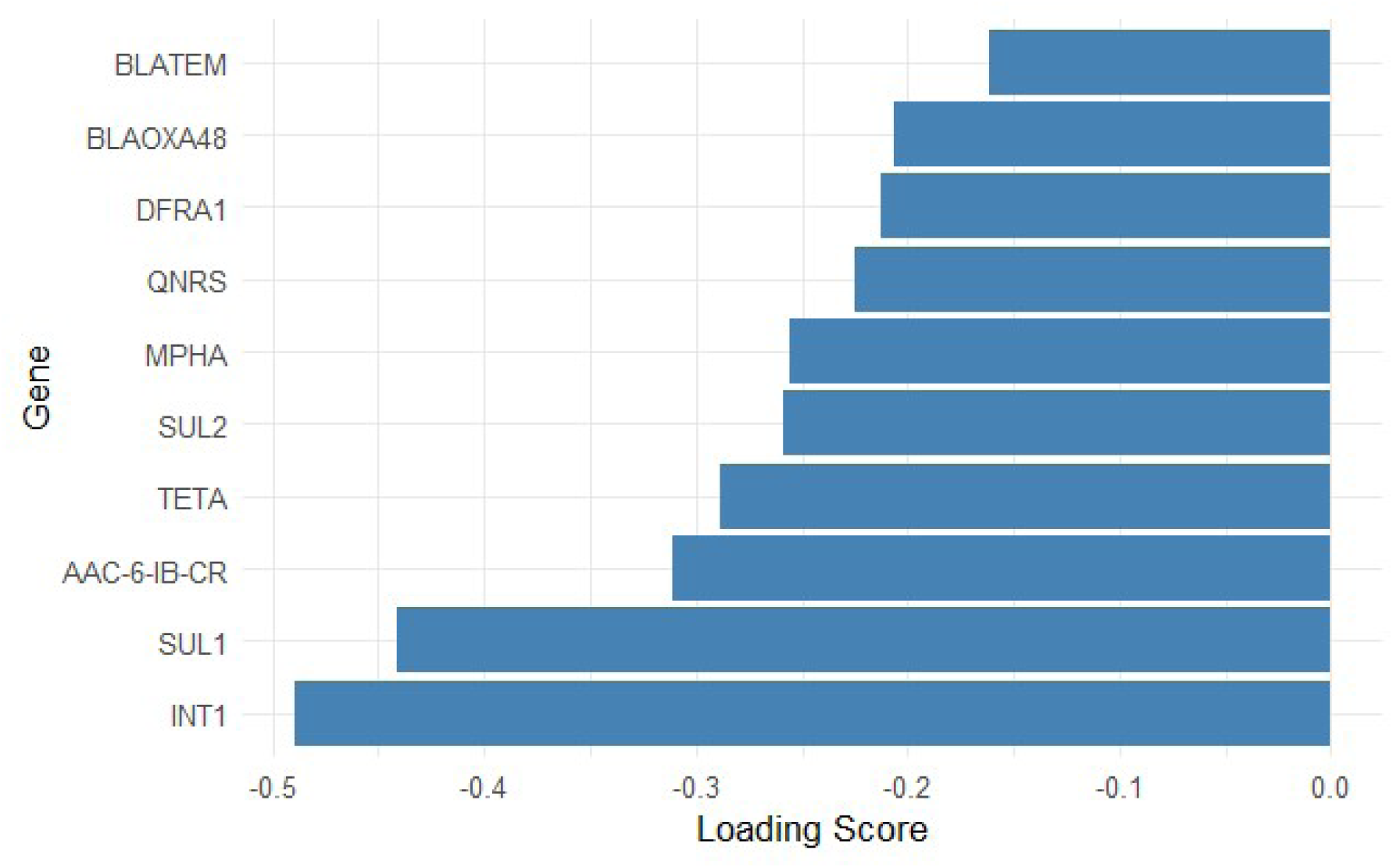
Loading scores for the top 10 antimicrobial resistance genes (ARGs) contributing to the first principal component (PC1). PC1 accounted for a significant portion of the total variance in the dataset. All top-loading genes share the same directional sign, indicating that these ARGs vary synchronously across the sampled locations.

### Environmental dissemination of ARGs

Detection frequencies for *bla_KPC_*, *bla_OXA-48_* and *bla_VIM7_* rose nominally in the post-Draft transect. However, detection frequency increases were non-significant for all carbapenemase genes. A paired Wilcoxon signed-rank test for PMMoV normalized *bla_KPC_* indicated an increased median concentration between the pre- and post CSO samplings (V = 614, p<0.001). Heterogeneity in effect could be observed between sites (Table 5, Figure 8).

**Figure 8.**
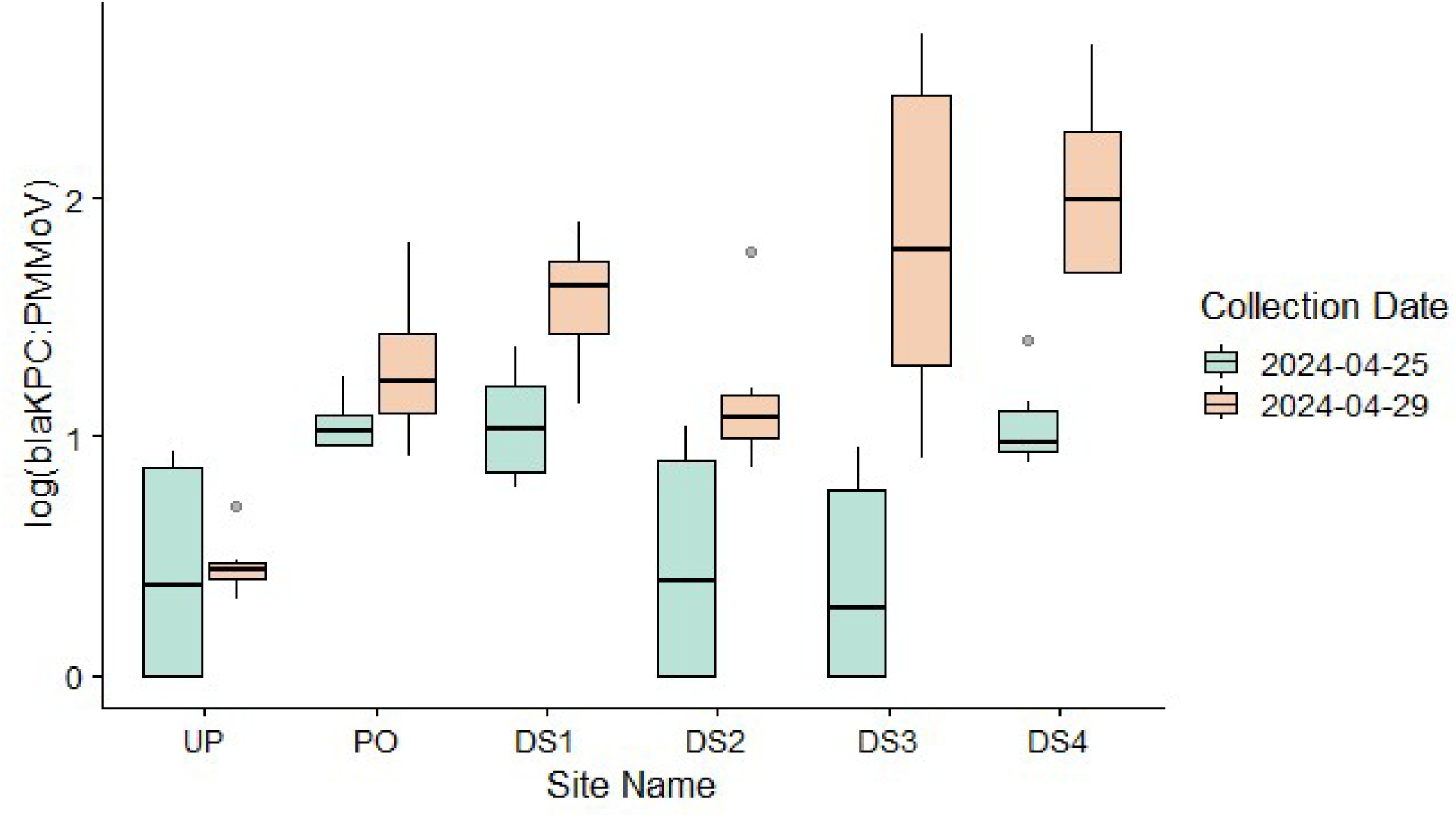
Log PMMoV normalized *blaKPC* concentrations before and after the 2024 NFL Draft and combined sewer overflow (CSO) at six sites in the Detroit River. Sites are arranged geographically along the x-axis.

**Table 5.** Median log PMMoV normalized *blaKPC* concentrations for each environmental sampling site before and after the 2024 NFL Draft and associated combined sewer overflow (CSO) event. The mean difference represents the magnitude of the concentration shift. P-values were calculated using site-specific paired Wilcoxon signed-rank tests

| Site | Median before | Median after | Mean difference | Wilcox p-value | Significance |
| --- | --- | --- | --- | --- | --- |
| DS1 | 1.030 | 1.631 | 0.522 | 0.031 | Significant |
| DS2 | 0.394 | 1.077 | 0.699 | 0.156 | Non-significant |
| DS3 | 0.286 | 1.787 | 1.429 | 0.031 | Significant |
| DS4 | 0.976 | 1.991 | 0.987 | 0.031 | Significant |
| PO | 1.023 | 1.233 | 0.238 | 0.094 | Non-significant |
| UP | 0.375 | 0.443 | 0.035 | 0.844 | Non-significant |

Detection frequencies were also calculated for ARGs included on the OpenArray® for both the pre- and post- CSO groupings. Detection frequency increased following the Draft weekend for 62.5% of ARGs assayed (Figure 9).

**Figure 9.**
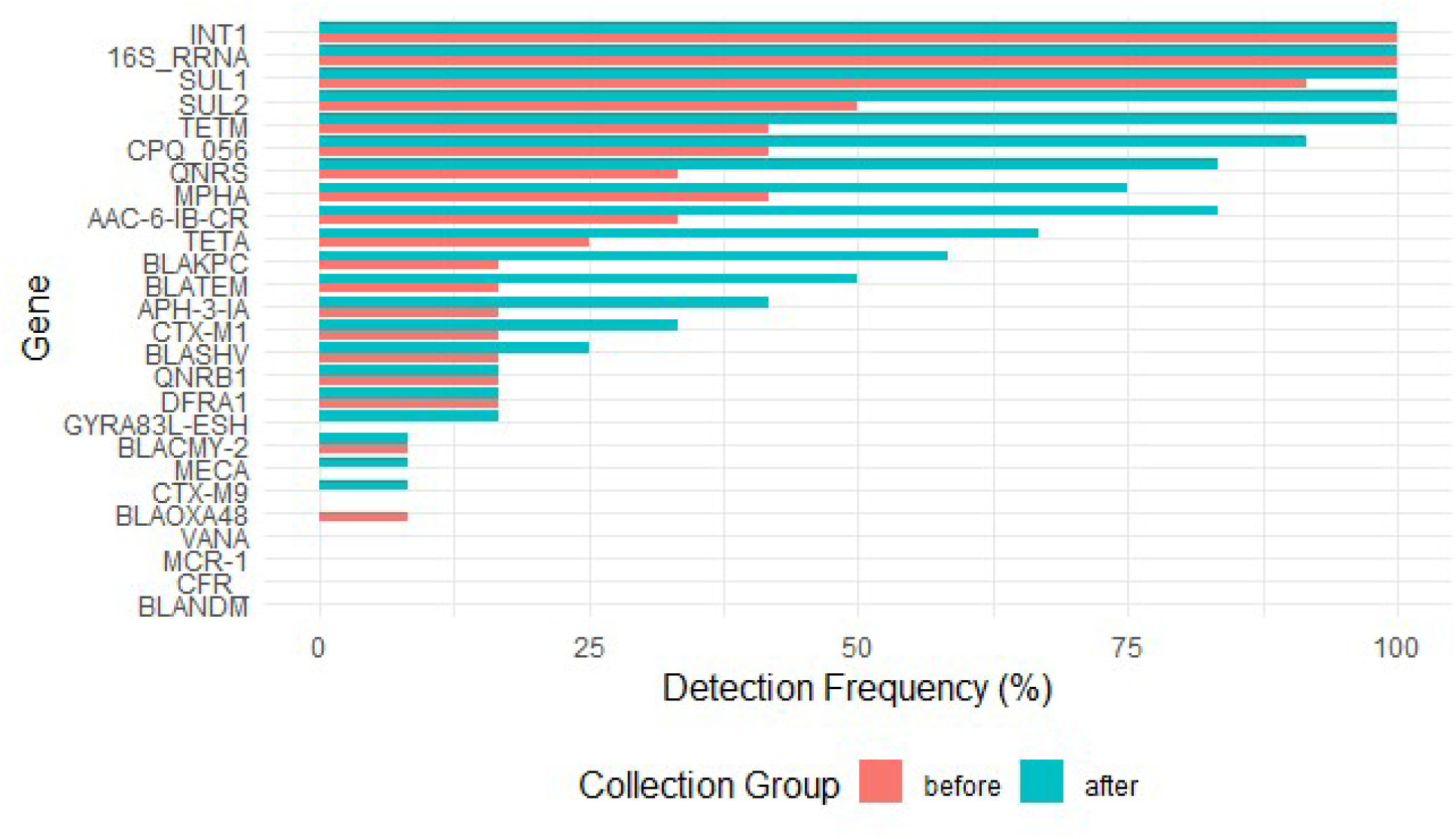
Detection frequences for ARGs in the Detroit River before and after the 2024 NFL Draft and combined sewer overflow event. Horizontal bars represent the percentage of samples positive for each genetic target before (red) and after (teal) the CSO event.

The binomial GLMM showed that sample collection timing had a significant effect on the probability of *crAssphage* detection (Estimate = 11.46, p = 0.0008), suggesting that the 2024 NFL Draft and CSO greatly increased detection probability. Additionally, the fitted model had high levels of variance for the random intercept (σ^2^ = 62.07, SD=7.88) and the random slope for the “after” group (σ^2^ =6.36, SD=2.52), indicating large differences in baseline detection probabilities and Draft effect between sites. Interaction terms between ARG and sampling group were not statistically significant, indicating that the relative magnitude of the post-Draft increase did not differ significantly among genes. Post-hoc pairwise comparisons for each gene included in the model found that detection probability increased for 6 genes including: *crAssphage, AAC(6’)-lb-cr, qnRS, tetA, mph(A),* and *bla_KPC_.* (Table S26). Increased detection probability post-Draft was corroborated by the paired Wilcoxon signed-rank test. Ten ARGs displayed statistically significant increases in concentration following the event, with no ARGs showing a significant decrease in abundance (Table S29).

The GLMM investigating site heterogeneity identified PO as the site with the highest detection probability, consistent with a point source of pollution (Table S30). An additional 4 ARGs showed statistically significant increases in concentration when the PO site was removed from the Wilcoxon signed-rank test (Table S33), suggesting that constitutively high ARG concentration at PO was masking significant increases in ARG concentration at other sites (Figure 10).

**Figure 10.**
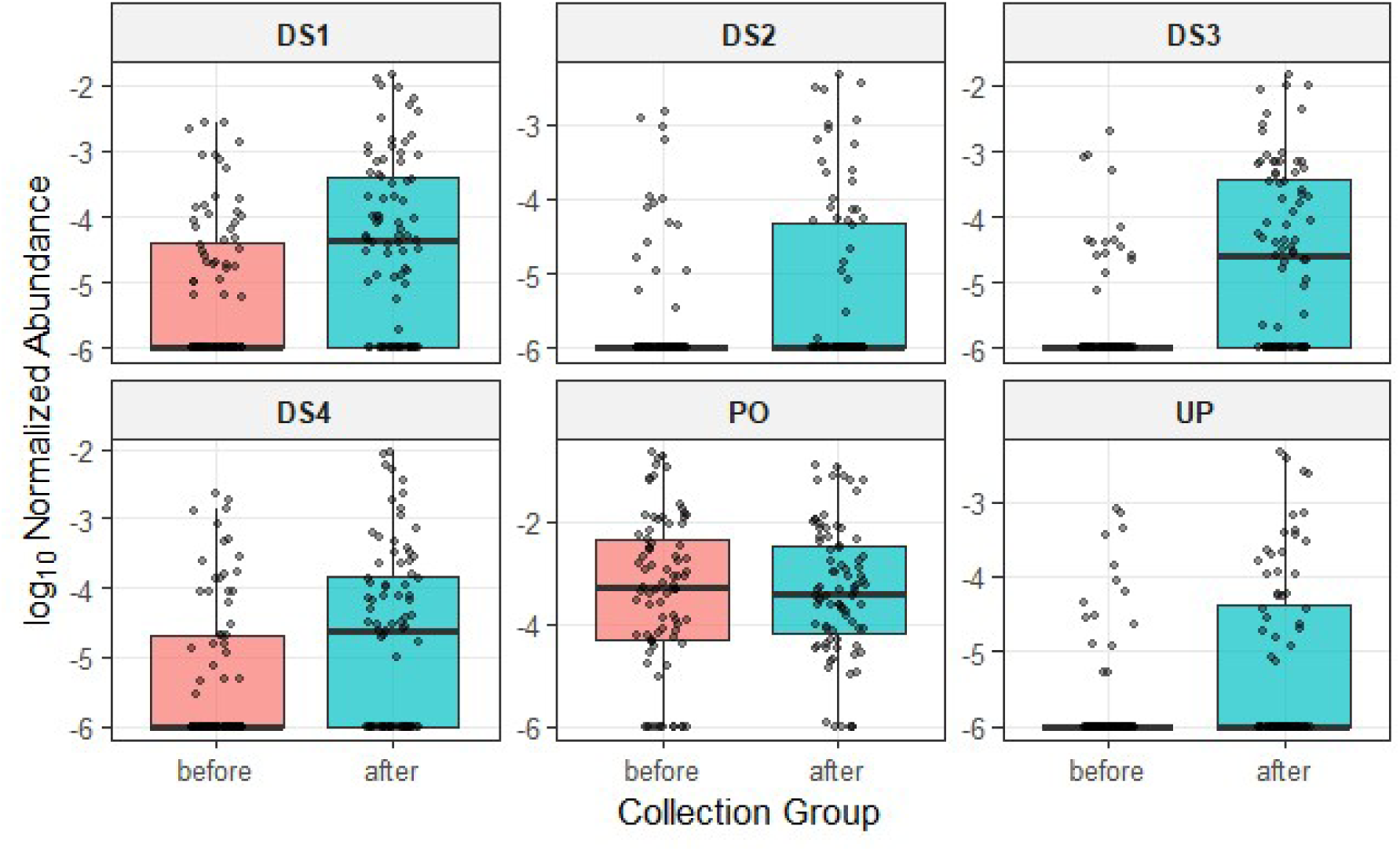
Log 16S rRNA normalized ARG abundances at six sites in the Detroit River before and after the 2024 NFL Draft and CSO event. Box plots represent the median and interquartile range of Log 16S rRNA normalized ARG concentrations with individual measurements over laid as a jitter. Facets represent each sampling site (DS1–DS4, PO, and UP). All sites save for PO show an increase in ARG abundance following the CSO.

### Method comparison

In wastewater, detection was 100% for all targets shared across platforms. In environmental samples, OpenArray showed lower sensitivity, missing low abundance targets detected by RT-qPCR (Table 6). Bland-Altman analysis showed that *bla_KPC_* had the narrowest LoA with *bla_NDM_* and *bla_OXA-48_* having wider LoAs (Figure S19). Both *bla_KPC_* and *bla_NDM_* had positive bias while *bla_OXA-48_* showed negative bias. Fold change analysis showed similar results, with the OpenArray having higher measurements in wastewater samples for *bla_KPC_* and *bla_NDM_* but lower for *blaOXA-48* (Figure S20).

**Table 6.** Comparison of RT-qPCR and nanofluidic RT-qPCR performance for detection of select antimicrobial resistance genes (ARGs) in different sample matrices

| Target | Matrix | No. Observations | RT-qPCR Positive | OpenArray Positive | Detection Gap | Kappa ($\kappa$) | p-value |
| --- | --- | --- | --- | --- | --- | --- | --- |
| *bla*<sub>KPC</sub> | Global | 42 | 42 (100%) | 31 (73.8%) | -26.20% | 0.000* | NA |
|  | Wastewater | 18 | 18 (100%) | 18 (100%) | 0% | 1.000** | NA |
|  | Environmental | 24 | 24 (100%) | 13 (54.2%) | -45.80% | 0.000* | NA |
| *bla*<sub>NDM</sub> | Global | 42 | 34 (81.0%) | 18 (42.9%) | -38.10% | 0.300 | 0.006 |
|  | Wastewater | 18 | 18 (100%) | 18 (100%) | 0% | 1.000** | NA |
|  | Environmental | 24 | 16 (66.7%) | 0 (0.0%) | -66.70% | 0.000* | NA |
| *bla*<sub>OXA-48</sub> | Global | 42 | 26 (61.9%) | 25 (59.5%) | -2.40% | 0.751 | < 0.001 |
|  | Wastewater | 18 | 18 (100%) | 18 (100%) | 0% | 1.000** | NA |
|  | Environmental | 24 | 8 (33.3%) | 7 (29.2%) | -4.10% | 0.516 | 0.011 |

The strength of monotonic agreement between the results of the OpenArray and RT-qPCR varied by target with the strongest agreement for *bla_KPC_* and weakest for *bla_OXA-48_* (Table 7). Pasing-Pablok and Demming regression revealed proportional bias for all targets (slopes >1.0), and large negative intercepts (Table S34). This pattern indicates a relative underestimation of ARG concentrations by the OpenArray® at lower analyte concentrations, transitioning to a relative overestimation at higher analyte concentrations. CCC was the highest for *bla_KPC_* (CCC=0.94) and lower for *bla_NDM_* (CCC = 0.88) and *bla*_OXA-48_ (CCC=0.75) (Table 7). Only *bla_KPC_* demonstrated substantial methodological agreement between platforms based on previously determined criteria^88^.

**Table 7.** Correlation and agreement of measurements for select ARGs assayed with RT-qPCR assays and nanofluidic RT-qPCR

| Target | n | Spearman | Pearson | CCC (95% CI) | Mean Bias (log) | LoA (95%) |
| --- | --- | --- | --- | --- | --- | --- |
| *bla*<sub>KPC</sub> | 16 | 0.91 | 0.97 | 0.94 (0.74–0.97) | 0.15 | -0.21 to 0.51 |
| *bla*<sub>NDM</sub> | 15 | 0.77 | 0.93 | 0.88 (0.69–0.94) | 0.29 | -0.77 to 1.34 |
| *bla*<sub>OXA-48</sub> | 18 | 0.64 | 0.82 | 0.75 (0.42–0.86) | -0.27 | -1.11 to 0.58 |

## Discussion

This study supports the utility of WES for monitoring and assessing the impact of mass gatherings on the spread of viral pathogens and antibiotic resistance genes. The 2024 NFL Draft served as a natural experiment. We measured the concentrations of public health relevant ARGs and seasonal respiratory pathogens in wastewater and in the Detroit River, which receives treated effluent from the Detroit-Windsor metropolitan region. WES detected no surge in respiratory viruses associated with the event, yet identified a transient, event-coincident rise in *bla_OXA-48_* in wastewater and broader ARG increases in surface waters during a concurrent wet-weather period with confirmed CSOs. A broad assessment of ARG concentrations in wastewater conducted over a limited timescale found that variability in the wastewater resistome was driven by site-level heterogeneity, potentially arising from differences in antimicrobial use between sewersheds.

### Viral pathogens

The 2024 NFL Draft did not act as a superspreader event for respiratory pathogens. Concentrations of SARS-CoV-2, IAV, IBV, and RSV remained steady or declined following the event. No evidence of increased transmission was observed despite the substantial increase in population in the monitored sewersheds and the mass gathering with increased transmission risk. Multiple factors may have contributed to the observed lack of increased transmission. The Draft was held outdoors and in large, well-ventilated buildings. This would have helped to limit the potential for aerosol transmission despite the large crowds^89^. Additionally, the Draft was held following the peak of the respiratory season in the northern hemisphere. Immunity for the most relevant strains of respiratory virus was likely high (acquired via infection or inoculation). The wastewater signal for most of the monitored pathogens was already low, indicating that prevalence was already low, reducing the probable number of infectious individuals attending Draft-related events. The lack of immediate increase in signal over the Draft weekend may be related to the relatively short duration of the Draft (∼3 days) in comparison with viral incubation times (∼5-6 days)^90^. The brevity of the mass gathering may not have allowed sufficient time for incubation or widespread transmission. Our analysis also indicated that Draft related transmission was low, with no significant increases in wastewater signal for any pathogens following the event. These findings demonstrate that large numbers of individuals can congregate without unreasonably increased risk of transmission if the gatherings occur outside of respiratory seasons or proper precautions to mitigate spread are implemented. The absence of a detectable signal does not imply complete absence of transmission; it does, however, suggest that the mass gathering event did not lead to a widespread increase in cases. Lack of extensive transmission is an important finding that can inform public health decision-making and provide feedback on event timing and the types of transmission mitigation strategies employed. Additionally, detection of transmission would be valuable for informed rapid action, especially when clinical case reporting is not feasible or delayed.

### Event-associated increases in carbapenemase genes

Unlike the monitored respiratory viruses, carbapenemase genes increased measurably following the mass gathering, driven by a large increase in *bla_OXA-48_* signal. The mass gathering event and influx of visitors were associated with a transient spike in the concentration of a clinically important ARG in the wastewater system. A lack of increase in any carbapenemase gene concentration at the spatially distinct control sites increases the likelihood that Draft-related travel was responsible for the increase rather than seasonal variability in ARG concentration. The increase in *bla_OXA-48_* observed in the present study may be partially explained by Draft attendees arriving from regions where OXA-48-like carbapenemases are more prevalent. These include parts of Europe, Africa, and the Middle East^92^. International travel is an important driver of the dissemination of OXA-48-like carbapenemases^93,94^ and in non-endemic areas, infections are often imported, though this modality may be changing^95^. Our results indicated that the increase in *bla_OXA-48_* was transient with no evidence of a sustained increase. However, the large increase in *bla_OXA-48_* concentration was notable, especially given that the baseline ARG signal was highly variable. This demonstrates that WES for ARGs may be sensitive to short term perturbations and that mass gatherings may induce short term changes in load for select ARGs. Wastewater flow was found to be a significant positive predictor of normalized ARG concentration, potentially a result of surface inputs to the sewer system or the mobilization of sewer biofilms^96,97^.

### Stable resistome with site-level heterogeneity

Multivariate analysis consistently indicated that the resistome structure in wastewater was dominated by sampling site characteristics rather than changes associated with the 2024 NFL Draft. This suggests that the influx of visitors during the 2024 NFL Draft did not significantly affect the resistome of the monitored sites in a way that was detectable in this analysis. Likely, sewershed level drivers such as population demographics, hospital effluent volumes, land use characteristics (i.e., amount of animal agriculture), and antibiotic prescription rates played a key role in determining the resistome. This was further supported by PCA loadings, which were dominated by common markers of anthropogenic pollution, like *int1* and *sul1*^98,99^. These ARGs co-varied across sites and sampling periods, indicating that the variation was driven by baseline pollution levels. Most PC1 loadings had the same sign, supporting the notion of covariance of ARGs being driven by variability in baseline pollution. ARG concentrations at the JEFF interceptor (the sampling site most closely linked to Draft activities) were not influenced by the Draft.

Conclusions from this analysis must be interpreted with caution as group sizes were uneven, sample sizes were small, and the study period was short. Combined, these limit the statistical power of the current analyses to detect relatively small changes in ARG concentration. Additionally, the OpenArray® has few targets compared to the diversity of ARGs, limiting our ability to detect ARGs that may have been introduced by the Draft.

Incorporation of ultra deep metagenomic sequencing may allow for the detection of novel, and low-abundance ARGs^100^. Longer-term data would help to confirm any Draft-associated changes. The results of this study are not generalizable. Mass gathering events still have the potential to contribute to ARG dissemination, especially if they are associated with travel between jurisdictions with different antimicrobial prescribing practices.

### Environmental dissemination of ARGs

Analyses revealed an increase in detection frequency and normalized concentration for multiple ARGs in the Detroit River following the 2024 NFL Draft weekend. The Draft event coincided with heavy precipitation, driving a CSO event. Detection probability increased for clinically relevant ARGs including: *AAC(6’)-lb-cr, qnRS, tetA, mph(A),* and *bla_KPC_*. These ARGs are associated with a variety of antimicrobial agents, indicating broad, rather than targeted increases in ARG detection following the Draft weekend. No gene × group interaction terms were significant in the GLMM, suggesting that the magnitude of the increase was consistent between targets. The rise in crAssphage concentrations in the post-Draft environmental samples and uniformity of detection probability increases across gene targets indicate that sewage contamination from the CSO is a major cause of the observed increase in ARG detections and abundance in the environmental samples. CSO events in isolation have been found to increase environmental concentrations of ARGs^101,102^. Mass gathering events can contribute to increased wastewater volumes and ARG loads^103^. However, given that OpenArray® -based analysis of wastewater samples did not detect a statistically significant increase in ARG concentrations attributable to the NFL Draft, the observed increase in ARG in the environment was likely caused by the rainfall-driven CSO events. The coincidence of the mass gathering event and the CSO may have introduced ARGs to the environment, imported to the region by event attendees, thus altering the watershed resistome. Such alterations have been observed in previous studies^104^.

Heterogeneity was observed among the sites of the sampling transect, with site-specific responses to the CSO and mass gathering. Samples collected at the outlet of the WEWTP exhibited evidence of saturated detection, accompanied by the highest normalized ARG concentrations. These findings support that the WWTP outlet was a major source of ARGs in the receiving waters with constitutively high ARG concentrations.

Inclusion of samples collected at the point source in data analysis obscured more subtle changes in ARG concentrations at other sites. This observation indicates the importance of site selection and highlights the complexity of interpretation for WES data in systems that are influenced by persistent inputs. In these systems, measuring baseline contamination levels is inherently challenging. Findings also indicated that the influence of the CSO was not limited to the pollution source. Increased concentrations of ARGs were found at the site upstream of the known WWTP outlet as well as the downstream sites. This observation could be explained by environmental sewage contamination originating from CSO events that occurred upstream within the Huron-Erie Corridor. Mass gatherings may increase the consequences of rainfall events in combined sewer systems, leading to alterations in the resistome of the receiving waters. Such alterations may have consequences for downstream utilization of water resources^105^.

Limitations of this analysis arise from the restricted temporal sampling and reliance on RT-qPCR for ARG measurement. RT-qPCR cannot detect which pathogen is hosting an ARG or if they are hosted on mobile genetic elements. Future work should incorporate longer-term monitoring efforts, hydrological modelling for better source tracking and should include metagenomic sequencing to help resolve ARG host identity.

### Comparison between OpenArray® and RT-qPCR

Comparison of the OpenArray ®and RT-qPCR quantification of three carbapenemase genes across two sample matrices ascertained the validity of the OpenArray® platform for ARG surveillance. The OpenArray® can be used for ARG trend assessment in concentrated wastewater matrices, but it is not directly quantitatively interchangeable with RT-qPCR, due to lower sample volumes. Agreement between the two methods was dependent on ARG abundance in the sample measured. Concordance between methods was 100% in high-concentration wastewater samples but was poor for environmental samples. Disagreement arose from non-detects by the OpenArray® rather than quantitative disagreement between methods. This observation is consistent with the approximately 600-fold smaller reaction volume (33nL vs 20μL) for the OpenArray®, which lowers detection probability at low copy numbers. Thus, the OpenArray® has limited utility for environmental monitoring without upstream concentration in the workflow^106–108^.

Assessment of quantitative agreement between OpenArray® in high concentration matrices also revealed significant discrepancies between methods. Passing-Bablok and Demming regression both revealed systematic proportional and constant bias between methods. Slopes greater than 1 and large negative intercepts are consistent with underestimation of samples at low concentrations and progressive overestimation at higher concentrations (consistent with nanolitre scale inputs, but a more efficient assay).

Agreement between the OpenArray® and RT-qPCR platforms was target dependent with *bla_KPC_* showing the strongest quantitative agreement (CCC=0.94), indicating methodological interchangeability. However, for *bla_NDM_* and *bla_OXA-48_*, overall agreement between methods was much lower (CCC=0.88 and CCC=0.75, respectively). For these targets, OpenArray® and RT-qPCR methods are not quantitatively interchangeable.

However, strong correlations were found between OpenArray® data and RT-qPCR data for all tested targets (Table 7). These correlations support reliable trend tracking for OpenArray® and RT-qPCR despite differences in the absolute concentration measured.

Tracking of trends over time is an important aspect of both wastewater-based surveillance and environmental monitoring, and despite the apparent quantitative issues with the OpenArray® platform, trend agreement is encouraging for the future development of the platform. The multitude of ARG targets to be monitored and high relative abundance in wastewater matrices make OpenArray® a promising platform for ARG monitoring in high concentration matrices with potential future applications for environmental monitoring with the implementation of pre-amplification and further methodological confirmations.

Low quantitative concordance between OpenArray® and RT-qPCR for *bla_OXA-48_* may help to explain why OpenArray® analysis failed to detect the spike in *bla_OXA-48_* that was detected via RT-qPCR. Optimization of multiplexed RT-qPCR allows for improved assay performance while cycling conditions for OpenArray® are set and uniform for all targets included on the chip. Additionally, OpenArray® analysis of the wastewater resistome relied on a limited number of samples while RT-qPCR benefited from a year-long survey of carbapenemase gene concentrations and a more sophisticated ITS modelling approach that attempted to account for baseline variability in carbapenemase concentrations.

## Conclusion

The main goals of this investigation were to determine whether WES is a viable tool for monitoring pathogen transmission dynamics associated with mass gathering events and to determine if the 2024 NFL Draft led to increased pathogen transmission or ARG dissemination. Findings provide support for the use of WES in this application, with results of respiratory pathogen monitoring indicating no discernible increase in transmission associated with the 2024 NFL Draft. WES to monitor trends in AMR is particularly promising as it can capture a dimension of infectious diseases that are more difficult to track via clinical surveillance methods. Additionally, environmental dissemination of ARGs is impossible to track via clinical surveillance but can have implications for population health. The ability of WES to detect transient resistome shifts makes the argument that it is a valuable public health surveillance tool during mass gatherings. A recent study investigated ARG load, diversity and dissemination in relation to mass gathering events^109^. Investigators found that the overall level of ARGs did not increase in relation to mass gathering events. More importantly, they determined that mass gatherings could introduce ARGs to local sewer systems that were previously absent. Findings of our investigation are similar, albeit more methodologically limited, as we only measured select ARGs. Both our investigation and the previous work found that ARG burden did not increase overall as a result of the mass gathering but that the concentration of select genes might increase in a transient manner and that the event itself may have led to the introduction of locally novel ARGs. Notably, the wastewater signal observed in this study was not driven by a uniform increase across all monitored targets. Instead, the response was selective, emphasizing the importance of targeted surveillance strategies and careful interpretation of WES data in event-based contexts. Dissemination of locally novel ARGs is concerning from a public health perspective as it is likely to lead to increased numbers of antibiotic-resistant infections or infections with novel resistance profiles. The combined evidence supports WES as a reliable tool for validating the safety of mass gatherings regarding respiratory spread while providing early insight into AMR dynamics and environmental dissemination under infrastructure stress. Integrating WES with hydrologic context (rainfall, CSO telemetry) can guide real-time operational responses (e.g., CSO management, advisories) during future events.

## Data Availability

All data produced in the present study are available upon reasonable request to the authors

## Acknowledgements

Operators, Laboratory Team, and Managers of the following wastewater treatment facilities: Wastewater Resource Recovery Facility, Great Lakes Water Authority; Pollution Control, City of Windsor; Pollution Control Centre, Municipality of Leamington, Water Pollution Control Plants, City of Chatham and City of Thunder Bay. Funding in support of the Ontario Wastewater Surveillance Initiative was provided by the Ontario Ministry of Environment, Conservation, and Parks. Additional support was provided by the Public Health Agency of Canada’s National Wastewater Surveillance Program (Contribution Agreement 2324-HQ-000197 to CGW, DDH, RS and RMM). The views expressed herein do not necessarily represent the views of the Public Health Agency of Canada.

