## Supplementary Materials for "Application of wastewater and environmental surveillance for pathogenic agents during the 2024 National Football League (NFL) Draft in Detroit, Michigan (USA)"

### Supplemental methods

#### S1. Detailed site information

WRRF- The Water Resource Recovery Facility (WRRF) is a large wastewater treatment plant operated by the Great Lakes Water Authority (GLWA) in Detroit. The plant serves much of Detroit and the surrounding suburban area totaling roughly 3.1 million individuals. The plant is fed by three main interceptors including the Detroit River Interceptor (JEFF), the North Interceptor-East Arm (NIEA) and the Oakwood-Northwest-Wayne County Interceptor (OAK).

WEWTP- A wastewater treatment facility in Windsor-Essex which treats the waste of roughly 180,000 residents.

CHWW- The Chatham Water Pollution Control Plant serves approximately 48,000 residents[1] and is located approximately 75km to the east of the Detroit-Windsor region. This community was unlikely to have seen a large influx of travellers/sports fans in relation to the 2024 NFL Draft in Detroit and thus can serve as a control site. Any trends in respiratory illness observed in Chatham may be attributed to seasonal fluctuations rather than the mass gathering itself.

LPCC- The Leamington Pollution Control Centre treats sewage from the urban area of Leamington, and waste transported from local septic systems. Leamington is a small agricultural city in southwestern Ontario with a population of 29,680 permanent residents[2]and approximately 7000 temporary residents[3], many of which work as agricultural labourers.

TB- The Water Pollution Control Plant (WPCP) serves the city of Thundery Bay, a mid-sized city located in northeastern Ontario with a population of 108,843[4]. The plant serves approximately 97524 residents.

Detroit River Transect- a six site transect of the Detroit River upstream and downstream of effluent discharge of a wastewater treatment facility. The final six sites were selected for their proximity to the effluent discharge of a wastewater treatment facility (WEWTP). One site was used as a control and was located upstream of the outfall. Samples were also collected at the outfall and at four separate locations downstream of the outfall. Figure 1 (main manuscript) summarises the locations of the sampling sites in relation to the discharge. Samples were collected on 2024-04-25, prior to the 2024 NFL Draft which was hosted in Detroit, Michigan, and on 2024-04-29 following the end of the 2024 NFL Draft.

**S2. RT-qPCR assays for respiratory virus quantification**

**S2.1. Severe Acute Respiratory Syndrome Coronavirus 2 (SARS-CoV-2)**

Assays for SARS-CoV-2 targeted regions of the nucleocapsid (N) gene using US CDC primers and probes for the N1 and N2 regions[5]. Reagents were supplied by Integrated DNA Technologies (Coralville, IA, USA). Reactions contained 5µL of RNA template mixed with 10µL of Luna Universal Probe One-Step Reaction Mix (2X), 1µL Luna WarmStart® RT Enzyme Mix (20X) (Luna® One-Step RT-qPCR Kit, Massachusetts, USA), forward primer (final concentration of 300nM), reverse primer (final concentration of 300nM), and probe (final concentration of 150nM) in a final reaction volume of 20µL. Reaction inhibition was assessed using VetMAX XENO Internal Positive Control RNA (Applied Biosystems Corp., Waltham, MA, USA). Due to repeated incidence of inhibition with wastewater samples processed by filtration, template was diluted 1:5 in all reactions. Technical triplicates were run for detection of gene targets. Thermal cycling was performed using a MA6000 qPCR thermocycler (Aumintec, Richmond Hill, ON, Canada). RT was performed at 60 °C for 10 min, followed by polymerase activation at 95 °C for 1 min, and 45 cycles of denaturation, annealing/extension at 95 °C for 10 sec, then 60 °C (N1) or 63 °C (N2) for 30 sec, respectively. The EDX SARS-CoV-2 synthetic RNA standard (Exact Diagnostics, Fort Worth, TX,160 USA) was used to create a 7-point standard curve to quantify gene targets. No template controls yielded no amplification, and we report a limit of detection (LOD) of 5 gene copies of N1 per reaction containing 5 µL of template RNA.

**S2.2. Influenza A Virus (IAV)**

The concentration of IAV RNA in samples was measured using an established assay that targets the IAV M1- gene[6]. Reactions contained 4µL of RNA template mixed with 10µL of Luna Universal Probe One-Step Reaction Mix (2X), 1µL Luna WarmStart® RT Enzyme Mix (20X) (Luna® One-Step RT-qPCR Kit, Massachusetts, USA), forward primer (final concentration of 500nM), reverse primer (final concentration of 500nM), and probe (final concentration of 250nM) in a final reaction volume of 20µL. RT was performed at 55 °C for 10 min, followed by polymerase activation at 95 °C for 1 min, and 45 cycles of denaturation, annealing/extension at 95 °C for 10 sec, then 58 °C for 45 sec, respectively. No template controls yielded no amplification, and the LOD for the assay was determined at 4 gene copies of IAV per reaction containing 4µL of template RNA, corresponding to a greater than 95% probability of detection. Twist Synthetic Influenza H3N2 RNA control (Twist Bioscience, San Francisco, CA) was used to create an 8-point standard curve to quantify gene targets. Reaction inhibition was assessed using VetMAX XENO Internal Positive Control RNA (Applied Biosystems Corp., Waltham, MA, USA). Due to repeated incidence of inhibition with wastewater samples processed by filtration, template was diluted 1:5 or 1:10 in all reactions. Technical triplicates were run for detection of gene targets. Thermal cycling was performed using a MA6000 qPCR thermocycler (Aumintec, Richmond Hill, ON, Canada).

**S2.3. Influenza B Virus (IBV)**

The concentration of IBV RNA in samples was measured using an established assay [6]. Reactions contained 4µL of RNA template mixed with 10µL of Luna Universal Probe One-Step Reaction Mix (2X), 1µL Luna WarmStart® RT Enzyme Mix (20X) (Luna® One-Step RT-qPCR Kit, Massachusetts, USA), forward primer (final concentration of 1200nM), reverse primer (final concentration of 1200nM), and probe (final concentration of 300nM) in a final reaction volume of 20µL. RT was performed at 58 °C for 10 min, followed by polymerase activation at 95 °C for 1 min, and 45 cycles of denaturation, annealing/extension at 95 °C for 10 sec, then 58 °C for 45 sec, respectively. No template controls yielded no amplification, and the LOD for the assay was determined at 4 gene copies of IBV per reaction containing 4µL of template RNA, corresponding to a greater than 95% probability of detection. Twist Synthetic Influenza B RNA control (Twist Bioscience, San Francisco, CA) was used to create an 8-point standard curve to quantify gene targets. Reaction inhibition was assessed using VetMAX XENO Internal Positive Control RNA (Applied Biosystems Corp., Waltham, MA, USA). Due to repeated incidence of inhibition with wastewater samples processed by filtration, template was diluted 1:5 or 1:10 in all reactions. Technical triplicates were run for detection of gene targets. Thermal cycling was performed using a MA6000 qPCR thermocycler (Aumintec, Richmond Hill, ON, Canada).

**S2.4. Respiratory Syncytial Virus (RSV)**

RT-qPCR was used to quantify the concentration of RSV RNA within wastewater samples. The assay amplifies the N-gene for both RSV A and RSV B[7]. Reagents for RT-qPCR were obtained from Integrated DNA Technologies (Coralville, IA, USA). Reactions contained 4µL of total nucleic acid extracted from wastewater (diluted 1:5), mixed with 10µL of Luna

Universal Probe One-Step Reaction Mix (New England Biolabs, Ipswich, MA, USA), 1μL of Luna WarmStart RT Enzyme Mix (20×), forward primer (final concentration of 600nM), reverse primer (final concentration of 600nM), and probe (final concentration of 200nM). Reaction inhibition was assessed using the VetMAX XENO Internal Positive Control RNA (Applied Biosystems Corp., Waltham, MA, USA). Inhibition was consistently observed in wastewater samples and as such, RNA was diluted 1:5 for all reactions. Each sample was run in technical triplicate or quadruplicate using a MA6000 thermal cycler (Aumintec, Richmond Hill, ON, Canada). The reverse transcription step was performed at 55°C for 10 min, followed by polymerase activation at 95°C for 1 min and 45 cycles of denaturation. Annealing/extension occurred at 95°C for 10 sec, then 55°C for 45 sec, respectively. To create a 8-point standard curve, a synthetic DNA gBlock diluted in ultra-pure water was used. No template controls (NTC) and blanks yielded no amplification. The LOD for this assay was 1 copy $\mu L^{-1}$ of template as determined through examination of 20 replicate standard curves.

#### S2.5. RT-qPCR assays for ARG quantification

A pair of multiplex qPCR assays were used to measure the concentration of ARGs within samples including $bla_{NDM}$, $bla_{KPC}$,$bla_{VIM}$ $bla_{VIM7}$, and $bla_{OXA\text{-}48}$. The first multiplex assay targeted $bla_{KPC}$, $bla_{NDM}$, and $bla_{VIM7}$. The second targeted $bla_{VIM}$ and $bla_{OXA\text{-}48}$. Primers and probes were purchased from Integrated DNA Technologies (Coralville, IA, USA). Sequences for primers and probes are reported in Table S1. For the assay that queried $bla_{KPC}$, $bla_{NDM}$, and $bla_{VIM7}$, each 20μL reaction was comprised of 5μL of DNA template as well as 10μL of Luna Universal Probe One-Step Reaction Mix (2X) (Luna® One-Step RT-qPCR Kit, Massachusetts, USA), and forward primer (final concentration of 800nM), reverse primer (final concentration of 800nM), and probe (final concentration of 400nM) for each target. For this multiplex assay polymerase activation was performed at 95 °C for 20 seconds, followed by 45 cycles of denaturation, annealing/extension at 95 °C for 5 seconds, then 55 °C for 30 sec, respectively. For the assay that targets $bla_{VIM}$ and $bla_{OXA\text{-}48}$, each 20μL reaction was comprised of 5μL of DNA template as well as 10μL of Luna Universal Probe One-Step Reaction Mix (2X) (Luna® One-Step RT-qPCR Kit, Massachusetts, USA). The final 5 μL volume was made up of forward primer (final concentration of 1200nM for $bla_{VIM}$ and 600nM for $bla_{OXA\text{-}48}$), reverse primer (final concentration of 1200nM for $bla_{VIM}$ and 600nM for $bla_{OXA\text{-}48}$), and probe (final concentration of 600nM for both targets). For the second multiplex assay polymerase activation was performed at 95 °C for 20 seconds, followed by 45 cycles of denaturation, annealing/extension at 95 °C for 5 seconds, then 53 °C for 30 sec, respectively. Synthetic double stranded DNA controls (gBlocks™ (Integrated DNA Technologies, Coralville, IA, USA)) were serially diluted to produce an 8-point standard curve for quantification of the assayed ARGs. The sequence of the fragment can be found in Table S2.

The LOD for each gene target measured was ascertained through analysis of the lowest concentration points of the standard curves. The LOD was 25 copies/well for for $bla_{NDM}$, $bla_{KPC}$, $bla_{VIM}$ and $bla_{VIM7}$ corresponding to 5gc/μL of template DNA. However, for $bla_{OXA\text{-}48}$ the LOD was 5 copies/well, corresponding to 1 gc/μL of template DNA. The reported LODs correspond to a more than 95% probability of detection. The limit of quantification (LOQ) was also determined for each of the qPCR assays. The LOQ for the assay querying $bla_{NDM}$ was 25.6 copies/well. The LOQ for the assay querying $bla_{KPC}$ was 61.0 copies/well. The LOQ for the assay querying $bla_{VIM}$ was 2295.1 copies/well. The LOQ for the assay querying $bla_{VIM7}$ was 9.0 copies/well. The LOQ for the assay querying $bla_{OXA\text{-}48}$ was 4.5 copies/well. LOQs were determined using the $CV_{ln}$ method[8–10].

**Table S1.** Primer and probe sequences for qPCR assays for select carbapenemases

| Gene | Forward (5'->3') | Reverse (5'->3') | Probe (5'->3') | Reference |
|---|---|---|---|---|
| $bla_{KPC}$ | GGCCGCCGTGCAATAC | GCCGCCCAACTCCTTCA | FAM/TGATAACGC/ZEN/CGCCGCCAATTTGT/3IABkFQ | [11] |
| $bla_{NDM}$ | GCAAATGGAAACTGGCGACC | TACCGCCCATCTTGTCCTGA | HEX/TCGCACCGA/ZEN/ATGTCTGGCAGCACA/3IABkFQ | [12] |
| $bla_{VIM}$ | TTGATTGATACAGCTTGGGGTA | ACGGYGATGCGTACGTTGC | Cy5/GACGCGGTC/TAO/GTCATGAAAGTGCGT-/3IAbRQSp | [12] |
| $bla_{VIM7}$ | GAGTTGCTTCTTATTGATACAG | AGGGTGAGGTGTACGTTGC | Cy5/CCGACTCGA/TAO/TCGTCATGGAAGTGCGT-/3IAbRQSp | [12] |
| $bla_{OXA\text{-}48}$ | CTTAAACGGGCGAACCAAGC | GTTCATCCTTAACCACGCCC | FAM/TTCCCAATA/TAO/GCTTGATCGCCCTCGATT-/3IAbRQSp | [12] |

**Table S2.** gBlocks™ sequences for used as synthetic positive controls and for the generation of standard curves

| Assay | gBlocks™ Sequence (5’->3’) |
|---|---|
| *aph(3')-Ia* | CCGTTTCTGTAATGAAGGAGAAAATTCACCGAGGCAGTTCCATAGGATGGCAAGATCCTGGTATCGGTCTGCGATTCC<br>GACTCGTCCAACATCAATACAACCTATTAATTTCCCCTCGTCAAAAATAAGGTTATCAAGTGAGAAATCACCA |
| *aac(6')-Ib-cr* | ATCCAAACGGACCCGTCGCCGAGCAACTTGCGAGCGATCCGATGCTACGAGAAAGCGGGGTTTGAGAGGCAAGGT<br>ACCGTAACCACCCCATATGGTCCAGCCGTGTACATGGTTCAAACACGCCAGGCATTCGAGCG |
| *bla*$_{IMP}$ | ACCAGTTTTGCCTTACCATATTCAGACTTCCATAATTCAGCGGACTTTGGCCATGCTTTAATATTTGCGTCACCCAAATTG<br>CCTAAACCGTACGGTTTAACAAAGCAACCACCGAATAACGTTTTCCTTTCAGGCAGCCAAACCACTACGTTATCTGGA<br>GTGT |
| *bla*$_{VIM}$ | GCATATCGCAACGCAGTCGTTTGATGGCGCAGTCTACCCGTCCAATGGTCTCATTGTCCGTGATGGTGATGAGTTGCTT<br>TTGATTGATACAGCGTGGGGTGCGAAAAACACAGCGGCACTTCTCGCGGAGATTGAGAAGCAAATTGGACTT |
| *bla*$_{CMY-2}$ | AATACTGGCCAGAACTGACAGGCAAACAGTGGCAGGGTATCCGCCTGCTGCACTTAGCCACCTATACGGCAGGCGG<br>CCTACCGCTGCAGATCCCCGATGACGTTAGGGATAAAGCCGCATTACTGCATTTTTATCAAAACT |
| *bla*$_{SHV}$ | ATGTTTTTCGCTGACCGGCGAGTAGTCCACCAGATCCTGCTGGCGATAGTGGATCTTTCGCTCCAGCTGTTCGTCACC<br>GGCATCCACCCGCGCACGCACTGCGCCGCAGAGCACTACTTTAAAGGTGCTCATCATGGGAAAGCGTTCATCGGC<br>GCGCCAGGCGGTCAGCGTGCGGCCGCTGGCCAGATCCA |
| *bla*$_{TEM}$ | TACTCACCAGTCACAGAAAAGCATCTTACGGATGGCATGACAGTAAGAGAATTATGCAGTGCTGCCATAACCATGAGTG<br>ATAACACTGACTCTAACTTACTTCTGACAACGATCGGAGGACCGAAGGAGCTAACCGCTTTT |
| *bla*$_{CTX-M1}$ | AGTCGGGAGGCAGACTGGGTGTGGCATTGATTAACACAGCAGATAATTCGTCAATACTTTATCGTGCTGATGAGCGCTT<br>TGCGATGTGCAGCACCAGTAAAGTGATGGCCGTGGCCGCGGTGCTGAAGAAAAGTGAAAGCGAACCGAATCTGTTAA<br>ATCAGCGAGTTGAGATCAAAAAATC |
| *bla*$_{CTX-M9}$ | GCGCCGCTTTATGCGCAGACGAGTGCGGTGCAGCAAAAGCTGGCGGCGCTGGAGAAAAGCAGCGGAGGGCGGC<br>TGGGCGTCGCGCTCATCGATACCGCAGATAATACGCAGGTGCTTTATCGCGGTGATGA |
| *mcr-1* | TGGTCGTATCATAGACCGTGCCATAAGTGTCAGTAAAATAACTGGTCACCGCGCCCATGATTAATAGCAAAATCAACAC<br>AGGCTTTAGCACATAGCGATACGATGATAACAGCGTGGTGATCAGTAGCATCGCGCCAAAGAGCACGACAGCGATCG<br>TCAGCACAAAGCCGAGATTGTCCGCGATGGGATAGGTTTGGCTGA |
| *gyrA83L-ESh* | TGCCCGTGTCGTTGGTGACGTAATCGGTAAATACCATCCCCACAGTGACTTGGCGGTCTATAACACGATCGTCCGCAT<br>GGCGCAGCCATTCTCGCTGCGTTATATGCTGGTAGACGGTCAGGGTAACTTCGGTTCTATCGACGGCGACTCT |
| *QnrB1* | CTGGTTTTGTAGCGCATATATCACGAATACCAATCTAAGCTACGCCAATTTTTCGAAAGTCGTGTTGGAAAAGTGTGAGCT<br>GTGGGAAAACCGTTGGATGGGTGCCCAGGTACTGGGCGCGACGTTCAGTGGTTCAGATCTCTCCGGCGGC |
| *qnrS* | AAGTTGGCATTGTTGGAAACTTGCATCACGAAGATCTGCGACATCAAAGTGGCAGCCTTCGATATCACCCTGTTCAATG<br>AACTCACAGTTGACGAATGTCGTATCACGCAAGTTAGCACGTCGAAAGT |
| *vanA* | CTGCAGTACGGAATCTTTCGTATTCATCAGGAAGTCGAGCCGGAAAAAGGCTCTGAAAACGCAGTTATAACCGTTCCC<br>GCAGACCTTTCAGCAGAGGAGCGAGGACGGATACAGGAAACGGCAAAAAAAATATATAAAGCGCTCGGCTGTA |
| *mphA* | CGCGGGTGCTGGCAATGCTCAAGAATCGCCTGCCGTTCGCGGTGCCGGACTGGCGCGTGGCCAACGCCGAGCTC<br>GTTGCCTATCCCATGCTCGAAGACTCGACTGCGATGGTCATCCAGCTTGATT |
| *cfr* | ATTACCATATAATTGACCACAAGCAGCGTCAATATCAATCCCAAATGTACTTCTAATTGTGACATGGATACCAGCAGACTT<br>CAAAACTTTGTAAAAGGCTTCTACCTGCCCTTCGTTTGCTTCTCCATACATCTCAGGTGCACTTATTGTAGG |
| *mecA* | AATTTTCAATATGTATGCTTTGGTCTTTCTGCATTCCTGGAATAATGACGCTATGATCCCAATCTAACTTCCACATACCATCT<br>TCTTTAACAAAATTAAATTGAACGTTGCGATCAATGTTACCGT |
| *int1* | TCTCCTGAAGCCAGGGCAGATCCGTGCACAGCACCTTGCCGTAGAAGAACAGCAAGGCCGCCAATGCCTGACGAT<br>GCGTGGAGACCGAAACCTTGCGCTCGTTCGCCAGCCAGCTCAGAAATGCCTCGACTTCGCTGCTGCCCAAGGTTG<br>CCGGGT |
| *16S rRNA* | AGCGGTGGAGCATGTGGTTTAATTCGATGATACGCGAGGAACCTTACCAGGGCTTGACATATACAGGACGTGCTTAGA<br>GATAAGTATTCCTCTTCGGAGGCTTGTATACAGGTGCTGCATGGTTGTCGTCAGCTCGTGCCGTGAGGTGTTGGGTTAA<br>GTCCCGCAACGAG |
| *sul1* | GTCTCGGTGTCGCGGAAATCCTTCTTGGGCGCCACCGTTGGCCTTCCTGTAAAGGATCTGGGTCCAGCGAGCCTTG<br>CGGCGGAACTTCACGCGATCGGCAATGGCGCTGACTACGTCCGCACCCAC |
| *sul2* | TTTTTTCTGGGGGCTGCTCCCGAAACCTCGCTCTCGGTGCTGGCGCGGTTCGATGAATTGCGGCTGCGCTTCGATTT<br>GCCGGTGCTTCTGTCTGTTTCGCGCAAATCCTTTCTGCGCGCGCTCACA |
| *TetA* | TTGCCGACGGCACAGGCTACATCCTGAGTGCCTTCGCGACACGGGGATGGATGGCGTTCCCGATCATGGTCCTGTC<br>TGCTTCGGGTGGCATCGGAATGCCGGCGCTGCAAGCAATGTTGTCCAGGCAGGTG |
| *TetM* | AAAACAGATTTTACAGTCCGTCACATTCCAACCATACAATCCTTGTTCACAACCATAGCGTATCCCTTCCATAACTGCATT<br>TTGAAATGATTGATTTAAGTATCCAAGAGAAACCGAGCTCTCATA |
| *dfrA1* | AATGCTCCCATTGATTCAAAAGTCTTGCGTCCAACCAACAGCCATTGGTTATAGGTAATAGCTTTAAACAGGAGCTGTTC<br>ACCTTTGGCACTCCATGGAATATCAGGGCCATTCCCGATAACTCCA |
| *bla*$_{KPC}$ | GCTGAGGAGCGCTTCCCACTGTGCAGCTCATTCAAGGGCTTTCTTGCTGCCGCTGTGCTGGCTCGCAGCCAGCAGC<br>AGGCCGGCTTGCTGGACACACCCATCCGTTACGGCAAAAATGCGCTGGTTCCGTGGTCACCCATCTCGGAAAAATAT<br>CTGACAACAGGCATGACGGTGGCGGAGCTGTCCGCGGCCGCCGTGCAATACAGTGATAACGCCGCCGCCAATTTG<br>TTGCTGAAGGAGTTGGGCGGCCCGGCCGGGCTGACGGCCTTCATGCGCTCTATCGGCGATACCACGTTAATTAT |
| *bla*$_{NDM}$ | TGAAATCCGCCCGACGATTGGCCAGCAAATGGAAACTGGCGACCAACGGTTTGGCGATCTGGTTTTCCGCCAGCTC<br>GCACCGAATGTCTGGCAGCACACTTCCTATCTCGACATGCCGGGTTTCGGGGCAGTCGCTTCCAACGGTTTGATCGT<br>CAGGGATGGCGGCCGCGTGCTGGTGGTCGATACCGCCTGGACCGATGACCAGACCGCCCAGATCCTCAACTGGAT |

| | |
|---|---|
| | CAAGCAGGAGATCAACCTGCCGGTCGCGCTGGCGGTGGTGACTCACGCGCATCAGGACAAGATGGGCGGTATGGA CGCGCTGCAT |
| *bla*$_{OXA-48}$ | CAGCAAGGATTTACCAATAATCTTAAACGGGCGAACCAAGCATTTTTACCCGCATCTACCTTTAAAATTCCCAATAGCTT GATCGCCCTCGATTTGGGCGTGGTTAAGGATGAACACCAAGTCTTTAAGT GGGATGGACA |
| *bla*$_{VIM}$ | GATGGTGATGAGTTGCTTTTGATTGATACAGCGTGGGGTGCGAAAAACACAGCGGCACTTCTCGCGGAGATTGAAAAG CAAATTGGACTTCCCGTAACGCGTGCAGTCTCCACGCACTTTCATGACGACCGCGTCGGCGGCGTTGATGTCCTTCG GGCGGCTGGGGTGGCAACGTACGCATCACCGTCGACACGCCGGCT |
| *bla*$_{VIM-7}$ | CGATGCTGATGAGTTGCTTCTTATTGATACAGCGTGGGGGGCGAAGAACACGGTAGCCCTTCTCGCGGAGATTGAAAA GCAAATTGGACTTCCAGTAACGCGCTCAATTTCTACGCACTTCCATGACGATCGAGTCGGTGGAGTTGATGTCCTCCG GGCGGCTGGAGTGGCAACGTACACCTCACCCTTGACACGCCAGC |
| PMMoV | GGTGGCAGCAAAGGTAATGGTAGCTGTGGTTTCAAATGAGAGTGGTTTGACCTTAACGTTTGAGAGGCCTACCGAAGC AAATGTCGCACTTGCATTGCAACCGACAATTACATCAAAGGAGGAAGGTTCGTTGAAGATTGAAATGTCTGA |

### S2.6. RT-qPCR assay for Pepper Mild Mottle Virus (PMMoV) quantification

Levels of Pepper Mild Mottled Virus (PMMoV) within the wastewater were also assayed using RT-qPCR. PMMoV is a plant pathogen and an indicator that is correlated with human fecal material[13–15] that can be used to normalize results for variability due to flow or other parameters. RT-qPCR reactions for quantification of PMMoV, were made up of 2.5μL of sample,10μL of Luna Universal Probe One-Step Reaction Mix (2X), 1μL Luna WarmStart® RT Enzyme Mix (20X) (Luna® One-Step RT-qPCR Kit, Massachusetts, USA), 3.5μL of PCR grade water and 3μL of forward primer, reverse primer, and probe each in a final concentration of 200nM. The primers and probes used for the quantification of PMMoV have been described previously[16]. Cycling conditions for PMMoV quantification are as follows: reverse transcription at 55 °C for 10 minutes, enzyme activation at 95°C for 1 minute, 40 cycles of denaturation and annealing/extension at 95°C for 10 seconds and 55°C for 30 seconds, respectively. A standard curve for the quantification of PMMoV was produced through the evaluation of a 7 concentration dilution series of a custom gBlock™. It was determined that the PMMoV assay had an LOD of 5 copy·$\mu L^{-1}$ of template RNA, which corresponds to a greater than 95% probability of detection. This LOD was calculated by evaluation of 20 replicate 7-point standard curves. Inhibition was measured with VetMAX XENO Internal Positive Control RNA (Applied Biosystems Corp., Waltham, MA, USA). The presence of inhibition was detected at a frequency that necessitated the template be diluted 1:5 or 1:10 in all reactions. Extraction blanks and qPCR negative controls yielded no amplification for any assay. All RT-qPCR and qPCR reactions were performed in triplicate using a MA6000 qPCR thermocycler (Aumintec, Richmond Hill, ON).

### S3. Detailed nanofluidic qPCR (OpenArray ®) methods

### S3.1. Development of the nanofluidic qPCR card using TaqMan® assays in the OpenArray ® platform

Assays for the quantification of clinically important and locally relevant ARGs and genes associated with mobile genetic elements (MGEs) were selected for inclusion on the OpenArray® card (Table 1, main manuscript). Two additional targets *16S rRNA* and

*CrAssphage* were included to proxy fecal/bacterial concentrations for the purpose of normalizing ARG concentrations[17]. Previously developed assays for the targets of interest were identified through a literature search (Table S3). Oligonucleotide sequences for primers and probes were modified to be compatible with TaqMan chemistry (MGB moiety) using Primer Express3 (Applied Biosystems, Life Technologies Corporation, Carlsbad, CA, USA). The specificity of the resulting assays and theoretical amplicon length were checked using the Basic Local Alignment Search Tool (BLAST; https://blast.ncbi.nlm.nih.gov/Blast.cgi)[18]. Assays were designed so that the melting temperature of primers and probes were approximately 60°C and 70°C respectively. Amplicon size ranged between 51 and 151 nucleotides. Additionally, BLAST was used to select sequences for synthetic DNA controls (gBlocks™ (Integrated DNA Technologies, Coralville, IA, USA). Assay information can be found in Table S3.

**Table S3.** Oligonucleotide sequences for TaqMan® assays included on nanofluidic qPCR card (OpenArray® platform)

| Target | Forward (5'->3') | Reverse (5'->3') | Probe (5'->3') | Amp (bp) | Reference |
|---|---|---|---|---|---|
| *aph(3')-Ia* | ACCGAGGCAGTTCCATAGGA | ACCTTATTTTTGACGAGGGGAA | TGGTATCGGTCTGCGATT | 104 | 19 |
| *aac(6')-Ib-cr* | AACTTGCGAGCGATCCGA | TGGCGTGTTTGAACCATGTAC | TACGGTACCTTGCCTCT | 101 | 19 |
| $bla_{KPC}$ | GGCCGCCGTGCAATAC | GCCGCCCAACTCCTTCA | ACGCCGCCGCCAA | 60 | 11 |
| $bla_{NDM}$ | GACCGCCCAGATCCTCAA | CGCGACCGGCAGGTT | TGGATCAAGCAGGAGAT | 51 | 11 |
| $bla_{OXA-48}$ | CTTAAACGGGCGAACCAAGC | GTTCATCCTTAACCACGCCC | CTTGATCGCCCTCGATT | 93 | 12 |
| $bla_{IMP}$ | AGTGGTTTGGCTGCCTGAAA | TTTGGCCATGCTTTAATATTTGC | TCGGTGGTTGCTTTGTTA | 80-150 | 20 |
| $bla_{VIM}$ | AATGGTCTCATTGTCCGTGATG | TACAGCGTGGGGTGCGA | TGATGAGTTGCTTTTGATTG | 61 | 21 |
| $bla_{CMY-2}$ | GGCAAACAGTGGCAGGGTAT | AATGCGGCTTTATCCCTAACG | CACTTAGCCACCTATACG | 101 | 19 |
| $bla_{SHV}$ | TCCCATGATGAGCACCTTTAAA | TCCTGCTGGCGATAGTGGAT | TGCCGGTGACGAACA | 80-150 | 20 |
| $bla_{TEM}$ | GCATCTTACGGATGGCATGA | GTCCTCCGATCGTTGTCAGAA | TGCTGCCATAACCATGA | 100 | 22 |
| $bla_{CTX-M1}$ | CTGGGTGTGGCATTGATTAACA | CTCGCTGATTTAACAGATTCGGTT | ATGAGCGCTTTGCGATGT | 151 | 19 |
| $bla_{CTX-M9}$ | GCTTTATGCGCAGACGAATG | ATCACCGCGATAAAGCACCT | TCGATACCACAGATAATAC | 80-150 | 20 |
| *mcr-1* | GATCGCTGTCGTGCTCTTTG | ACCGCGCCCATGATTAATAG | CGATGCTACTGATCACC | 80-150 | 20 |
| *gyrA83L-ESh* | TGGTGACGTAATCGGTAAATACCA | CCGAAGTTACCCTGACCGTCT | ACCGCCAAGTCAC | 80-150 | 20 |
| *QnrB1* | GTAGCGCATATATCACGAATACCAATC | ATCTGAACCACTGAACGTCGC | TCGTGTTGGAAAAGTGT | 131 | 19 |
| *qnrS* | CGACGTGCTAACTTGCGTGA | GGCATTGTTGGAAACTTGCA | AGTTCATTGAACAGGGTGA | 118 | 21 |
| *vanA* | GCCGGAAAAAGGCTCTGAA | TTTTTTGCCGTTTCCTGTATCC | AGTTATAACCGTTCCCGCAGA | 65 | 23 |
| *mphA* | GTGCTGGCAATGCTCAAGAA | TGACCATCGCAGTCGAGTCT | AGCTCGTTGCCTATC | 80-150 | 20 |
| *cfr* | AATAAGTGCACCTGAGATGTATGGAG | CATATAATTGACCACAAGCAGCG | CGAAGGGCAGGTAGA | 109 | 24 |
| *mecA* | CGCAACGTTCAATTTAATTTTGTTAA | TGGTCTTTCTGCATTCCTGGA | ACGCTATGATCCCAATCT | 92 | 21 |

| | | | | | |
|---|---|---|---|---|---|
| *int1* | GGCAACTTTGGGCAGCA | CTGAAGCCAGGGCAGATCC | TTCGGTCTCCACGCAT | 147 | [24] |
| 16S rRNA | TGGAGCATGTGGTTTAATTCGA | TGCGGGACTTAACCCAACA | CTGACGACAACCATGCA | 137 | [11] |
| crAssphage | CAGAAGTACAAACTCCTAAAAAACGTAGAG | GATGACCAATAAACAAGCCATTAGC | ACGATTTACGTGATGTAAC | 126 | [17] |
| *sul1* | CCGTTGGCCTTCCTGTAAAG | TTGCCGATCGCGTGAAGT | CAGCGAGCCTTGC | 67 | [19] |
| *sul2* | CGGCTGCGCTTCGATT | CGCGCGCAGAAAGGATT | TGCTTCTGTCTGTTTCG | 60 | [19] |
| *TetA* | CGACGGCACAGGCTACATC | CCTGGACAACATTGCTTGCA | TGGCGTTCCCGATCA | 80-150 | [20] |
| *TetM* | CGGTTTCTCTTGGATACTTAAATCAATC | AGTCCGTCACATTCCAACCATAC | CACAGCCATAGCGTATC | 102 | [19] |
| *dfrA1* | TCGGGAATGGCCCTGATAT | GTCCAACCAACAGCCATTGG | CCATGGAGTGCCAAAG | 80-150 | [20] |

### S3.2. ARG Quantification using OpenArray® card

Samples were assayed in duplicate 33-nl reactions on a QuantStudio™ 12K Flex Real-Time PCR System which can perform duplicate qPCR measurements for 28 distinct targets. This resulted in four measurements of fluorescence (Crt values[25]) per target for each of the samples assayed. The OpenArray ® plate was manufactured with primers and probes tagged with carboxyfluorescein (FAM) at the 5 ́-end and Black Hole Quencher®-1(BHQ1) at the 3'-end. Prior to loading, 2.5μl of sample was combined with 2.5 μl of TaqMan® OpenArray® Real-Time PCR Master Mix (Applied Biosystems™). The QuantStudio 12K Flex Accufill System (Applied Biosystems™), was used to load the mixture on the OpenArray ® plate and samples were run on the QuantStudio 12K Flex Real-Time PCR System (Applied Biosystems) using standard thermocycling parameters: initial denaturation at 95°C for 3 min, followed by 40 cycles of denaturation at 95°C for 10s and annealing/extension at 60°C for 10 s. A 7-point standard curve, created through a serial dilution of pooled gBlocks™ (Integrated DNA Technologies, Coralville, IA, USA), was run on each OpenArray ® plate. The approximate concentrations for the points of standard curve were 100, 500, 1000, 10000, 100000, 1000000, and 10000000 gene copies per microlitre or as low as ~5.5 copies per 33nL well. The efficiency for each assay was determined through examination of the slope of individual standard curves using the formula:

$$E = -1 + 10^{\left(-1/k\right)}$$

Where *k* is the slope of the standard curve. Standard curves exceeded the requirements described in the Protocol for Evaluations of RT-qPCR Performance Characteristics: Technical Guidance (slope between -3.1 to -3.6 and $R^2$ of a minimum 0.98)[10]. Standard curve information for each assay can be found in Table S4. LOD/LOQ for individual assays included on the OpenArray® card have yet to be ascertained as the minimum number of samples required for this assessment have yet to be run. Negative controls were run in duplicate with each run on each OpenArray ® plate. Crt values produced via OpenArray® card were converted into concentrations using the formula:

$$copies = 10^{\left(\frac{Crt-}{k}\right)}$$

Where *b* is the intercept of the standard curve and *k* is the slope of standard curve. Finally, concentrations were converted into gene copies per litre using the following formula:

$$Amount\,\frac{gene\ copies}{L} = Rxn\ vol.\times\frac{Extract\ vol.}{Loaded\ vol.}\times\frac{1000nL}{1\mu L}\times Total\ Extract\ vol.\times\ Dilution\ factor\times\frac{1}{Sample\ vol.}\times\frac{1000mL}{1L}$$

$$Amount\left(\frac{gc}{L}\right) = 33nL\times\frac{2.5\mu L}{5\mu L}\times\frac{1000nL}{1\mu L}\times 50\mu L\times Dilution\ factor\times\frac{1}{sample\ volume}\times\frac{1000mL}{1L}$$

**Table S4.** Standard curve information for TaqMan® assays included on nanofluidic qPCR card (OpenArray ® platform)

| Antimicrobial Agent | Assay | Slope | Efficiency (%) | $R^2$ | Dynamic Range | |
|---|---|---|---|---|---|---|
| | | | | | Low | High |
| Aminoglycoside | *aph(3')-Ia* | -3.3725 | 97.93 | 0.995 | 7.1 | 709500 |
| | *aac(6')-Ib-cr* | -3.3523 | 98.75 | 0.997 | 7.8 | 782100 |
| Carbapenems | $bla_{KPC}$ | -3.2995 | 100.94 | 0.993 | 5.5 | 550000 |
| | $bla_{NDM}$ | -3.2326 | 103.86 | 0.992 | 5.5 | 550000 |
| | $bla_{OXA48}$ | -3.2428 | 103.41 | 0.994 | 5.5 | 550000 |
| | $bla_{IMP}$ | -3.3641 | 98.27 | 0.996 | 6.6 | 656700 |
| | $bla_{VIM}$ | - | - | - | - | - |
| Cephalosporins/extended spectrum ß-lactam (ESBL) antibiotics | $bla_{CMY-2}$ | -3.3888 | 97.28 | 0.997 | 7.6 | 760650 |
| | $bla_{SHV}$ | -3.3765 | 97.77 | 0.995 | 5.6 | 561000 |
| | $bla_{TEM}$ | -3.4186 | 96.12 | 0.997 | 7.6 | 760650 |
| | $bla_{CTX-M1}$ | -3.4284 | 95.74 | 0.995 | 5.9 | 592350 |
| | $bla_{CTX-M9}$ | -3.4357 | 95.46 | 0.991 | 16.2 | 1620300 |
| Colistin | *mcr-1* | -3.3382 | 99.32 | 0.993 | 10.6 | 1062600 |
| Fluoroquinolones | *gyrA83L-Esh* | -3.3932 | 97.11 | 0.996 | 14.2 | 1420650 |
| | *QnrB1* | -3.4146 | 96.27 | 0.996 | 14.1 | 1412400 |
| | *qnrS* | -3.4571 | 94.65 | 0.993 | 16.7 | 1666500 |
| Glycopeptides (vancomycin) | *vanA* | -3.4183 | 96.13 | 0.992 | 14.1 | 1412400 |
| Macrolides | *mphA* | -3.5024 | 92.98 | 0.993 | 16.9 | 1699500 |
| Methicillin | *mecA* | -3.4361 | 95.45 | 0.995 | 17.2 | 1716000 |
| Tetracycline | *tetA* | -3.3308 | 99.63 | 0.996 | 16.3 | 1630200 |
| | *tetM* | -3.3279 | 99.75 | 0.995 | 17.2 | 1716000 |
| Sulfonamides | *sul1* | -3.4037 | 96.69 | 0.997 | 16.9 | 1699500 |
| | *sul2* | -3.4285 | 95.74 | 0.994 | 17.2 | 1716000 |
| Trimethoprim | *dfrA1* | -3.414 | 96.29 | 0.998 | 17.2 | 1716000 |
| Associated with resistance to multiple therapeutics | *cfr* | -3.3858 | 97.40 | 0.997 | 14.1 | 1410750 |
| | *int1* | -3.4002 | 96.83 | 0.997 | 13.8 | 1376100 |
| Normalization/Control | 16S rRNA | -3.3307 | 99.64 | 0.996 | 12.5 | 1252350 |
| | crAssphage | -3.3472 | 98.96 | 0.997 | 10.6 | 1059300 |

### S4. Statistical methods

#### S4.1. Interrupted time series analysis for respiratory pathogens

Interrupted Time Series (ITS) was used to test if the 2024 NFL Draft in Detroit coincided with altered concentrations of respiratory pathogens in wastewater. Analyses were conducted independently for sites likely to be affected by the Draft (NIEA, OAK, JEFF and WEWTP; proximate to the Draft location) and control sites (CHWW, TB and LPCC; located outside of the Windsor-Detroit metropolitan area). Generalized linear mixed models (GLMMs) were fit for each respiratory virus (outcome variable) with an ITS analysis structure. An individual GLMM was run for each of the viral pathogens and false discovery rate (FDR) correction was applied to correct for multiple testing. Corrections were applied within each site cluster (control vs affected).

For SARS-CoV-2, a mean viral signal generated by averaging N1 and N2 concentrations was used. For all respiratory pathogens, points falling ± 3 standard deviations from the mean were treated as outliers and removed from the dataset prior to downstream analysis. Missing covariate data was filled via linear interpolation within each site.

Temporal structure within the data was addressed via a combination of natural splines (df=2-4, depending on pathogen), sine and cosine cyclical seasonal terms (52-week cycle) and to account for temporal dependence within sites an autoregressive structure was included (AR1). Finally, temperature, precipitation, flow, pH and the concentration of the other respiratory viruses were included in the models as covariates. PMMoV concentration was also included as a covariate. Prior to inclusion as covariates in the GLMMs, fixed effects were centered and scaled to have a mean of 0 and a standard deviation of 1. Data were collected between May 2023 and July 2024. Weekly averages were used for all data sets. The general model specification was:

$$\log(\mathbb{E}[Y_{it}]) = \beta_0 + \beta_1 \cdot event_{it} + \beta_2 \cdot posttime_{it} + \sum_{k=3}^{N} \beta_k \cdot X_{it}^{k} + u_{sitename(i)} + \varepsilon_{it},\ \varepsilon_{it} \sim AR(1)$$

Where $\mathbb{E}[Y_{it}]$ is the expected value of the response variable $Y_{it}$ at site *i* and time *t*, *event* is a binary variable that captures the immediate change in the outcome associated with the occurrence of the event, $posttime$ represents the change in outcome over time following the event, the summation $\sum_{k=3}^{N} \beta_k \cdot X_{it}^{k}$ represents additional covariates such as other virus concentrations, environmental variables, time and cyclical seasonal terms. A site specific random intercept $u_{(i)}$ accounts for baseline differences between sites. Models were fit using the *glmmTMB* package with a Tweedie distribution and a log link. These were selected to accommodate zero inflation, skew and over dispersion, all characteristic of wastewater data. Model fit was evaluated using the *DHARMa* package. Uniformity, dispersion, autocorrelation of residuals and outliers were all evaluated. Finally, likelihood ratio tests (LRTs) were used to compare models with and without the “event” parameter via AIC. To account for multiple testing, raw p-values were adjusted using the Benjamini-Hochberg false discovery rate (FDR) method. This was applied to adjust p-values across all models. Model predictions for all pathogens at both the control and affected sites were generated using the fitted models. Predictions with 95% confidence intervals generated from the standard error of predictions were plotted along with the observed measurements. Statistical methods and model fitting were largely the same for the control sites. One difference was the use of PMMoV normalized SARS-CoV-2 concentration rather than raw SARS-CoV-2 concentration at the control sites. This was done to allow for improved model fit. Tweaks to the temporal terms were also made to improve model fit.

**S4.2. Interrupted time series analysis for carbapenemase genes**

Data analysis was performed in R (version 4.5.1)[26] with data manipulation and visualization relying on the *tidyverse* suite of packages[27]. To determine if carbapenemase concentrations in wastewater were affected by the mass gathering associated with the 2024 NFL Draft an interrupted timeseries analysis was carried out using a generalized linear mixed modelling (GLMM) framework using the *glmmTMB* package [28] on two sets of sites. The first set was likely affected by Draft-associated transient population increases (n=4). The second set of sites was a control set of sites that were geographically removed from Detroit and are unlikely to have been influenced by Draft-related travel (n=3). Before modelling, missing covariate data was imputed and standardized via z-score. Individual carbapenemase concentrations were normalized using PMMoV (a human fecal indicator[13]) concentration and the influence of extreme outliers in ARG concentration was mitigated via Winsorization where values ±3 standard deviations from the mean were set to the 3 standard deviation level. A composite carbapenemase index was calculated by taking the average of the standardized normalized gene concentrations for $bla_{KPC}$, $bla_{NDM}$, $bla_{OXA\text{-}48}$, $bla_{VIM}$, and $bla_{VIM7}$. A small positive constant (1e-6) was added to this mean to ensure a strictly positive dataset. This composite measure was used for the initial ITS model. When composite normalized carbapenemase concentration was the response variable a GLMM with a Tweedie distribution and a log link function was used, allowing for the handling of zero-inflated continuous, right skewed data. Separate models were run for each carbapenemase gene measured to better understand the effect of the Draft on individual ARGs. Individual gene targets were modelled using a Gamma distribution and a log link. Both the composite measure and individual gene targets were modeled with the same modeling framework (save the different distribution). Within this framework, epidemiological week was used as the time variable, and a binary “dummy” variable was used to indicate the occurrence of the Draft. A second fixed term was used to investigate the possibility of a gradual change in ARG concentration following the Draft. Other fixed factors included wastewater temperature, wastewater pH, influent flow, and precipitation. The non-linear effects of flow and pH were modelled using natural splines (df=3). A random intercept was included to account for baseline variation between sites. Additionally, site-level autocorrelation was modeled using an autoregressive covariance term (AR1). Model fit and residual diagnostics including residual uniformity, dispersion, outlier frequency and temporal autocorrelation were assessed via simulations in the *DHARMa* package [29]. Additionally, autocorrelation function (ACF) plots were generated to visually check if the AR term adequately addressed within site autocorrelation. For each response variable, a reduced model (excluding intervention effects) was compared to the full model via a likelihood ratio test, and counterfactual predictions were generated by setting the dummy variables to 0, allowing estimation of carbapenemase concentrations in the absence of the Draft. For ARGs with low detection frequencies ($bla_{VIM}$ and $bla_{VIM7}$ at the control sites) GLMMs would not converge due to data sparsity and thus are not reported. The FDR method (Benjamini-Hochberg procedure) was used to adjust all model derived p-values for gene-specific /individual ARG models to account for multiple comparisons within each cluster of sites (control and affected). Statistical significance was evaluated at the $\alpha=0.05$ level.

#### S4.3. Multivariate resistome characterization

The OpenArray ® was used to analyze a select set of samples collected from wastewater treatment facilities during the period surrounding the 2024 NFL Draft. Gene concentrations were normalized to the concentration of *crAssphage*, a bacteriophage associated with human feces and a known human fecal indicator[17,30]. Normalized concentrations were transformed via fourth-root transformation. The data were split into 3 uneven groups (“before”, “during” and “after”) relative to the timing of the NFL-Draft. Characterization of resistome structure was carried out using non-metric multidimensional scaling (NMDS), using Bray-Curtis dissimilarity matrices. A Permutational Multivariate Analysis of Variance (PERMANOVA) was then run on the data using the adonis2 function[31] to explore sources of variation. A marginal PERMANOVA was employed to partition variance to each factor and distance-based redundancy analysis was used for visualization of variance partitioning. A Permutational Analysis of Multivariate Dispersions (PERMDISP) was used to confirm that any observed differences between groups arose from differences in resistome rather than from within group variance. A principal component analysis (PCA) was run to identify which ARGs were the main drivers of variability between sites and groups. The marginal PERMANOVA and the PCA were run again on data set with 16S rRNA data to ensure the robustness of results. Finally, the JEFF interceptor was analyzed in isolation from the other sites as it drains the NFL Draft location and had the most complete sampling regime. A Kruskal-Wallis test was used for this location to determine if significant differences could be observed between the “before”, “during” and “after” groupings for any of the assayed gene-targets. The p-values were adjusted for multiple comparisons using the Benjamini-Hochberg method. All analyses were performed in R (4.5.1) [26].

**S4.4. Detection of carbapenemase genes in environmental samples collected in receiving waters surrounding the 2024 NFL Draft.**

RT-qPCR was used to assess the concentration of PMMoV, $bla_{KPC}$, $bla_{NDM}$, $bla_{OXA\text{-}48}$, $bla_{VIM}$ and $bla_{VIM7}$ in river samples collected both before and after the Draft in a six site transect along the Detroit River. A major rainfall event coincided with the NFL Draft weekend, and combined sewage overflow (CSO) events were recorded on both the Canadian and American side of the border[32]. Detections were sparse for all assayed genes save PMMoV and $bla_{KPC}$, so a detection frequency analysis was carried out to assess if the Draft and CSO event were associated with altered detection probabilities. Detection frequencies were calculated both before and after the Draft and Fisher’s exact test was used to compare detection frequencies for each gene. The p-values were adjusted for multiple comparisons using the Benjamini-Hochberg method. In addition, a paired Wilcoxon signed-rank test was employed to test if a change in the median PMMoV normalized concentration of $bla_{KPC}$ could be detected between the pre- and post- Draft periods. All analyses were performed in R (4.5.1) [26].

**S4.5. Measurement of ARGs in the Detroit River using a Open Array nanofluidic RT-qPCR card**

The concentration of a wide array of ARGs was assessed in river samples collected from six sites in the Detroit River using an Open Array nanofluidic RT-qPCR card. Samples were collected both before and after the 2024 NFL Draft weekend. The Draft occurred in Detroit and coincided with a major rainfall event which led to combined sewer overflow (CSO) events in the region draining into the Detroit River. Prior to quantitative analyses, ARG concentrations were normalized to *16S rRNA* concentration, and logarithmically transformed. Samples where only one technical replicate amplified were treated as non-detects. Samples were categorized as either “before” or “after” based on the timing of their collection in relation to the 2024 NFL Draft. Detection frequencies (based on binary presence/absence) were calculated for all the ARGs assayed on the Open Array nanofluidic RT-qPCR card. Logistic regression was used to test if detection frequency for ARGs changed over the Draft weekend. ARGs that were detected in all samples either before or after the NFL Draft and ARGs that were not detected at all were not included in the detection frequency analysis to avoid “complete separation” during logistic regression and allow model convergence. All gene targets were included in descriptive statistics and quantitative abundance analyses.

16s rRNA was detected in all samples collected and was used as a normalizing gene, thus it was not included in the detection frequency analysis. Likewise, *int1* was found in all samples and thus its detection frequency did not change over the Draft weekend. Additionally, $bla_{NDM}$, *mcr1*, *cfr* and *vana* were not detected in any samples. *Sul1*, *sul2* and *tetm* were excluded from modeling because detection frequency increased to 100% following the Draft. $bla_{OXA\text{-}48}$ was one of the few targets that was not detected in any of the samples in the post-Draft grouping, leading to its exclusion from the logistic regression. Finally, *ctx-m9*, *gyrA83L-ESh*, and *mecA* were all excluded from modelling since they were not detected in the pre-Draft group. Thus, 13 gene targets were included in the logistic regression detection frequency analysis.

A generalized linear mixed model (GLMM) with a binomial distribution and a logit link function was fitted to the data to inspect the impact of the Draft on the detection probability of ARGs in the environment. The structure of the model was as follows:

$$Detection \sim group \times gene + (1 + group|site)$$

Fixed effects included *gene* (ARG identity), *group* (sample collection timing in relation to the Draft ("before" vs "after)) and the interaction between *gene* and *group* to allow the effect of the Draft to differ between genes. Random intercepts and random slopes for group were included at the site level to account for differences in baseline detection probability and site-specific responses to the Draft. Following model fitting, estimated marginal means were calculated and pair-wise comparisons were conducted between *groups* "before" and "after" for each ARG.

A second GLMM with a logit link and binomial distribution was fitted with site and group as fixed effects and a random intercept for gene target. The goal of this model was to directly test the effect of sampling site on detection probability. The model was specified as:

$$Detection \sim group + site + (1|target)$$

Model fit for each GLMM was assessed using simulated residuals in the *DHARMa* package. Finally, paired Wilcoxon signed-rank tests were used to determine if ARG concentration differed between the "before" and "after" groupings. A sensitivity analysis was conducted to determine if ARG saturation at the point source of contamination was masking temporal trends in ARG concentration. Pairings were based on sampling site and ARG identity and p-values were adjusted to account for multiple comparisons using the Benajmini-Hochberg method to controlFDR.

#### S4.6. Comparison of nanofluidic qPCR card using TaqMan® assays in the OpenArray ® platform and RT-qPCR

OpenArray ® assays were validated via comparison to established RT-qPCR assays on a subset of wastewater and environmental samples that were analyzed by both methods (n=42). Comparison was limited to assays for $bla_{KPC}$, $bla_{NDM}$, and $bla_{OXA\text{-}48}$ as they were shared between the OpenArray ® and the multiplexed RT-qPCR assays. Data analysis was performed in R (version 4.5.1)[26] using *mcr, boot,* and *DescTools* [33–35]. Concentrations were subjected to logarithmic transformation at the technical replicate level to stabilize variance and approximate normality. Measurements were paired by sample and by gene and the average value across technical replicates (n=2 for OpenArray ®, n=3 for RT-qPCR) was used for further analysis. Samples were defined as positive if the mean concentration was greater than zero. Wastewater and environmental samples had different concentration distributions and thus environmental samples were analyzed for detection level agreement while wastewater samples were analyzed for both quantitative agreement and detection level agreement. For environmental samples, binary detection was tested using Cohen's Kappa (κ) and a comparison of detection frequencies between platforms. Tests were performed on the full data set and individually on wastewater and environmental samples. Outliers were classified as sample pairs where the log-transformed fold change was greater than 1.5× *IQR* from the median. Normality of transformed data was assessed on a per target basis using Shapiro-Wilk tests and non-parametric tests were prioritized as a result. Bland-Altman analysis was used to assess mean bias and 95% limits of agreement (LoA) on the log scale. The ratio of raw concentrations between the OpenArray ® and RT-qPCR results was calculated to determine relative recovery rates (fold change). Correlations (Spearman and Pearson) were also calculated for each target. A non-parametric measure of proportional and constant bias, Passing-Bablok regression[36], was implemented as a primary measure of method comparison. This was followed by Deming regression, a variance weighted regression technique that accounts for measurement error in both means being analyzed[37]. Deming regression was performed with bootstrapping (1000 iterations) and was used as validation of Passing-Bablok results. Total agreement was quantified using concordance correlation coefficients (CCC) with bootstrapped (1000 iterations) 95% confidence intervals.

### S5. Supplemental results

#### S5.1. Interrupted time series analysis for respiratory pathogens

#### Affected sites

The GLMM (n = 228; AIC = 4469.0, Table S5) suggests that SARS-CoV-2 concentrations did not increase significantly in wastewater following the 2024 NFL Draft ($\beta$=0.092, SE=0.277, p = 0.811). Evidence of a decreasing trend in SARS-COV-2 concentration post NFL-Draft did not survive FDR correction ($\beta$=–0.317, SE=0.177, p = 0.168). Visual inspection of the data corroborated model results (Figure S1). PMMoV was a strong positive predictor of SARS-CoV-2 concentration ($\beta$ = 0.379; SE=0.065, p<0.001), while average wastewater flow was inversely associated ($\beta$=–0.288, SE=0.115, p = 0.035). Temporal terms, seasonal and long-term trends (splines, sine/cosine terms) significantly improved model fit. *DHARMa* simulated residual diagnostics generally indicated adequate model fit. However, a quantile test flagged a minor departure (p = 0.00042) that is small in magnitude (Table S6). Including *event* and *posttime* did not significantly improve model fit (LRT $\chi^2$=3.9, *p* = 0.142; ΔAIC≈0.1, Table S7).). The model provided no evidence of an increase in SARS-CoV-2 concentration resulting from the gathering of individuals for the 2024 NFL Draft. SARS-CoV-2 concentrations were primarily driven by seasonal variations and wastewater concentration, as indicated by significant PMMoV and flow covariates (Table S8)

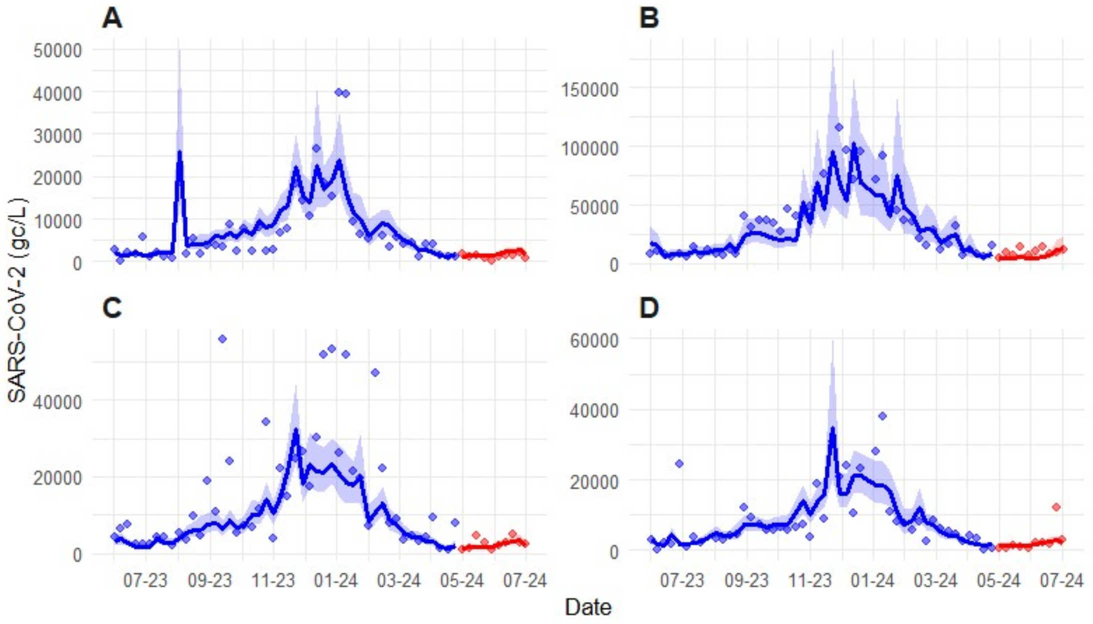


**Figure S1.** Longitudinal wastewater surveillance data for SARS-CoV-2 at the four draft proximal sites (**A -JEFF**, **B-WEWTP, C-NIEA, D-OAK**) both before (blue), and after (red) the 2024 NFL draft. Points represent mean SARS-CoV-2 concentrations in gene copies per litre. Solid lines and shaded areas denote predicted mean concentrations and 95% confidence intervals.

The GLMM (n = 226; AIC = 3072.9; Table S5) run to explain variability in IAV signal suggests that the NFL Draft did not significantly influence wastewater concentrations of IAV (β=-0.312, SE=0.619, p = 0.745 ). Additionally, the posttime parameter was non-significant (β=0.049, SE=0.324, p = 0.912) indicating no post event rise in IAV. Visualization of the data was consistent with model results (Figure S2). Non-significant NFL Draft related parameter estimates were corroborated by LRT which suggested that including *event* and *posttime* may have worsened model fit (LRT $\chi^2$=0.255, p = 0.142; ΔAIC≈-3.7; Table S7). Temporal terms proved very important in explaining IAV variability, as is expected for a seasonal respiratory illness. Finally, precipitation (β=0.317, SE=0.079, p<0.001) and flow (β=-0.467, SE=0.160, p = 0.012) were found to be significant predictors of IAV concentration in wastewater (Table S8). Increased flow was associated with decreased IAV concentrations, consistent with dilution, but precipitation was a positive predictor of IAV levels. Diagnostic tests relying on *DHARMa* simulated residuals showed no evidence of model misfit (Table S6).

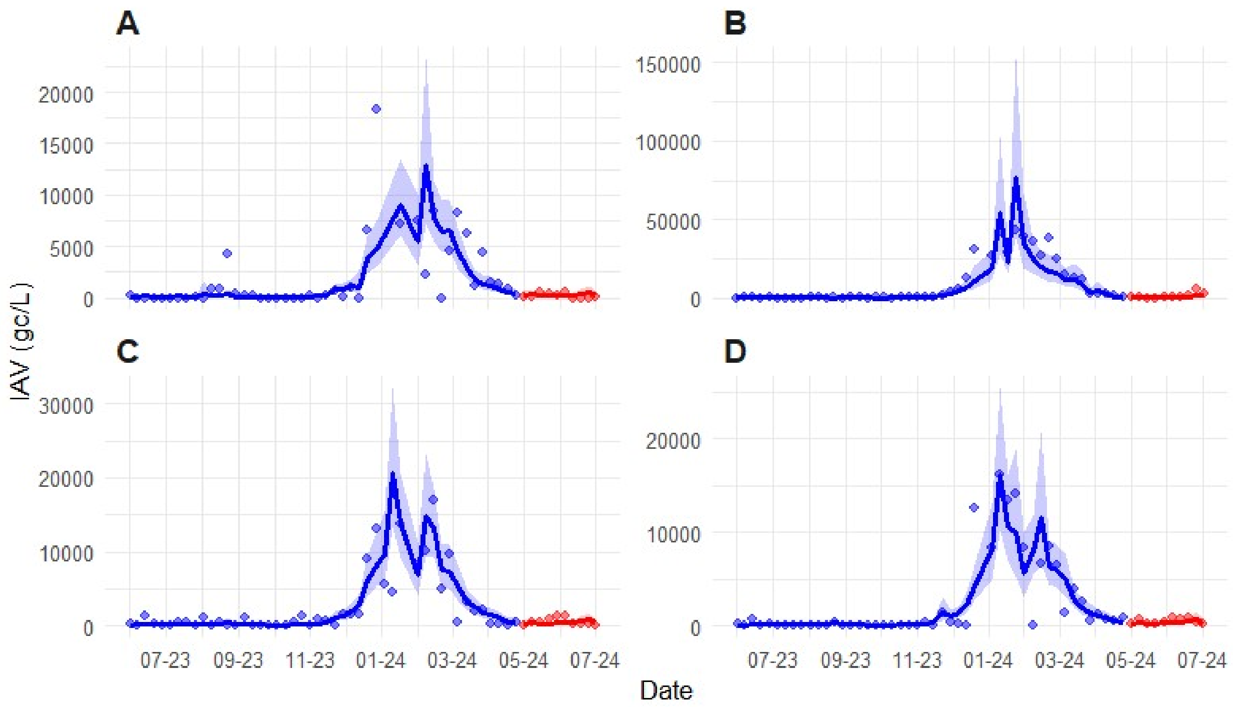


**Figure S2.** Longitudinal wastewater surveillance data for IBV at the four draft proximal sites (**A -JEFF** , **B-WEWTP, C-NIEA, D-OAK**) both before (blue), and after (red) the 2024 NFL draft. Points represent IBV concentrations in gene copies per litre. Solid lines and shaded areas denote predicted mean concentrations and 95% confidence intervals, respectively.

Modelling and visualization for IBV (n = 224; AIC = 1556.9; Table S5) indicates that the NFL Draft did not cause an immediate increase in IBV concentration (β=-0.881, SE=0.597, p = 0.244). However, the posttime parameter was significant (β=-0.356, SE=0.143, p = 0.035) suggesting a decline in IBV concentration following the Draft (Table S8, Figure 2, main manuscript). This is consistent with the cessation of the IBV season and is not likely linked to the event. LRT supported the inclusion of *event* and *posttime* in the model as they improved model fit (LRT $\chi^2$=9.507, p = 0.009; ΔAIC≈5.438; Table S7). Precipitation was again positively associated with pathogen concentration (β=0.345, SE=0.138, p = 0.035) while flow was strong negative predictor (β=-0.924, SE=0.284, p = 0.005; Table S8). Finally, IBV was found to be positively associated with IAV concentration (β=0.814, SE=0.244, p = 0.004). Tests of *DHARMa* simulated residuals were acceptable (Table S6).

The GLMM run for RSV (n = 225; AIC = 2609.9, Table S5) indicated that the NFL Draft did not influence RSV concentration in wastewater (β=-0.240, SE=0.586, p = 0.801). Additionally, the posttime parameter was non-significant (β=0.014, SE=0.199, p = 0.959) and the inclusion of *event* and *posttime* did not significantly improve model fit (LRT $\chi^2$=0.171, *p* = 0.918; ΔAIC≈-3.83; Table S7). Modelling results were supported by visualization of the data (Figure S3). PMMoV was a positive predictor of RSV concentration (β= 0.455, SE= 0. 141, p = 0.005) while IAV was a negative predictor (β= -0.318, SE= 0.128 p = 0.035; Table S8). The negative association of IAV with RSV may rise from non-overlapping respiratory season or may be indicative of virus-virus interactions at the population level [38–40]. Again, residual diagnostics indicate no issues with model fit (Table S6).

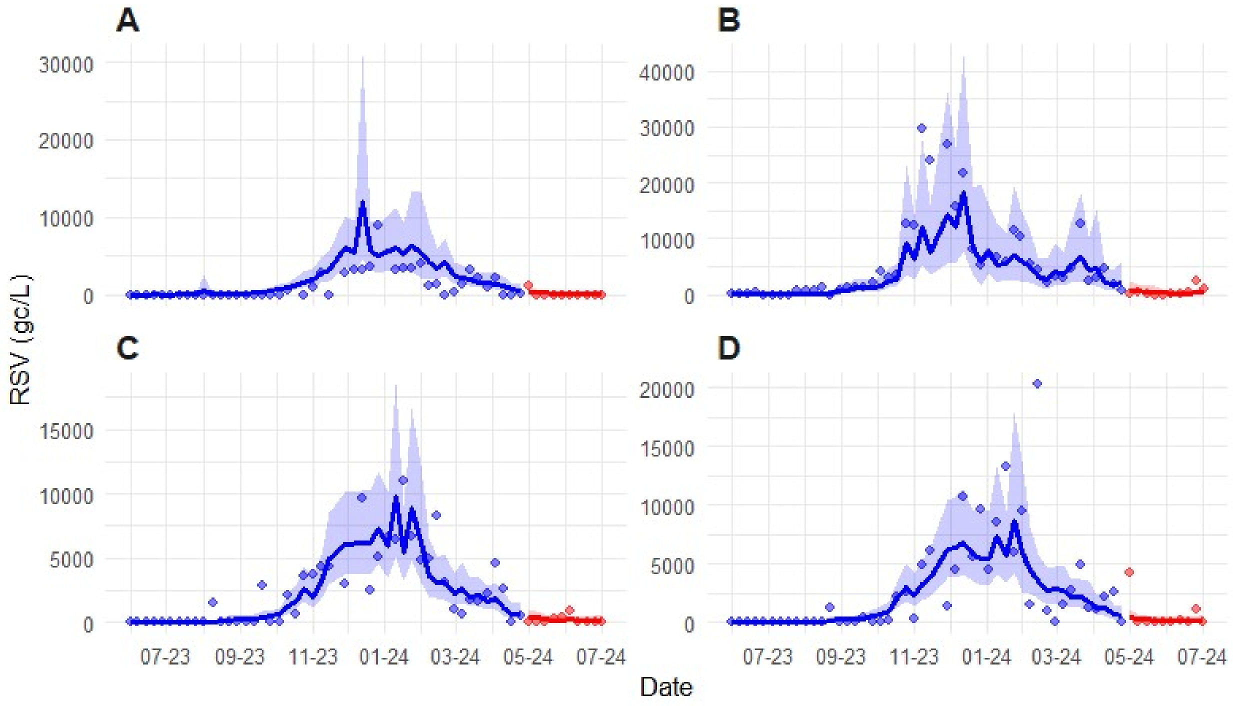


**Figure S3.** Longitudinal wastewater surveillance data for RSV at the four draft proximal sites (**A -JEFF** , **B-WEWTP, C-NIEA, D-OAK**) both before (blue), and after (red) the 2024 NFL draft. Points represent RSV concentrations in gene copies per litre. Solid lines and shaded areas denote predicted mean concentrations and 95% confidence intervals.

Modelling for all four pathogens supported strong temporal and seasonal variations in wastewater concentrations consistent with seasonal respiratory infections (Table S8). PMMoV concentrations and wastewater flow measurements were consistent predictors of concentration variability with increased PMMoV associated with higher pathogen concentrations and flow associated with lower pathogen concentrations, observations in line with the expected results of dilution. Precipitation was an important predictor of influenza viruses (A and B) but was found to be of less importance for predicting SARS-CoV-2 and RSV (Table S8). Finally, interactions between viruses were noted, with IAV positively predicting IBV but being negatively associated with RSV. This observation is likely due to differences in timing between IAV and RSV seasons but could also hint at virus-virus interactions.

**Table S5.** Summary of GLMMs for respiratory virus ITS analysis at affected sites using Tweedie distribution, log link function, and an AR(1) correlation structure. Random intercept variance ($\sigma^2$) represents site level heterogeneity. The AR(1) variance and correlation describe within site temporal dependency of viral concentrations

| Respiratory Virus | No. observations | AIC | BIC | logLik | Site Intercept ($\sigma^2$) | AR(1) ($\sigma^2$) | AR(1) $\rho$ | Tweedie Disp |
|---|---|---|---|---|---|---|---|---|
| SARS-CoV-2 | 228 | 4469.0 | 4541.1 | −2213.5 | 0.0312 | 0.3026 | 0.03 | 65.9 |
| IAV | 226 | 3072.9 | 3148.2 | −1514.5 | <0.0001 | 0.1840 | 0.47 | 40.6 |
| IBV | 224 | 1556.9 | 1618.3 | −760.5 | 0.0518 | 0.3355 | 0.92 | 48.6 |
| RSV | 225 | 2609.9 | 2678.2 | −1284.9 | <0.0001 | 0.1672 | 0.99 | 105.0 |

**Table S6.** Residual diagnostics for respiratory virus ITS GLMMs run on affected site data performed using simulated residuals (*DHARMa* package)

| Respiratory Virus | KS | Dispersion | Quantile | Outliers (n) |
|---|---|---|---|---|
| SARS-CoV-2 | 0.257 | 0.178 | <0.001 | 0.077 (2) |
| IAV | 0.961 | 0.698 | 0.211 | 0.364 (1) |
| IBV | 0.381 | 0.302 | 0.222 | 1.000 (0) |
| RSV | 0.517 | 0.846 | 0.656 | 1.000 (0) |

**Table S7.** Results of Likelihood Ratio Tests comparing full respiratory virus ITS GLMMs (including “event” and “posttime” terms) with reduced models for the affected site cluster. A significant $\chi^2$ value indicates that the inclusion of event related variables significantly improved model fit, suggesting a detectable impact on viral loads.

| Respiratory Virus | $\chi^2$ | *Degrees of freedom* | p-value | Interpretation |
|---|---|---|---|---|
| SARS-CoV-2 | 3.904 | 2 | 0.142 | Non-significant |
| IAV | 0.255 | 2 | 0.880 | Non-significant |
| IBV | 9.507 | 2 | 0.0086 | Significant |
| RSV | 0.171 | 2 | 0.918 | Non-significant |

**Table S8.** Fixed effects coefficients for respiratory virus ITS GLMM predictors remaining significant following Benjamini-Hochberg FDR correction (p<0.05) for the affected site cluster. Estimates are reported on the link scale with 95% Confidence Intervals (CI).

| Model/Respiratory Virus | Predictor | Estimate | 95% CI | Std. Effect | FDR p-value |
|---|---|---|---|---|---|
| SARS-CoV-2 | ns(time_index)1 | 37.264 | 17.994–56.533 | 0.00192 | $8.58\times10^{-4}$ |
| SARS-CoV-2 | ns(time_index)2 | −31.976 | −48.980–−14.973 | −0.00165 | $1.18\times10^{-3}$ |
| SARS-CoV-2 | ns(time_index)3 | 17.143 | 6.696–27.590 | 0.000883 | $4.63\times10^{-3}$ |
| SARS-CoV-2 | sin52 | 15.126 | 6.997–23.256 | 0.000779 | $1.26\times10^{-3}$ |
| SARS-CoV-2 | cos52 | −9.744 | −14.629–−4.859 | −0.000502 | $5.85\times10^{-4}$ |
| SARS-CoV-2 | avg_PMMoV_s | 0.379 | 0.252–0.506 | $1.95\times10^{-5}$ | $2.75\times10^{-7}$ |
| SARS-CoV-2 | avg_flow_s | −0.288 | −0.513–−0.062 | $-1.48\times10^{-5}$ | $3.52\times10^{-2}$ |

| | | | | | |
|---|---|---|---|---|---|
| IAV | ns(time_index)2 | −45.902 | −66.995–−24.809 | −0.00602 | $1.90\times10^{-4}$ |
| IAV | ns(time_index)3 | 43.367 | 24.964–61.770 | 0.00569 | $4.57\times10^{-5}$ |
| IAV | ns(time_index)4 | −9.905 | −14.545–−5.266 | −0.00130 | $2.32\times10^{-4}$ |
| IAV | sin52 | −24.936 | −35.536–−14.336 | −0.00327 | $4.57\times10^{-5}$ |
| IAV | cos52 | 11.076 | 6.492–15.659 | 0.00145 | $4.13\times10^{-5}$ |
| IAV | PRCP_s | 0.317 | 0.162–0.472 | $4.16\times10^{-5}$ | $4.42\times10^{-4}$ |
| IAV | avg_flow_s | −0.467 | −0.781–−0.154 | $-6.13\times10^{-5}$ | $1.18\times10^{-2}$ |
| IBV | ns(time_index)2 | 6.398 | 3.782–9.015 | 0.00285 | $4.13\times10^{-5}$ |
| IBV | posttime | −0.356 | −0.638–−0.075 | −0.000159 | $3.52\times10^{-2}$ |
| IBV | avg_IAV_s | 0.814 | 0.336–1.292 | 0.000363 | $3.67\times10^{-3}$ |
| IBV | PRCP_s | 0.345 | 0.074–0.616 | 0.000154 | $3.52\times10^{-2}$ |
| IBV | avg_flow_s | −0.924 | −1.480–−0.368 | −0.000412 | $4.55\times10^{-3}$ |
| RSV | avg_IAV_s | −0.318 | −0.569–−0.067 | $-7.01\times10^{-5}$ | $3.52\times10^{-2}$ |
| RSV | avg_PMMoV_s | 0.455 | 0.179–0.730 | 0.000100 | $4.62\times10^{-3}$ |

### Control sites

An additional set of parallel models were fitted to data collected from sites not likely to be influenced by the NFL Draft. The parallel set of models were run to control for seasonal variation in pathogen concentration. The GLMM for SARS-CoV-2 differed in that a gamma GLMM with a log link fit the data better than a GLMM with a Tweedie distribution. Additionally, PMMoV normalized SARS-CoV-2 was the response variable (Table S9). The GLMM for SARS-CoV-2 did not support the notion that the NFL-Draft increased pathogen concentrations at the control sites as neither *event* nor *posttime* were significant. However, results of LRT indicated that the inclusion of these dummy variables slightly improved model fit (LRT $\chi^2$=0.623, $p$ = 0.044; ΔAIC≈2.228; Table S10). The only parameter to remain significant following p-value adjustment was flow, which was found to be a positive predictor of normalized SARS-CoV-2 concentration (β=9.364, SE= 1.869, p <0.001; Table S11). Given the inverse relationship between PMMoV concentration and flow, it might be expected that a normalized response variable be positively related with flow. This result seems to indicate that PMMoV may be more sensitive to dilution than SARS-CoV-2. The IAV GLMM found no significant effects for event or posttime and LRT analysis supported the notion that *event* and *posttime* did not contribute to model fit (LRT $\chi^2$=3.053, $p$ = 0.217; ΔAIC≈0.946; Table S10). Similarly, IBV was found to have no significant predictors and LRT indicated that *event* and *posttime* did not contribute to the model, slightly reducing AIC (LRT $\chi^2$=0.605, $p$ = 0.739; ΔAIC≈3.395; Table S10). The GLMM fit to the RSV data followed the same trend with no significant predictors following FDR correction. LRT showed that inclusion of event variables did not improve model fit, leading to an increased AIC (LRT $\chi^2$=830, $p$ = 0.661; ΔAIC≈3.170; Table S10). Seasonal harmonics and autoregressive terms were consistent in explaining variance via temporal structure (Table S11). Diagnostics run on *DHARMa* simulated residuals indicate acceptable model fit for all the models run on the control data set (Table S12). No statistically robust evidence of an increase in pathogen presence related to the timing of the NFL Draft could be observed at the control sites. Thus, results of control site analysis do not invalidate the analysis run on affected sites. If findings had suggested a parallel increase in pathogen concentration at the control and affected sites, it would invalidate analysis and undermine the notion that the mass gathering associated with the NFL Draft caused an increase in pathogen presence in wastewater (mirroring a community wide increase in prevalence). Visualization of pathogen trajectories at control sites are displayed in Figures S4-S6.

**Table S9.** Summary of GLMMs for ITS analysis of respiratory viruses at control sites using Tweedie distribution, log link function, and an AR(1) correlation structure. Random intercept variance (σ2) represents site level heterogeneity. The AR(1) variance and correlation describe within site temporal dependency of viral concentrations.

| Respiratory Virus | No. observations | AIC | BIC | logLik | Site Intercept ($\sigma^2$) | AR(1) ($\sigma^2$) | AR(1) $\rho$ | Tweedie/ gamma Disp |
|---|---|---|---|---|---|---|---|---|
| SARS-CoV-2 | 169 | -2219.2 | -2169.1 | 1125.6 | $1.68 \times 10^{-9}$ | 0.1391 | 0.52 | 0.185 |
| IAV | 170 | 2402.5 | 2465.2 | -1181.3 | $1.04 \times 10^{-5}$ | 0.6584 | 0.80 | 22.6 |
| IBV | 169 | 1563.1 | 1628.8 | -760.6 | 0.0043 | 0.9205 | 0.53 | 51.0 |
| RSV | 171 | 2108.5 | 2174.5 | -1033.3 | $5.61 \times 10^{-8}$ | 0.3304 | 0.71 | 70.2 |

**Table S10.** Results of Likelihood Ratio Tests comparing full respiratory virus ITS GLMMs (including "event" and "posttime" terms) with reduced models for the control site cluster. A significant χ2 value indicates that the inclusion of event related variables significantly improved model fit, suggesting a detectable impact on viral loads.

| Respiratory Virus | $\chi^2$ | *Degrees of freedom* | p-value | interpretation |
|---|---|---|---|---|
| SARS-CoV-2 | 6.227 | 2 | 0.044 | Significant |
| IAV | 3.053 | 2 | 0.217 | Non-significant |
| IBV | 0.605 | 2 | 0.739 | Non-significant |
| RSV | 0.830 | 2 | 0.661 | Non-significant |

**Table S11.** Fixed effects coefficients for predictors remaining significant following Benjamini-Hochberg FDR correction (p<0.05) for the control site cluster for respiratory virus ITS using a GLMM. Estimates are reported on the link scale with 95% Confidence Intervals (CI).

| Model/Respiratory Virus | Predictor | Estimate | 95% CI | Std. Effect | FDR p-value |
|---|---|---|---|---|---|
| SARS-CoV-2 | sin52 | -0.513 | -0.743 to -0.282 | NA | $1.83 \times 10^{-4}$ |
| SARS-CoV-2 | cos52 | -1.352 | -1.588 to -1.117 | NA | $1.21 \times 10^{-27}$ |
| SARS-CoV-2 | avg_flow_s | 9.364 | 5.701-13.027 | NA | $9.94 \times 10^{-6}$ |
| IAV | sin52 | -3.047 | -3.920 to -2.174 | $2.77 \times 10^{-4}$ | $2.18 \times 10^{-10}$ |
| IBV | None | - | - | - | - |
| RSV | None | - | - | - | - |

**Table S12.** Residual diagnostics for ITS GLMMs run on control site respiratory virus data performed using simulated residuals (DHARMa package).

| Respiratory Virus | KS | Dispersion | Quantile | Outliers (n) |
|---|---|---|---|---|
| SARS-CoV-2 | 0.494 | 0.650 | 0.068 | 0.287 (1) |
| IAV | 0.753 | 0.932 | 0.170 | 1.000 (0) |
| IBV | 0.714 | 0.194 | 0.155 | 1.000 (0) |
| RSV | 0.752 | 0.662 | 0.163 | 0.290 (1) |

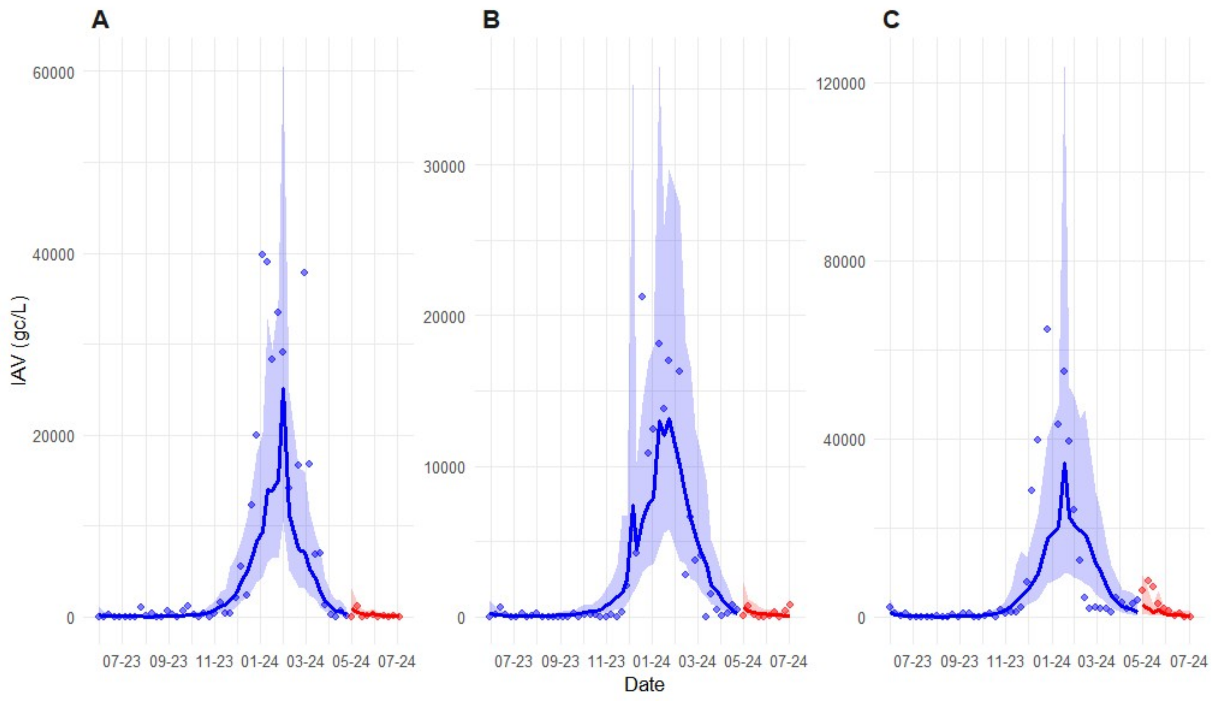


**Figure S4.** Longitudinal wastewater surveillance data for IAV at the three control sites (**A -CHWW**, **B-LPCC, C-TB**) both before (blue), and after (red) the 2024 NFL draft. Points represent mean IAV gene concentrations. Solid lines and shaded areas denote predicted mean concentrations and 95% confidence intervals, respectively.

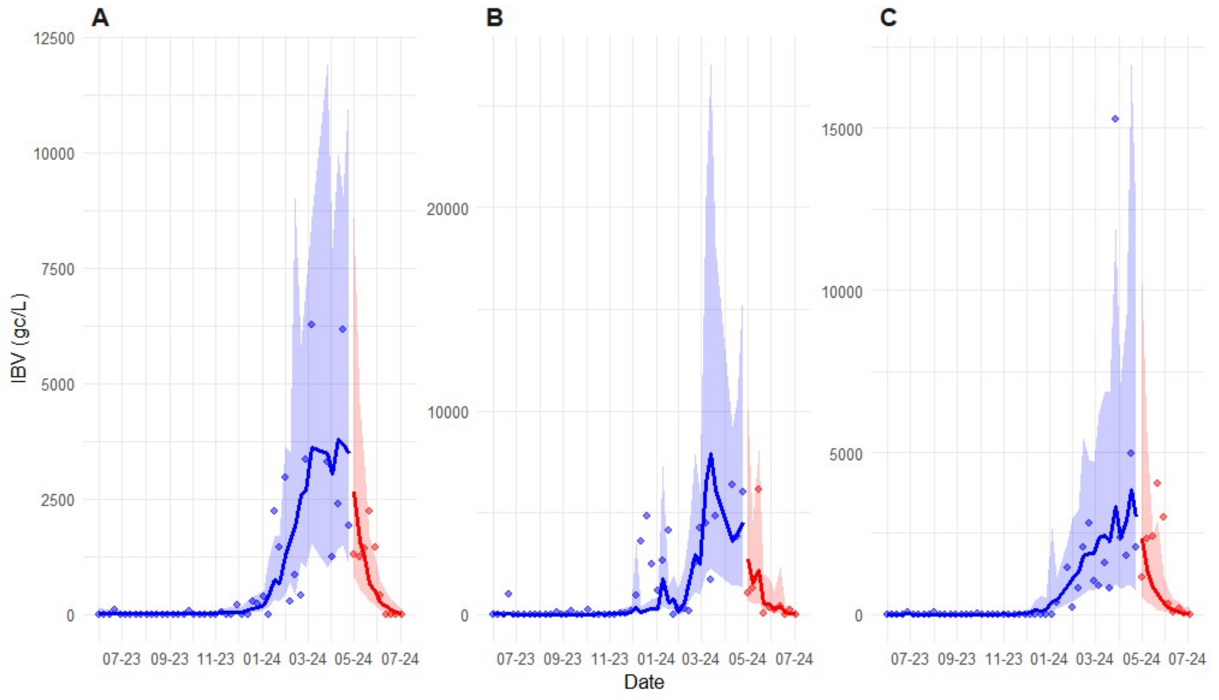


**Figure S5.** Longitudinal wastewater surveillance data for IBV at the three control sites (**A -CHWW**, **B-LPCC, C-TB**) both before (blue), and after (red) the 2024 NFL draft. Points represent mean IBV gene concentrations. Solid lines and shaded areas denote predicted mean concentrations and 95% confidence intervals, respectively.

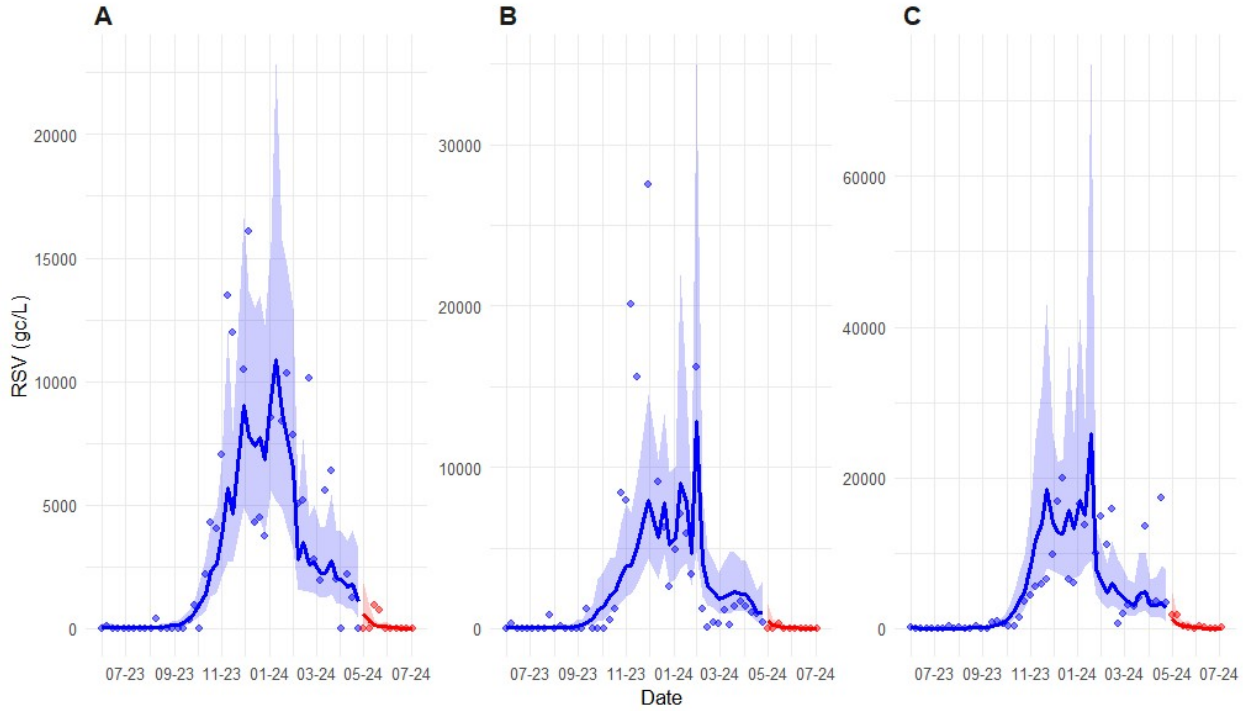


**Figure S6.** Longitudinal wastewater surveillance data for RSV at the three control sites (**A -CHWW**, **B-LPCC**, **C-TB**) both before (blue), and after (red) the 2024 NFL draft. Points represent mean RSV gene concentrations. Solid lines and shaded areas denote predicted mean concentrations and 95% confidence intervals, respectively.

### S5.2. Results interrupted time series analysis for carbapenemase genes

#### Affected sites

At the four Draft-proximal sites, the ITS specified GLMM (Table S13) showed the composite measure of carbapenemase concentration increased in a statistically significant manner concurrent with the 2024 NFL Draft. The binary variable indicating the occurrence of the Draft was positively associated with the composite measure of carbapenemase concentration (β=0.625, SE=0.290, p=0.031), corresponding to an approximate 87% increase in combined ARG burden immediately following the Draft (Table S14). The post-event temporal trend variable was non-significant (p=0.96), indicating the influence of the event was abrupt with no gradual temporal shifts in ARG concentration occurring following the event. Flow was the only significant environmental covariate and showed a significant positive non-linear influence on composite carbapenemase concentration (p<0.001; Table S14). LRT comparing the full model with a reduced model excluding both the intervention terms shows a marginal improvement in model fit with the inclusion of both Draft effect terms ($\chi^2$ =5.60, *df* = 2, p=0.061; Table S15). LRT comparing the reduced model to a model with only the step change term shows that inclusion of the event variable significantly improves model fit ($\chi^2$ =5.60, *df* = 1, p=0.018; Table S15). Simulation based tests of residuals indicate adequate model fit with no evidence of overdispersion, misspecification, non-uniformity, or autocorrelation (Table S16). Visualization of counterfactual predictions supports the influence of the Draft on increasing carbapenemase concentration in wastewater as they consistently underestimated concentrations in comparison to observed data (Figure 4, main manuscript).

Modelling of individual carbapenemase ARGs indicates that the increase in ARG composite index was primarily driven by $bla_{OXA\text{-}48}$ which exhibited the largest event associated increase. The occurrence of the Draft showed a strong positive association with PMMoV normalized $bla_{OXA\text{-}48}$ concentration (β=1.38, SE=0.374, p=0.006; Table S14), indicating an approximate 400% increase in concentration following the mass gathering associated with the 2024 NFL Draft. Once again, the effect of the Draft was stepwise with the intervention parameter modeling gradual changes in $bla_{OXA\text{-}48}$ concentration not showing statistical significance. Flow was the only significant covariate, showing a strong positive association with $bla_{OXA\text{-}48}$ signal (p<0.001; Table S14). LRT indicated that the full model (with the event associated covariates included), had a substantially better fit than the reduced model, indicating the Draft parameter was significant in explaining variability in $bla_{OXA\text{-}48}$ concentration ($\chi^2$ =13.41, *df* = 2, p=0.001; Table S15). Visualization of counterfactual predictions supports the influence of the Draft on increasing $bla_{OXA\text{-}48}$ concentration in wastewater as they consistently underestimated concentrations in comparison to observed data (Figure 5, main manuscript).

$bla_{KPC}$ may have contributed to the increase in the composite measure of carbapenemase concentration observed following the 2024 NFL Draft, although its contribution is less certain (Figure S7). Modelling indicates increases in normalized $bla_{KPC}$ concentrations following the Draft with marginal statistical significance (β=0.447, SE=0.212, p=0.035). The significance of this effect did not survive FDR correction (p=0.18). No significant gradual post event shift in concentration was observed (p=0.55), again indicating an abrupt shift in concentration rather than slow change. Flow was positively associated with $bla_{KPC}$ signal (p=0.019; Table S14) while precipitation was negatively associated with $bla_{KPC}$ signal but was not significant following FDR correction(p=0.060). Temperature showed positive association with $bla_{KPC}$ concentration that did not survive FDR correction (p=0.22). LRT suggests that the inclusion of the intervention parameters did not improve model fit ($\chi^2$ =3.53, *df* = 2, p=0.17), indicating that the NFL Draft may have had a minor influence on $bla_{KPC}$ signal (Table S15). Model diagnostics show no evidence of non-uniformity, dispersion or outlier influence (Table S16). Autocorrelation was adequately accounted for with the AR(1) structure of the GLMM.

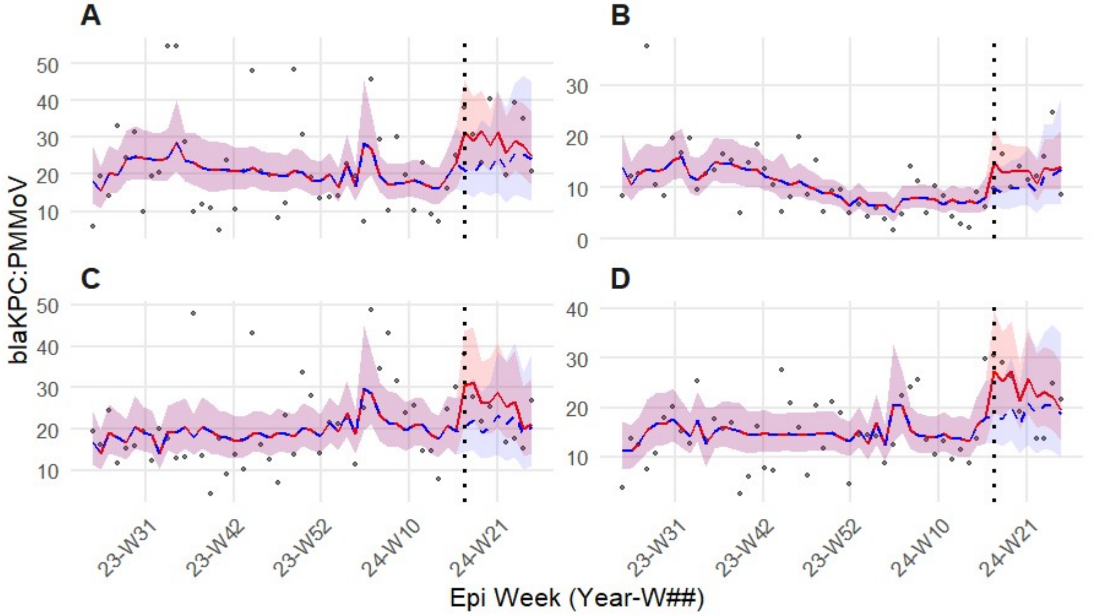


**Figure S7.** Longitudinal wastewater surveillance data for PMMoV normalized $bla_{KPC}$ concentrations at the four draft proximal sites (**A -JEFF** , **B-WEWTP, C-NIEA, D-OAK**) both before, and after the 2024 NFL draft (vertical dotted line). Points represent normalized wastewater concentrations. The redline and shading represent model predicted factual fit and 95% confidence intervals. The dashed blue line and shading represent the counter factual scenario (model predictions excluding draft related parameters).

Modelling for the remaining three carbapenemase genes ($bla_{NDM}$, $bla_{VIM}$, and $bla_{VIM7}$) indicates that the 2024 NFL Draft did not significantly impact normalized ARG concentration despite event estimates all indicating post-event increases (Figures S8-S10).

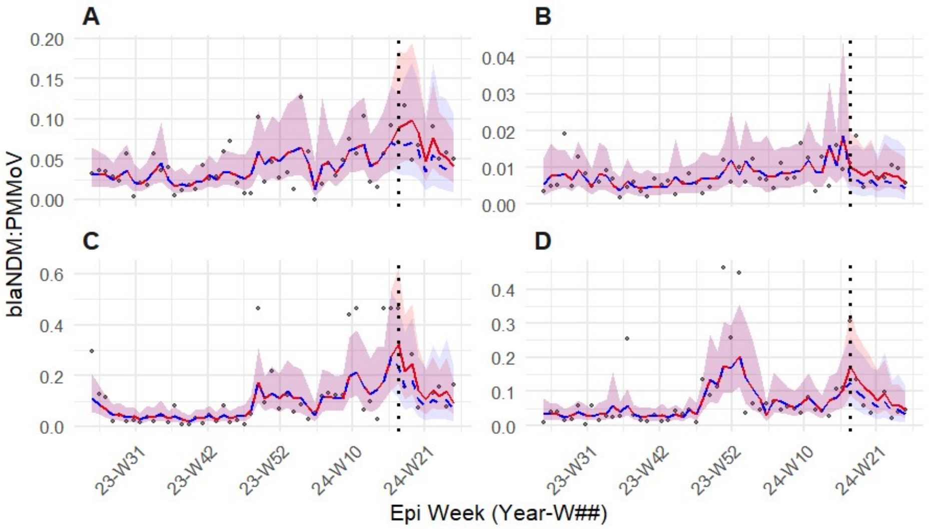


**Figure S8.** Longitudinal wastewater surveillance data for PMMoV normalized $bla_{NDM}$ concentrations at the four draft proximal sites (**A -JEFF** , **B-WEWTP, C-NIEA, D-OAK**) both before, and after the 2024 NFL draft (vertical dotted line). Points represent normalized wastewater concentrations. The redline and shading represent model predicted factual fit and 95% confidence intervals. The dashed blue line and shading represent the counter factual scenario (model predictions excluding draft related parameters).

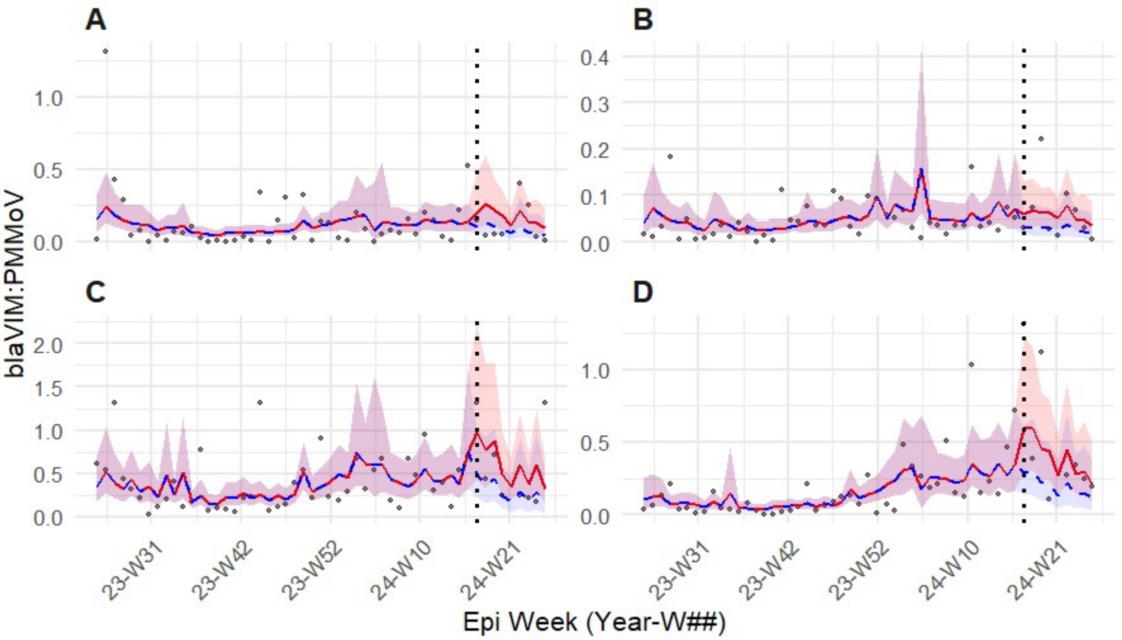


**Figure S9.** Longitudinal wastewater surveillance data for PMMoV normalized $\textit{bla}_{VIM}$ concentrations at the four draft proximal sites (**A -JEFF** , **B-WEWTP, C-NIEA, D-OAK**) both before, and after the 2024 NFL draft (vertical dotted line). Points represent normalized wastewater concentrations. The redline and shading represent model predicted factual fit and 95% confidence intervals. The dashed blue line and shading represent the counter factual scenario (model predictions excluding draft related parameters).

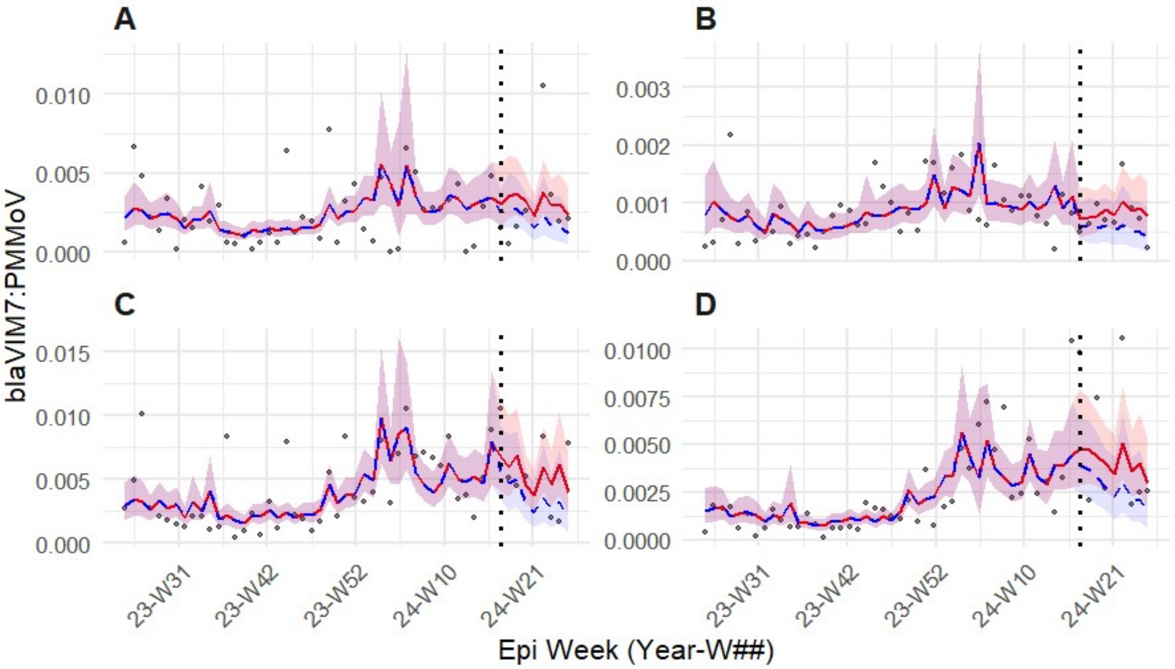


**Figure S10.** Longitudinal wastewater surveillance data for PMMoV normalized $\textit{bla}_{VIM7}$ concentrations at the four draft proximal sites (**A -JEFF** , **B-WEWTP, C-NIEA, D-OAK**) both before, and after the 2024 NFL draft (vertical dotted line). Points represent normalized wastewater concentrations. The redline and shading represent model predicted factual fit and 95% confidence intervals. The dashed blue line and shading represent the counter factual scenario (model predictions excluding draft related parameters).

The strongest predictor of normalized *$bla_{NDM}$* concentration was wastewater influent flow, which showed a significant positive non-linear influence on *$bla_{NDM}$* signal ($p<0.039$; Table S14). The strongest predictor of normalized *$bla_{VIM}$* concentration was wastewater temperature, but its effect did not remain significant after FDR correction ($p=0.099$). For *$bla_{VIM7}$* both temperature (p= 0.036) and flow (p= 0.036) were statistically significant predictors (p= 0.036) (Table S14). Temperature was negatively associated with ARG concentration while flow as positively associated with ARG concentration (Table S14). Overall, wastewater flow as the most consistently significant explanatory variable. It was a significant predictor of ARG concentration across 4 of 5 gene targets. Higher flows were consistently associated with increased normalized ARG concentrations. Temperature was only identified as a significant predictor of ARG for *$bla_{VIM7}$*, where it was found to be negatively associated with ARG concentration.

**Table S13.** Summary of GLMMs for ITS analysis of carbapenemase genes at affected sites using Gamma (and Tweedie) distribution, log link function, and an AR(1) correlation structure. Random intercept variance ($\sigma^2$) represents site level heterogeneity. The AR(1) variance and correlation describe within site temporal dependency of ARG concentrations.

| Carbapenemase gene/model | No. obs | AIC | BIC | logLik | Site Intercept (σ2) | AR(1) (σ2) | AR(1) ρ | Dispersion (σ2) |
|---|---|---|---|---|---|---|---|---|
| Composite carbapenemases/ Tweedie (log) | 209 | 125.1 | 181.9 | -45.6 | 0.0164 | 0.034 | 0.46 | 0.359 |
| $bla_{KPC}$/ Gamma(log) | 209 | 1462.3 | 1502.4 | -719.1 | <0.0001 | 0.046 | 0.94 | 0.217 |
| $bla_{OXA48}$/ Gamma(log) | 209 | 307.7 | 361.2 | -137.9 | 0.1570 | 0.222 | 0.19 | 0.367 |
| $bla_{NDM}$/ Gamma(log) | 209 | -954.2 | -894.1 | 495.1 | 0.0724 | 0.140 | 0.68 | 0.450 |
| $bla_{VIM}$/ Gamma(log) | 209 | -367.1 | -313.7 | 199.6 | <0.0001 | 0.349 | 0.99 | 1.020 |
| $bla_{VIM7}$/ Gamma(log) | 209 | -2180.6 | -2127.1 | 1106.3 | <0.0001 | 0.101 | 0.99 | 0.481 |

**Table S14.** Fixed effects coefficients for predictors remaining significant following Benjamini-Hochberg FDR correction ($p<0.05$) for GLMM ITS analysis of carbapenemase genes at affected sites. Estimates are reported on the link scale with 95% Confidence Intervals (CI). Standardized effects allow for comparison of magnitude across variables

| Model/Respiratory Virus | Predictor | Estimate | 95% CI | Std. Effect | FDR p-value |
|---|---|---|---|---|---|
| *$bla_{OXA-48}$* | Event | 1.378 | 0.646-2.110 | 0.805 | 0.0059 |
| *$bla_{OXA-48}$* | ns(Flow_s,3)1 | 2.358 | 1.311-3.405 | 1.377 | 0.0005 |
| *$bla_{KPC}$* | Flow_s | 0.269 | 0.108-0.431 | 0.026 | 0.0189 |
| *$bla_{NDM}$* | ns(Flow_s,3)1 | 1.5374 | 0.505-2.570 | 16.103 | 0.0356 |
| *$bla_{NDM}$* | ns(Flow_s,3)2 | 3.097 | 0.928-5.265 | 32.435 | 0.0389 |
| *$bla_{VIM7}$* | WWTemp_s | -0.376 | -0.633- -0.120 | -146.221 | 0.0356 |
| *$bla_{VIM7}$* | ns(Flow_s,3)1 | 1.330 | 0.461-2.199 | 516.710 | 0.0356 |

**Table S15.** Results of Likelihood Ratio Tests comparing full ITS GLMMs for carbapenemase genes (including "event" and "posttime" terms) with reduced models at the affected site cluster. A significant $\chi^2$ value indicates that the inclusion of event related variables significantly improved model fit, suggesting a detectable impact on ARG abundance.

| Carbapenemase gene/model | $\chi^2$ | *Degrees of freedom* | p-value | Interpretation |
|---|---|---|---|---|
| $bla_{OXA48}$ | 13.405 | 2 | 0.0012 | Significant |
| Composite carbapenemase | 5.604 | 2 | 0.0607 | Marginal |
| $bla_{KPC}$ | 3.527 | 2 | 0.1714 | Non-significant |
| $bla_{VIM}$ | 2.750 | 2 | 0.2529 | Non-significant |
| $bla_{VIM7}$ | 2.345 | 2 | 0.3096 | Non-significant |
| $bla_{NDM}$ | 0.925 | 2 | 0.6297 | Non-significant |

**Table S16.** Residual diagnostics for ITS GLMMs run on affected site carbapenemase data performed using simulated residuals (*DHARMa* package).

| Carbapenemase gene/model | KS | Dispersion | Quantile | Outliers (n) |
|---|---|---|---|---|
| Composite of carbapenemases* | 0.061 | 0.442 | 0.879 | <0.001 (4) |
| $bla_{KPC}$ | 0.910 | 0.952 | 0.240 | 1.000 (0) |
| $bla_{OXA-48}$ | 0.707 | 0.726 | 0.799 | 1.000 (0) |
| $bla_{NDM}$ | 0.112 | 0.266 | 0.543 | 0.342 (1) |
| $bla_{VIM}$ | 0.782 | 0.879 | 0.494 | <0.001 (2) |
| $bla_{VIM7}$ | 0.181 | 0.826 | 0.252 | 0.009(3) |
| *Tweedie distribution used instead of Gamma | | | | |

### Control sites

ITS specified GLMMs (Table S17) showed no evidence of a change in the level of the composite measure of ARG at the control sites following the 2024 NFL Draft (Figure S11). Neither the Draft intervention parameter (β=-0.039, SE=0.34, p=0.906) nor the post-event trend parameter (β=0.051, SE=0.05, p=0.334) were associated with changes to the composite measure of carbapenemase abundance. This finding was corroborated by LRT testing which indicated that inclusion of event related parameters did not improve model fit ($\chi^2$ =1.56, *df* = 2, p=0.457; Table S18). A positive association was observed between composite ARG signal and pH. However, this association did not remain significant following FDR correction.

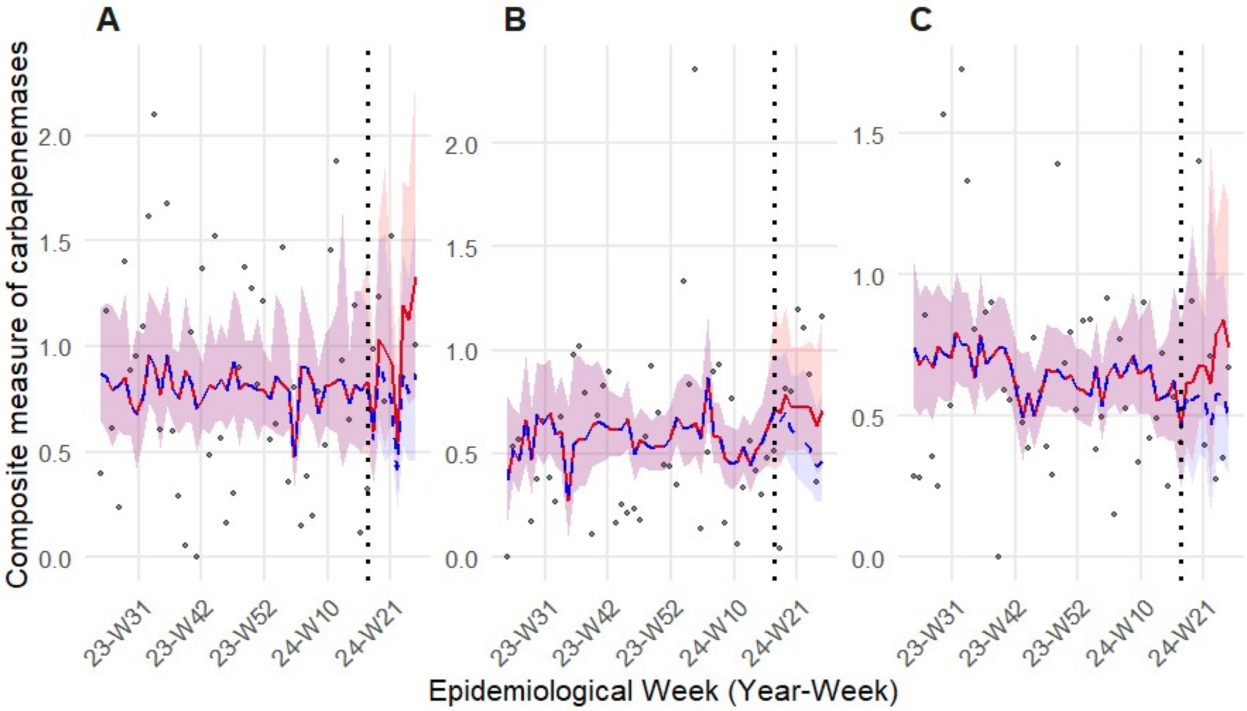


**Figure S11.** Longitudinal wastewater surveillance data for a composite measure of carbapenemase gene concentrations at the three control sites (**A -CHWW**, **B-LPCC, C-TB**) both before, and after the 2024 NFL draft (vertical dotted line). Points represent a composite carbapenemase index calculated by taking the mean of z-score standardized normalized gene concentrations for $bla_{KPC}$, $bla_{NDM}$, $bla_{OXA-48}$, $bla_{VIM}$, and $bla_{VIM7}$. The redline and shading represent model predicted factual fit and 95% confidence intervals. The dashed blue line and shading represent the counter factual scenario (model predictions excluding draft related parameters).

Individual GLMMs were run for carbapenemase genes for the control site cluster to ensure that creation of the composite had not masked a trend (Table S17). For $bla_{OXA-48}$ no statistically significant change could be detected in connection with the 2024 NFL Draft at the control sites (Figure S12). The Draft intervention parameter was non-significant (β=0.82, SE=0.45, p=0.278). The post-Draft change term was also non-significant (p=0.581). LRT testing also showed that the Draft did not influence the concentration of $bla_{OXA-48}$ at the control site ($\chi^2$ =3.49, *df* = 2, p=0.174; Table S18). Flow and precipitation exhibited marginal positive and negative associations respectively, but these were not significant after accounting for multiple testing.

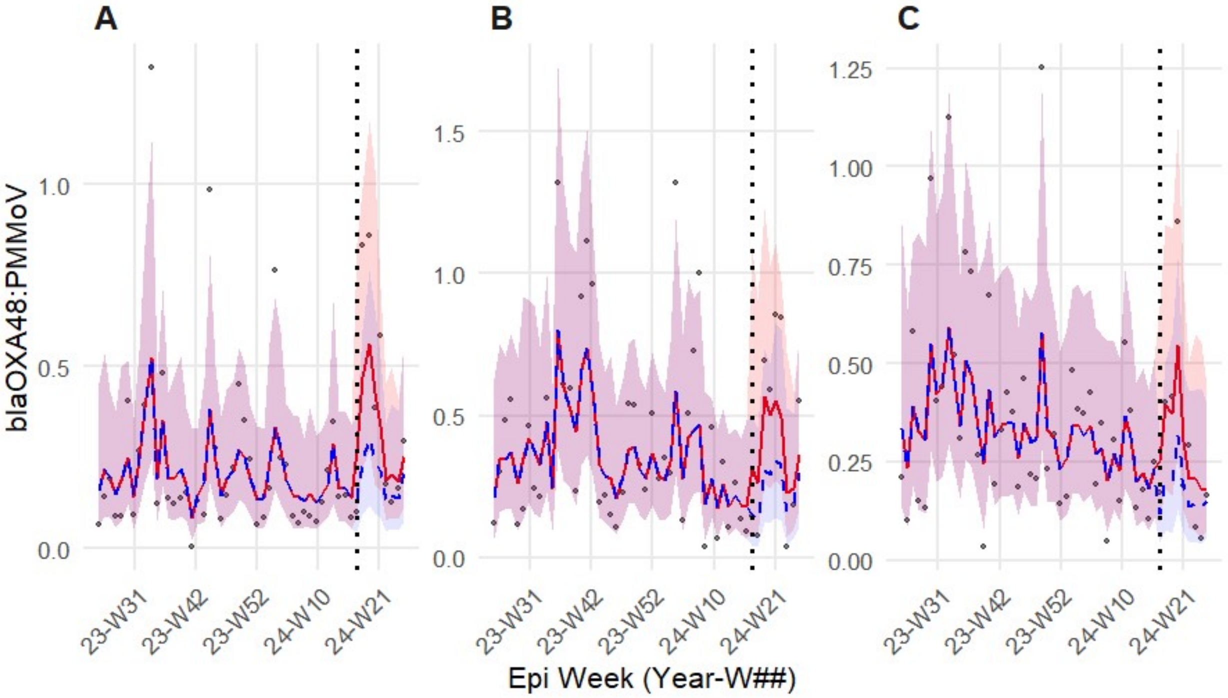


**Figure S12.** Longitudinal wastewater surveillance data for PMMoV normalized $bla_{OXA-48}$ concentrations at the three control sites (**A -CHWW**, **B-LPCC, C-TB**) both before, and after the 2024 NFL draft (vertical dotted line). Points represent normalized wastewater concentrations. The redline and shading represent model predicted factual fit and 95% confidence intervals. The dashed blue line and shading represent the counter factual scenario (model predictions excluding draft related parameters).

Similar results were found for $bla_{KPC}$ at the control sites with no evidence for event related changes in normalized gene concentrations (Figure S13). The stepwise intervention indicator was non-significant (β=0.17, SE=0.31, p=0.59) and the post-intervention time parameter showed no evidence of a gradual change in $bla_{KPC}$ concentration arising from the NFL Draft (p=0.51). Again, LRT corroborated this result with the exclusion of event related parameters not significantly improving model fit ($\chi^2$ =0.46, *df* = 2, p=0.80; Table S18). The strongest predictors of $bla_{KPC}$ concentration were wastewater flow, which showed a negative association, and wastewater temperature, which showed a positive association. However, these were not significant following FDR correction for multiple comparisons.

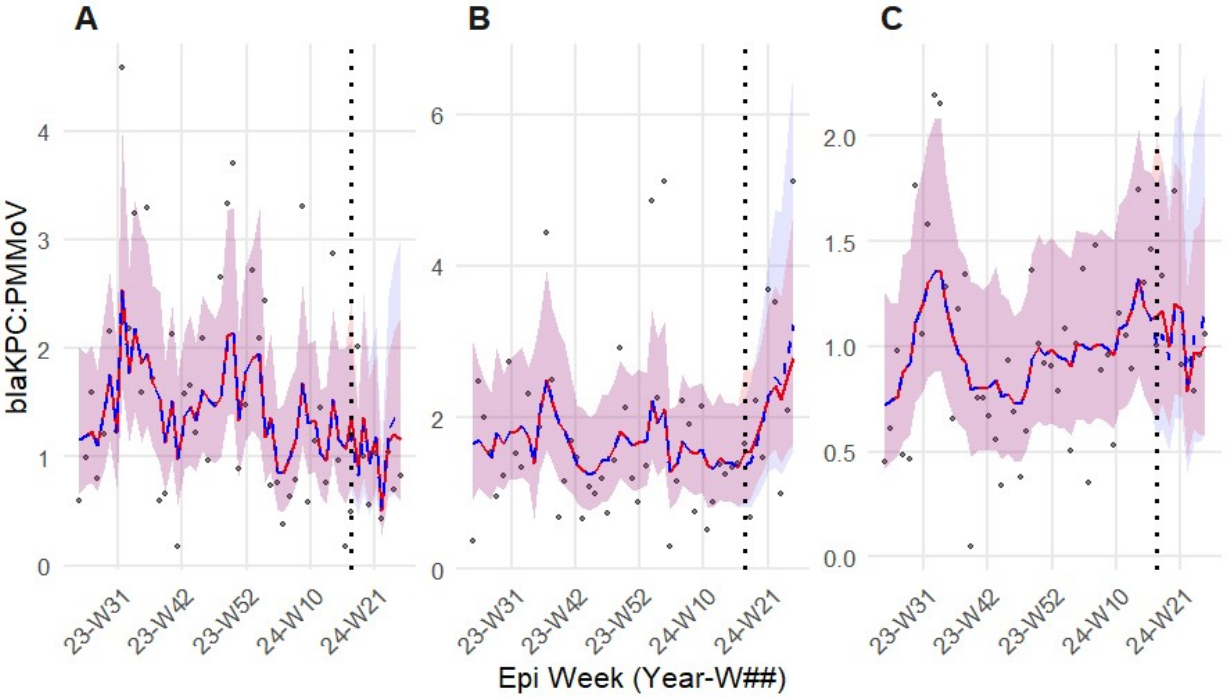


**Figure S13.** Longitudinal wastewater surveillance data for PMMoV normalized $bla_{KPC}$ concentrations at the three control sites (**A -CHWW**, **B-LPCC, C-TB**) both before, and after the 2024 NFL draft (vertical dotted line). Points represent normalized wastewater concentrations. The redline and shading represent model predicted factual fit and 95% confidence intervals. The dashed blue line and shading represent the counter factual scenario (model predictions excluding draft related parameters).

Analysis of the control site cluster indicates no immediate changes in normalized $bla_{NDM}$ concentrations associated with the 2024 NFL Draft (β=-0.34, SE=0.35, p=0.58; Figure S14). No gradual changes in $bla_{NDM}$ concentration were noted either (p=0.69). Model fit was not improved by the inclusion of the Draft related parameters ($\chi^2$ =0.92, *df* = 2, p=0.628; Table S18). Wastewater temperature and pH measurements explained part of the variability in $bla_{NDM}$ concentration, but these were not significant following FDR correction. For the control site cluster, both $bla_{VIM}$ *and* $bla_{VIM7}$ did not have sufficient detection frequency to allow modelling. *DHARMa* simulated residuals for the control site models showed no evidence of misspecification, indicating adequate model fit (Table S19).

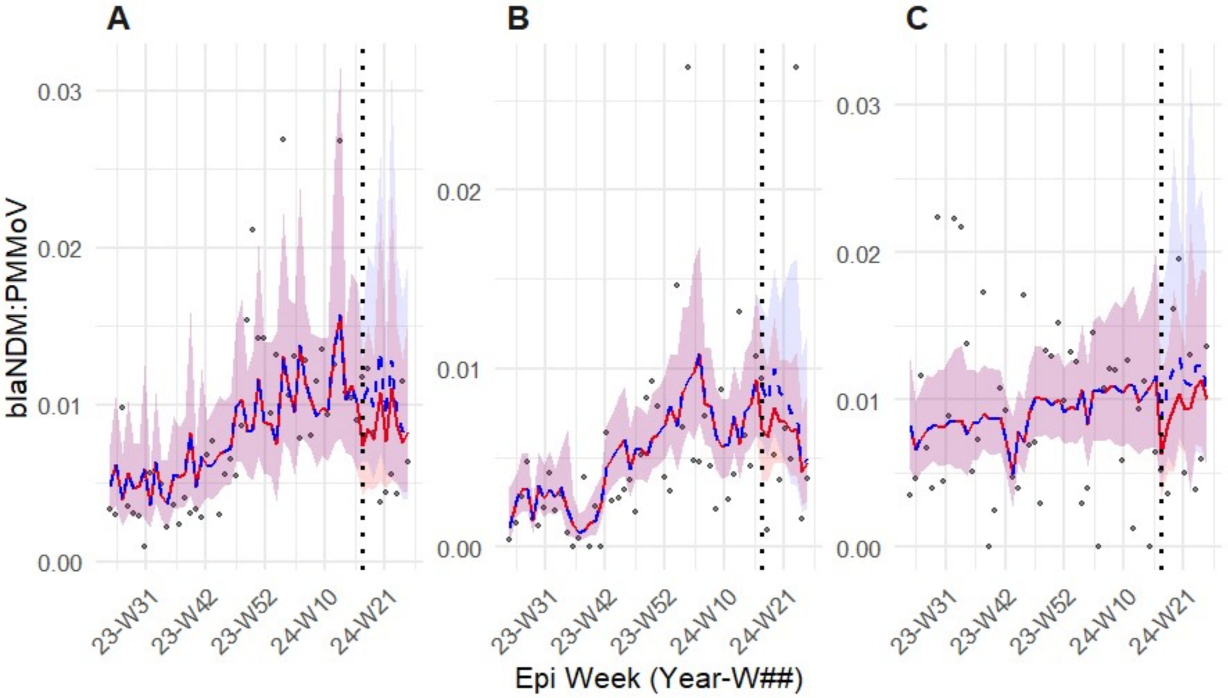


**Figure S14.** Longitudinal wastewater surveillance data for PMMoV normalized $bla_{NDM}$ concentrations at the three control sites (**A -CHWW**, **B-LPCC, C-TB**) both before, and after the 2024 NFL draft (vertical dotted line). Points represent normalized wastewater concentrations. The redline and shading represent model predicted factual fit and 95% confidence intervals. The dashed blue line and shading represent the counter factual scenario (model predictions excluding draft related parameters).

**Table S17.** Summary of GLMMs for ITS analysis of carbapenemase genes at the control sites using Gamma (and Tweedie) distribution, log link function, and an AR(1) correlation structure. Random intercept variance ($\sigma^2$) represents site level heterogeneity. The AR(1) variance and correlation describe within site temporal dependency of ARG concentrations.

| Carbapenemase gene/model | No. obs | AIC | BIC | logLik | Site Intercept ($\sigma^2$) | AR(1) ($\sigma^2$) | AR(1) $\rho$ | Dispersion ($\sigma^2$) |
|---|---|---|---|---|---|---|---|---|
| Composite carbapenemases/ Tweedie (log) | 159 | 150.4 | 196.5 | -60.2 | <0.0001 | NA | NA | 0.377 |
| $bla_{KPC}$/ Gamma(log) | 159 | 357.7 | 394.5 | -166.8 | <0.0001 | 0.0878 | 0.77 | 0.260 |
| $bla_{OXA-48}$/ Gamma(log) | 159 | -30.2 | 18.9 | 31.1 | 0.069 | 0.263 | 0.23 | 0.373 |
| $bla_{NDM}$/ Gamma(log) | 159 | -1286.1 | -1227.8 | 662.0 | <0.0001 | 0.019 | 0.82 | 0.10 |

**Table S18.** Results of Likelihood Ratio Tests comparing full ITS GLMMs for carbapenemase genes (including "event" and "posttime" terms) with reduced models at the control site cluster. A significant $\chi^2$ value indicates that the inclusion of event related variables significantly improved model fit, suggesting a detectable impact on ARG abundance

| Carbapenemase gene/model | $\chi^2$ | *Degrees of freedom* | p-value | Interpretation |
|---|---|---|---|---|
| Composite carbapenemase | 1.564 | 2 | 0.4575 | Non-significant |
| $bla_{OXA-48}$ | 3.493 | 2 | 0.1743 | Non-significant |
| $bla_{KPC}$ | 0.456 | 2 | 0.7962 | Non-significant |
| $bla_{NDM}$ | 0.930 | 2 | 0.6282 | Non-significant |

**Table S19.** Residual diagnostics for ITS GLMMs run on control site carbapenemase data performed using simulated residuals (DHARMa package).

| Carbapenemase gene/model | KS | Dispersion | Quantile | Outliers (n) |
|---|---|---|---|---|
| Composite of carbapenemases | 0.5563 | 0.292 | 0.2413 | 0.2724 (1) |
| $bla_{KPC}$ | 0.5085 | 0.762 | 0.4766 | 1 (0) |
| $bla_{OXA-48}$ | 0.9892 | 0.7956 | 0.243 | 1 (0) |
| $bla_{NDM}$ | 0.281 | 0.754 | 0.7053 | 0.0042 (3) |

#### S5.3. Multivariate resistome characterization

Normalized, scaled gene concentrations were visualized as a heatmap (Figure S15). No clear trends were observed following visual inspection. NMDS analysis revealed a 2-dimensional solution with good to excellent representation of data structure and little distortion (stress=0.068). Observations drawn from samples collected at the same site clustered closely regardless of temporal collection group (Figure 6, main manuscript).

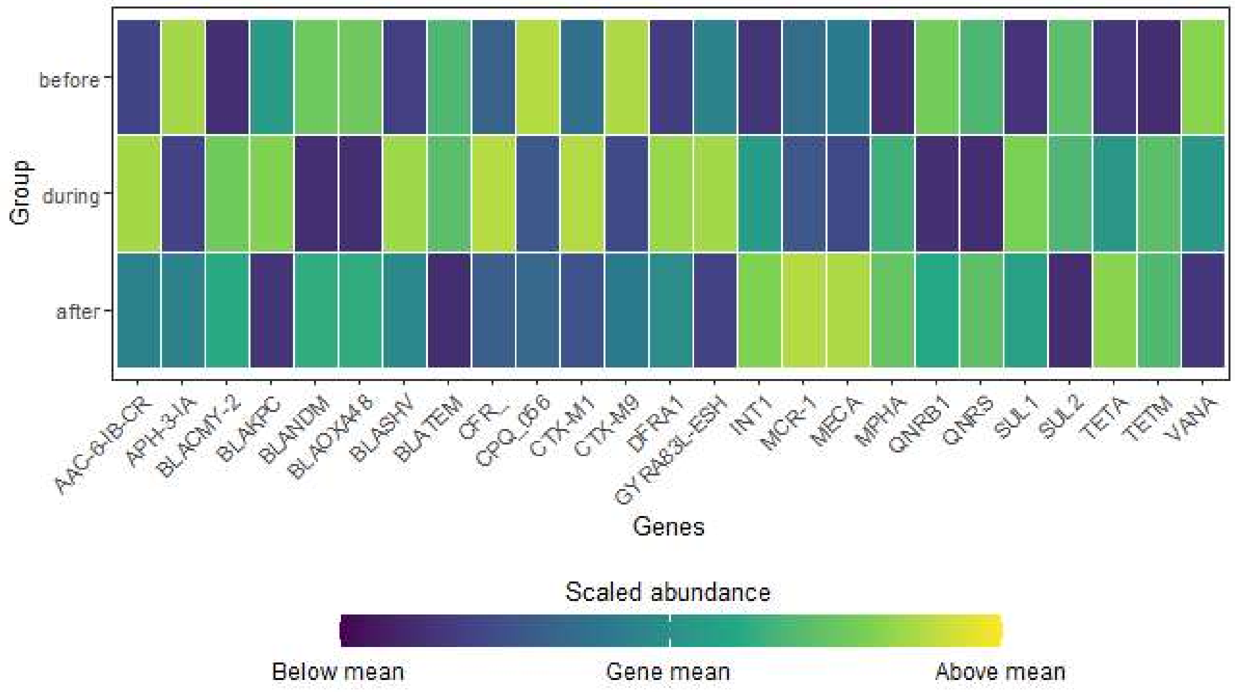


**Figure S15.** Mean relative abundance of antimicrobial resistance genes across the three collection periods. Data standardized via z-score. No distinct visual clustering or directional shifts in gene abundance were observed in relation to the NFL Draft event.

The PERMANOVA applied to test the effect of sampling period and site identity was significant ($F_{5,16}$=5.94, $R^2$ = 0.650, p = 0.001), indicating that site identity and sampling period explain approximately 65% of the variance in the data (Table S20). The marginal PERMANOVA run to discern the influence of site identity or sampling period explained a greater proportion of variance found that site identity drove almost all the variance in resistome structure ($R^2$ = 0.611, $F_{3,16}$= 9.30, p = 0.001; Figure S16). Sample collection period explained less variability and was statistically insignificant ($R^2$ = 0.033, $F_{2,16}$= 0.76, p = 0.531; Table 3, main manuscript).

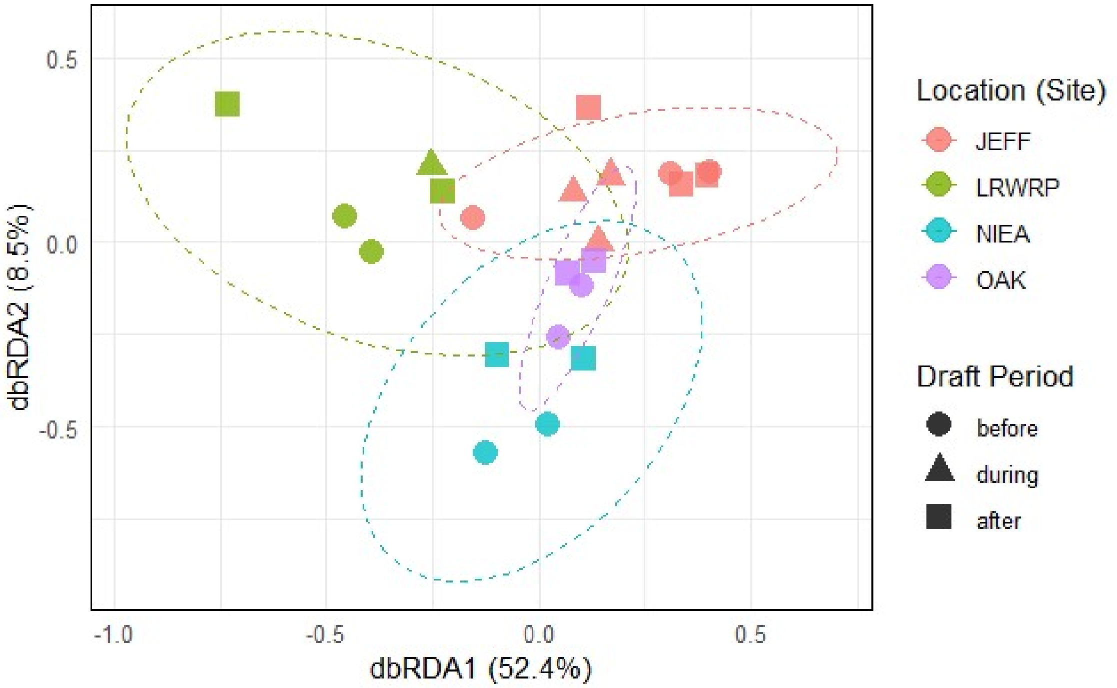


**Figure S16.** Distance-based Redundancy Analysis (dbRDA) illustrating the influence of geography (Site) versus time (Draft Period) on AMR profiles. Ellipses represent 95% confidence intervals for each site.

**Table S20.** Permutational multivariate analysis of variance (PERMANOVA) based on Bray-Curtis dissimilarities of fourth-root transformed AMR gene abundances normalized to crAssphage. The model evaluates the variance in resistome composition explained by sampling location and temporal period relative to the draft event (Before, During, and After). Results are based on 999 permutations.

| Df | SumOfSqs | $R^2$ | F | Pr(>F) |
|---|---|---|---|---|
| 5 | 0.048 | 0.650 | 5.939 | 0.001 |
| 16 | 0.026 | 0.350 | NA | NA |
| 21 | 0.074 | 1.000 | NA | NA |

To confirm that variability in the resistome was largely attributable to site identity, the marginal PERMANOVA corroborated via sensitivity analysis (re-running the PERMANOVA with the 16S rRNA gene). The sensitivity marginal PERMANOVA confirmed that site identity drove almost all the variance in resistome structure ($R^2$ = 0.547, $F_{3,16}$= 7.07, p = 0.001) while sample collection period remained statistically insignificant ($R^2$ = 0.036, $F_{2,16}$= 0.70, p = 0.621).

The PERMDISP test for homogeneity of dispersion showed that no significant difference in dispersion could be found for the sampling groups ($F_{2,19}$ = 0.90, p = 0.421) or site identity ($F_{3,18}$ = 1.55, p = 0.236; Figure S17). The first two principal components of the PCA explained a cumulative 77.96% of variance. PC1 was primarily driven by *int1*, *sul1*, *aac-6-ib-cr, teta, sul2, mpha, qnrs, dfra1,* $bla_{OXA\text{-}48}$, and $bla_{TEM}$, (highest absolute value loadings). PC1 loadings had the same sign (Table S21), suggesting that the top contributors varied together across sites with no gene specific trends. Additionally, PC1 was strongly driven by *int1* and *sul1*, suggesting sites differed by baseline anthropogenic pollution load, rather than by the occurrence of rare ARGs. The sensitivity analysis (re-running the PCA with the 16S rRNA gene) showed that with 16S rRNA included, the first two principal components accounted for 83.3% of variation with 16S rRNA contributing heavily to PC1 loadings.

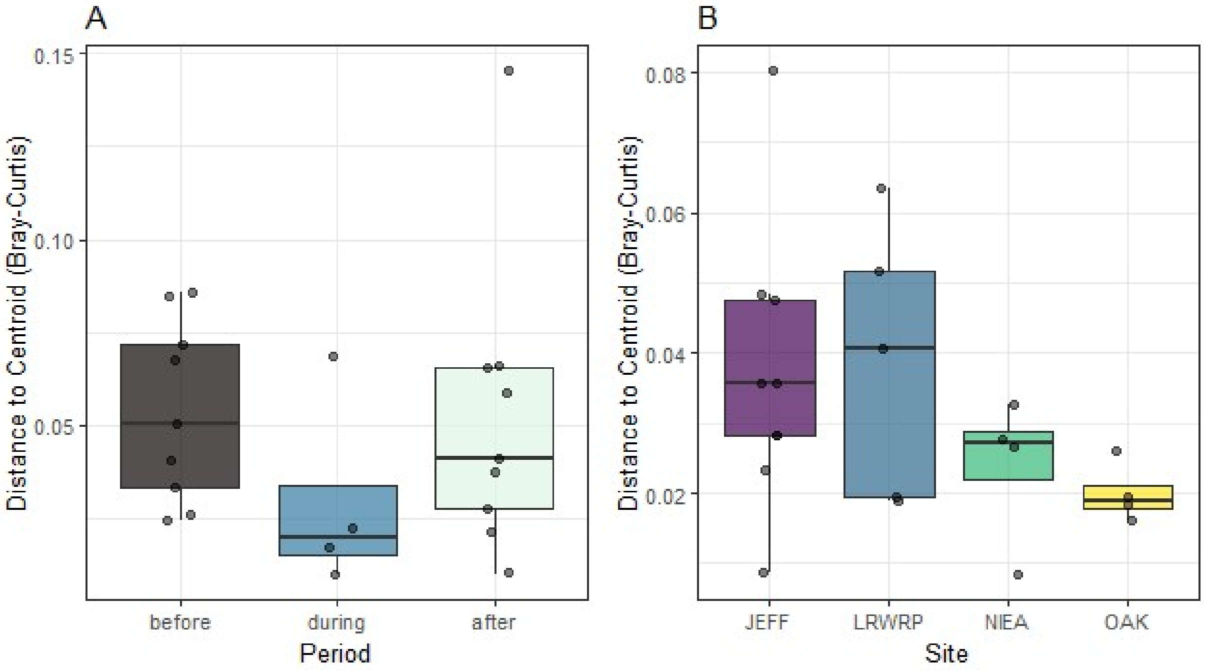


**Figure S17.** Boxplots representing the distance to group centroids for (A) Draft Period and (B) Site Location. Analysis of variance on the distances confirmed that the differences observed in resistome structure were not driven by heterogeneity of variance, supporting the validity of the PERMANOVA results.

**Table S21**. Detailed list of all 24 AMR gene targets and their corresponding loading scores for Principal Components 1 and 2. These values represent the contribution of each gene to the overall variance captured in the multivariate analysis.

| Gene | PC1 | PC2 |
|---|---|---|
| *aph(3')-Ia* | -0.139 | 0.028 |
| *aac(6')-Ib-cr* | -0.311 | -0.122 |
| $bla_{KPC}$ | -0.107 | 0.033 |
| $bla_{NDM}$ | -0.065 | 0.211 |
| $bla_{OXA-48}$ | -0.207 | 0.060 |
| $bla_{CMY-2}$ | -0.020 | -0.061 |
| $bla_{SHV}$ | -0.071 | -0.071 |
| $bla_{TEM}$ | -0.161 | 0.056 |
| $bla_{CTX-M1}$ | -0.101 | -0.023 |
| $bla_{CTX-M9}$ | -0.002 | 0.074 |
| *mcr-1* | 0.083 | 0.011 |
| *gyrA83L-Esh* | -0.016 | 0.487 |
| *QnrB1* | -0.036 | 0.108 |
| *qnrS* | -0.225 | 0.166 |
| *vanA* | -0.151 | -0.076 |
| *mphA* | -0.255 | 0.053 |
| *cfr* | 0.014 | 0.007 |
| *mecA* | -0.013 | -0.113 |
| *int1* | -0.489 | -0.133 |
| crAssphage | 0.000 | 0.000 |
| *sul1* | -0.441 | -0.121 |
| *sul2* | -0.259 | 0.030 |
| *tetA* | -0.289 | 0.105 |
| tetM | 0.087 | -0.746 |
| *dfrA1* | -0.212 | -0.137 |

A Shapiro-Wilk test for normality and Levene's test for homogeneity of variance were run for data collected at the JEFF interceptor on a per-target basis. Assumptions of normality and homogeneity of variance were not met for all gene targets across the "before", "during" and "after" groups and thus a Kruskal-Wallis test was run to test per gene differences between groups. Benjamini-Hochberg correction was used to adjust raw p-values to account for multiple testing. No significant difference was found between the concentration of any ARG in the "before", "during", or "after" groups (Table S22).

**Table S22.** Univariate analysis of individual AMR genes at the JEFF interceptor. Results of Kruskal-Wallis tests comparing log-normalized concentrations of specific ARGs across "before," "during," and "after" draft periods.

| ARG | No. Observations | H | df | p-value | p.adj | Significance |
|---|---|---|---|---|---|---|
| 16S rRNA | 18 | 1.626 | 2 | 0.444 | 0.820 | ns |
| *aac(6')-Ib-cr* | 18 | 0.924 | 2 | 0.630 | 0.863 | ns |
| *aph(3')-Ia* | 18 | 2.117 | 2 | 0.347 | 0.803 | ns |
| $bla_{CMY-2}$ | 18 | 5.158 | 2 | 0.076 | 0.636 | ns |
| $bla_{KPC}$ | 18 | 1.450 | 2 | 0.484 | 0.830 | ns |
| $bla_{NDM}$ | 18 | 3.895 | 2 | 0.143 | 0.740 | ns |
| $bla_{OXA-48}$ | 18 | 1.684 | 2 | 0.431 | 0.820 | ns |
| $bla_{SHV}$ | 18 | 0.573 | 2 | 0.751 | 0.863 | ns |
| $bla_{TEM}$ | 18 | 1.205 | 2 | 0.548 | 0.863 | ns |
| crAssphage | 18 | 0.000 | 2 | 1.000 | 1.000 | ns |
| $bla_{CTX-M1}$ | 18 | 0.292 | 2 | 0.864 | 0.922 | ns |
| $bla_{CTX-M9}$ | 18 | 2.020 | 2 | 0.364 | 0.803 | ns |
| *dfrA1* | 18 | 5.064 | 2 | 0.080 | 0.636 | ns |
| *gyrA83L-Esh* | 18 | 3.690 | 2 | 0.158 | 0.740 | ns |
| *int1* | 18 | 3.380 | 2 | 0.185 | 0.740 | ns |
| *mecA* | 18 | 0.246 | 2 | 0.884 | 0.922 | ns |
| *mphA* | 18 | 0.608 | 2 | 0.738 | 0.863 | ns |
| *QnrB1* | 18 | 5.298 | 2 | 0.071 | 0.636 | ns |
| *qnrS* | 18 | 2.000 | 2 | 0.368 | 0.803 | ns |
| *sul1* | 18 | 0.854 | 2 | 0.653 | 0.863 | ns |
| *sul2* | 18 | 0.713 | 2 | 0.700 | 0.863 | ns |
| *tetA* | 18 | 2.047 | 2 | 0.359 | 0.803 | ns |
| tet*M* | 18 | 0.561 | 2 | 0.755 | 0.863 | ns |
| *vanA* | 18 | 2.117 | 2 | 0.347 | 0.803 | ns |

### S5.4. Detection of carbapenemase genes in environmental samples collected in receiving waters surrounding the 2024 NFL Draft

Detection frequency rose following the Draft for $bla_{KPC}$, $bla_{OXA-48}$, and $bla_{VIM7}$. However, $bla_{VIM}$ was not detected in any samples and the detection frequency of $bla_{NDM}$ decreased across the Draft weekend (Table S23). Fisher's Exact test indicated a statistically significant rise in detection frequency for $bla_{KPC}$ ($p = 0.025$), but the significance did not survive adjustment for multiple comparisons. Detection frequency differences were non-significant for all other measured genes ($p>0.05$). Shapiro-Wilk testing showed evidence of non-normality of paired differences ($W=0.92$, $p=0.016$) and so a nonparametric paired Wilcoxon signed-rank test was used. Wilcoxon signed-rank tests for normalized $bla_{KPC}$ concentrations show significantly higher values for the post-Draft group ($V = 614$, $p<0.001$). Mean and median normalized $bla_{KPC}$ concentrations increased at all sites measured (Table S24). However, differences were not significant for all sites (Table S24). DS1, DS3 and DS4 all had statistically significant differences in median $bla_{KPC}$ concentration. Summary statistics indicate increases in $bla_{KPC}$ at PO and DS2, but these changes were not statistically significant. Finally, the UP site had minimal differences in normalized $bla_{KPC}$ concentrations with almost no change in median or mean concentration. Visualization of the log transformed PMMoV normalized $bla_{KPC}$ concentrations corroborate statistical findings with clear visual increases observable at all sites save UP (Figure 8, main manuscript).

**Table S23.** Detection frequencies of carbapenemase genes and the fecal indicator PMMoV in environmental samples collected before (n = 36) and after (n = 36) the draft/CSO event.

| ARG | Detection frequency before | Detection frequency after | p-value | Odds ratio | Fisher's Exact Test | p-value (FDR adj) |
|---|---|---|---|---|---|---|
| $bla_{KPC}$ | 0.833 (30/36) | 1.000(36/36) | 0.025 | Inf | Tested | 0.075 |
| $bla_{NDM}$ | 0.611 (22/36) | 0.444(16/36) | 0.238 | 0.514 | Tested | 0.267 |
| $bla_{OXA48}$ | 0.167 (6/36) | 0.306(11/36) | 0.267 | 2.176 | Tested | 0.267 |
| PMMoV | 0.972 (35/36) | 1.000(36/36) | 1.000 | Inf | Not tested | NA |
| $bla_{VIM}$ | 0.000 (0/36) | 0.000 (0/36) | NA | NA | Not tested | NA |
| $bla_{VIM7}$ | 0.000 (0/36) | 0.028(1/36) | 1.000 | Inf | Not tested | NA |

**Table S24.** Median log transformed PMMoV normalized $bla_{KPC}$ concentrations across six environmental sampling sites before and after the 2024 NFL Draft and associated combined sewer overflow (CSO) event.

| Site name | Median before | Median after | Mean difference | Wilcoxon p-value |
|---|---|---|---|---|
| DS1 | 1.030 | 1.631 | 0.522 | 0.031 |
| DS2 | 0.394 | 1.077 | 0.699 | 0.156 |
| DS3 | 0.286 | 1.787 | 1.429 | 0.031 |
| DS4 | 0.976 | 1.991 | 0.987 | 0.031 |
| PO | 1.023 | 1.233 | 0.238 | 0.094 |
| UP | 0.375 | 0.443 | 0.035 | 0.844 |

### S5.5. Measurement of ARGs in the Detroit River using an Open Array nanofluidic RT-qPCR card

Overall, detection frequency was higher in the "after" group with more ARGs being detected at a higher frequency. Detection frequency increased following the Draft weekend for 62.5% (15/24) of ARGs analyzed. Of the 15 targets with increased detection frequency following the Draft, 3 were not detected before the Draft, only being detected in samples collected following the Draft weekend. A further 33.3% of targets did not change detection frequence (8/24). Finally, $bla_{OXA\text{-}48}$ was only detected in the pre-Draft sample group, leading to decreased detection frequency (Figure 9, main manuscript).

The fitted GLMM had levels of variance for the random intercept, indicating large differences in baseline detection probabilities between sites ($\sigma^2$ = 62.07, SD=7.88). The variance of the random slope for the "after" group was large as well, suggesting that the effect of the 2024 NFL Draft on detection probability differed between sites ($\sigma^2$ =6.36, SD=2.52) with sites DS1 and DS3 having a larger relative increase (Table S25). Log-odds of detection are provided by the fixed effects of the GLMM with *crAssphage* concentrations in the before grouping acting as the baseline. Two genes, *tetA* and $bla_{CMY\text{-}2}$, were detected significantly less frequently than crAssphage in the before group (Table S26). Collection group had a significant effect on the probability of crAssphage detection (Estimate = 11.46, p = 0.0008), suggesting that the 2024 NFL Draft and CSO greatly increased detection probability for this fecal indicator. Interaction terms between gene and sampling group were not statistically significant, indicating that the relative magnitude of the post-Draft increase did not differ significantly among genes. However, post-hoc pairwise comparisons for each gene included in the model found that detection probability increased for 6 genes including: crAssphage*, AAC(6')-lb-cr, qnRS, tetA, mph(A),* and *bla*$_{KPC}$ (Table S27, Figure S18). *DHARMa* simulated residual diagnostics indicated that model fit was adequate (Table S28).

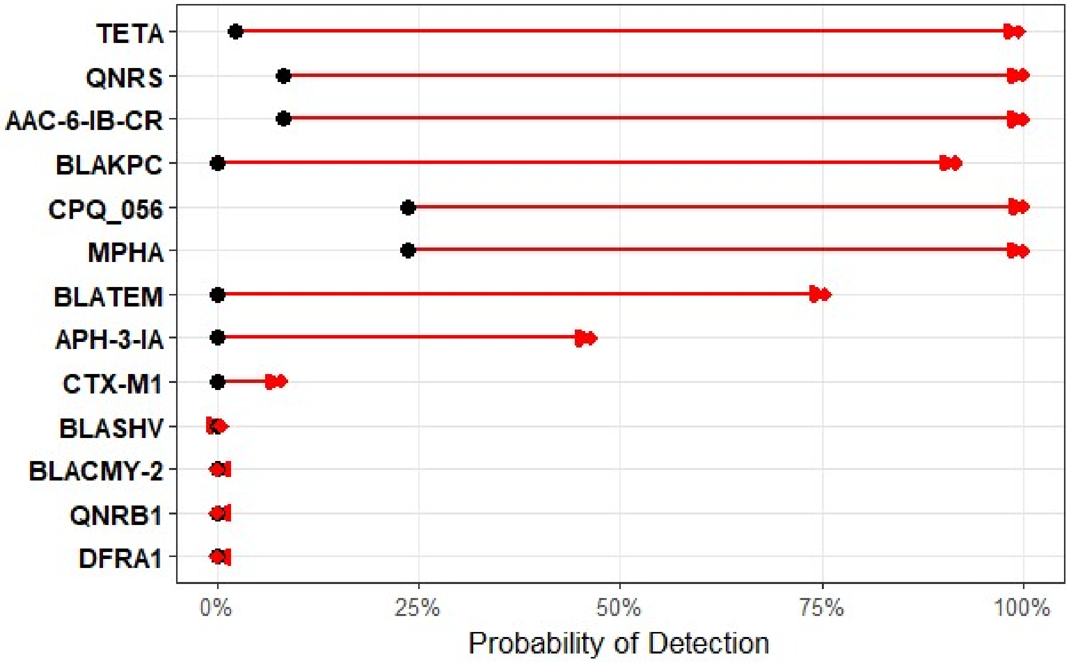


**Figure S18.** Predicted probability of detection before and after the CSO event. Estimated marginal means (EMMs) were derived from the GLMM with a binomial distribution and a logit link function. Black circles represent baseline detection probability, while red diamonds and arrows indicate the predicted probability following the CSO.

**Table S25**. Conditional modes of the random effects for the logistic regression model. The intercept represents site-specific deviations from the baseline detection probability, while the 'groupafter' column indicates random slopes, representing how the effect of the CSO varied across the six sampling locations.

| Site Name | (Intercept) | groupafter |
|---|---|---|
| DS1 | 3.226 | 1.939 |
| DS2 | -8.592 | -1.023 |
| DS3 | -2.267 | 1.804 |
| DS4 | 1.522 | -3.24 |
| PO | 14.291 | 0.784 |
| UP | -7.555 | -0.892 |

**Table S26.** Results of the logistic regression and post-hoc pairwise comparisons for ARG detection. The table displays the predicted probabilities of detection and 95% confidence intervals (CI) for each gene target before and after a CSO event.

| Predictor | log_odds | SE | z | p | OR | CI |
|---|---|---|---|---|---|---|
| (Intercept) | -1.166 | 3.689 | -0.316 | 0.752 | 0.312 | 0.000 – 429.825 |
| groupafter | 11.464 | 3.402 | 3.370 | 0.001 | 95,238.064 | 121.003 – 74958950.552 |
| targetAAC-6-IB-CR | -1.245 | 1.147 | -1.085 | 0.278 | 0.288 | 0.030 – 2.729 |
| targetAPH-3-IA | -7.598 | 5.267 | -1.443 | 0.149 | 0.001 | 0.000 – 15.258 |
| targetBLACMY-2 | -13.162 | 4.005 | -3.286 | 0.001 | 0.000 | 0.000 – 0.005 |
| targetBLAKPC | -7.598 | 5.267 | -1.443 | 0.149 | 0.001 | 0.000 – 15.257 |
| targetBLASHV | -7.598 | 5.267 | -1.443 | 0.149 | 0.001 | 0.000 – 15.258 |
| targetBLATEM | -7.598 | 5.267 | -1.443 | 0.149 | 0.001 | 0.000 – 15.258 |
| targetCTX-M1 | -7.598 | 5.267 | -1.443 | 0.149 | 0.001 | 0.000 – 15.258 |
| targetDFRA1 | -7.598 | 5.267 | -1.443 | 0.149 | 0.001 | 0.000 – 15.258 |
| targetMPHA | 0.000 | 1.161 | 0.000 | 1.000 | 1.000 | 0.103 – 9.727 |
| targetQNRB1 | -7.598 | 5.267 | -1.443 | 0.149 | 0.001 | 0.000 – 15.258 |
| targetQNRS | -1.245 | 1.147 | -1.085 | 0.278 | 0.288 | 0.030 – 2.729 |
| targetTETA | -2.570 | 1.258 | -2.043 | 0.041 | 0.077 | 0.006 – 0.901 |
| groupafter:targetAAC-6-IB-CR | -0.014 | 1.632 | -0.009 | 0.993 | 0.986 | 0.040 – 24.158 |
| groupafter:targetAPH-3-IA | -2.845 | 6.100 | -0.466 | 0.641 | 0.058 | 0.000 – 9047.259 |
| groupafter:targetBLACMY-2 | -12.235 | 7.703 | -1.588 | 0.112 | 0.000 | 0.000 – 17.533 |
| groupafter:targetBLAKPC | -0.314 | 6.059 | -0.052 | 0.959 | 0.730 | 0.000 – 105016.011 |
| groupafter:targetBLASHV | -7.934 | 6.723 | -1.180 | 0.238 | 0.000 | 0.000 – 189.234 |
| groupafter:targetBLATEM | -1.583 | 6.068 | -0.261 | 0.794 | 0.205 | 0.000 – 30002.360 |
| groupafter:targetCTX-M1 | -5.145 | 6.385 | -0.806 | 0.420 | 0.006 | 0.000 – 1586.519 |
| groupafter:targetDFRA1 | -12.814 | 8.122 | -1.578 | 0.115 | 0.000 | 0.000 – 22.313 |
| groupafter:targetMPHA | -2.511 | 1.705 | -1.473 | 0.141 | 0.081 | 0.003 – 2.295 |
| groupafter:targetQNRB1 | -12.814 | 8.122 | -1.578 | 0.115 | 0.000 | 0.000 – 22.313 |
| groupafter:targetQNRS | -0.014 | 1.632 | -0.009 | 0.993 | 0.986 | 0.040 – 24.158 |
| groupafter:targetTETA | -2.633 | 2.618 | -1.006 | 0.315 | 0.072 | 0.000 – 12.163 |
| Fit statistics: AIC:235.1, BIC: 363.7, Log-Likelihood:-88.5, Observations: 624<br>Random Effects: Observations are nested by collection site (n=6). Random intercept variance = 62.07 (SD=7.88). Random slope for groupafter variance =6.36 (SD=2.52); correlation 0.06 | | | | | | |

**Table S27.** Pairwise comparisons for ARG detection before and after a CSO event. Predicted probabilities (marginal means) and 95% confidence intervals (CI) for each gene target are displayed.

| Target | Prob (Before) | 95% CI (Before) | Prob (After) | 95% CI (After) | Log-Odds | SE | z | p-value | Odds Ratio |
|---|---|---|---|---|---|---|---|---|---|
| *crAssphage* | 0.238 | 0.00 – 1.00 | 1.000 | 0.87 – 1.00 | -11.46 | 3.40 | -3.37 | < 0.001 | 1.05 × 10^-5 |
| *aac(6')-Ib-cr* | 0.082 | 0.00 – 0.99 | 1.000 | 0.67 – 1.00 | -11.45 | 3.37 | -3.39 | < 0.001 | 1.07 × 10^-5 |
| *aph(3')-Ia* | 0.000 | 0.00 – 0.98 | 0.464 | 0.00 – 1.00 | -8.62 | 5.63 | -1.53 | 0.126 | 1.81 × 10^-4 |
| *bla*$_{CMY\text{-}2}$ | 0.000 | 0.00 – 0.02 | 0.000 | 0.00 – 0.03 | 0.77 | 6.69 | 0.12 | 0.908 | 2.161 |
| *bla*$_{KPC}$ | 0.000 | 0.00 – 0.98 | 0.916 | 0.01 – 1.00 | -11.15 | 5.64 | -1.98 | 0.048 | 1.44 × 10^-5 |
| *bla*$_{SHV}$ | 0.000 | 0.00 – 0.98 | 0.005 | 0.00 – 0.94 | -3.53 | 5.93 | -0.60 | 0.551 | 0.029 |
| *bla*$_{TEM}$ | 0.000 | 0.00 – 0.98 | 0.753 | 0.00 – 1.00 | -9.88 | 5.63 | -1.76 | 0.079 | 5.12 × 10^-5 |
| *bla*$_{CTX\text{-}M1}$ | 0.000 | 0.00 – 0.98 | 0.080 | 0.00 – 0.99 | -6.32 | 5.75 | -1.10 | 0.272 | 0.002 |
| *dfrA1* | 0.000 | 0.00 – 0.98 | 0.000 | 0.00 – 0.74 | 1.35 | 7.29 | 0.19 | 0.853 | 3.856 |
| *mphA* | 0.238 | 0.00 – 1.00 | 1.000 | 0.36 – 1.00 | -8.95 | 3.36 | -2.66 | 0.008 | 1.29 × 10^-4 |
| *QnrB1* | 0.000 | 0.00 – 0.98 | 0.000 | 0.00 – 0.74 | 1.35 | 7.29 | 0.19 | 0.853 | 3.856 |
| *qnrS* | 0.082 | 0.00 – 0.99 | 1.000 | 0.67 – 1.00 | -11.45 | 3.37 | -3.39 | < 0.001 | 1.07 × 10^-5 |
| *tetA* | 0.023 | 0.00 – 0.97 | 0.994 | 0.06 – 1.00 | -8.83 | 2.90 | -3.05 | 0.002 | 1.46 × 10^-4 |

**Table S28.** Residual diagnostics performed using simulated residuals in the DHARMa package for the logistic regression model using gene target as a fixed effect to determine if all genes reacted in the same way in response to the CSO event

| Test | Statistic | p-value | Interpretation |
|---|---|---|---|
| KS Test (Uniformity) | D=0.035 | 0.4269 | Residuals follow expected distribution |
| Dispersion Test | Ratio = 1.13 | 0.65 | No significant overdispersion detected |
| Outlier Test | Freq = 0 | 1 | No significant outliers detected |
| Quantile Test | N/A | 0.3108 | No significant deviations in quantiles |

The paired Wilcoxon signed-rank test conducted on logarithmically transformed normalized ARG concentrations corroborated detection frequency analyses. Generally, ARG concentrations increased following the 2024 NFL Draft weekend with ten ARGs having statistically significant increases in concentration and no ARGs showing a significant decrease in abundance (Table S29).

**Table S29.** Paired Wilcoxon Signed-Rank Test results for gene abundance across all environmental sampling sites for log normalized concentrations of ARGs before and after a CSO event.

| target | .y. | n1 | n2 | statistic | p | p.adj | significance | diff_median | diff_mean |
|---|---|---|---|---|---|---|---|---|---|
| *aac(6')-Ib-cr* | log_norm | 24 | 24 | 37 | 0.002 | 0.006 | ** | 1.548 | 0.825 |
| *aph(3')-Ia* | log_norm | 24 | 24 | 14 | 0.030 | 0.057 | ns | 1.221 | 0.377 |
| $bla_{CMY-2}$ | log_norm | 24 | 24 | 5 | 0.590 | 0.619 | ns | 0.000 | -0.020 |
| $bla_{KPC}$ | log_norm | 24 | 24 | 21 | 0.016 | 0.034 | * | 1.414 | 0.566 |
| $bla_{OXA-48}$ | log_norm | 24 | 24 | 13 | 0.675 | 0.675 | ns | 0.000 | -0.057 |
| $bla_{SHV}$ | log_norm | 24 | 24 | 14 | 0.055 | 0.088 | ns | 0.069 | 0.276 |
| $bla_{TEM}$ | log_norm | 24 | 24 | 28 | 0.074 | 0.110 | ns | 0.971 | 0.299 |
| crAssphage | log_norm | 24 | 24 | 10 | 0.000 | 0.000 | *** | 0.933 | 0.777 |
| $bla_{CTX-M1}$ | log_norm | 24 | 24 | 6 | 0.032 | 0.057 | ns | 0.000 | 0.317 |
| $bla_{CTX-M9}$ | log_norm | 24 | 24 | 0 | 0.181 | 0.224 | ns | 0.000 | 0.145 |
| *dfrA1* | log_norm | 24 | 24 | 10 | 0.083 | 0.116 | ns | 0.000 | 0.390 |
| *gyrA83L-Esh* | log_norm | 24 | 24 | 1 | 0.201 | 0.235 | ns | 0.000 | 0.351 |
| *int1* | log_norm | 24 | 24 | 10 | 0.000 | 0.000 | *** | 0.813 | 0.573 |
| *mecA* | log_norm | 24 | 24 | 0 | 0.181 | 0.224 | ns | 0.000 | 0.144 |
| *mphA* | log_norm | 24 | 24 | 8 | 0.000 | 0.001 | ** | 1.775 | 0.994 |
| *QnrB1* | log_norm | 24 | 24 | 8 | 0.353 | 0.390 | ns | 0.000 | 0.082 |
| *qnrS* | log_norm | 24 | 24 | 24 | 0.001 | 0.003 | ** | 1.344 | 1.004 |
| *sul1* | log_norm | 24 | 24 | 10 | 0.000 | 0.000 | *** | 1.155 | 0.859 |
| *sul2* | log_norm | 24 | 24 | 15 | 0.000 | 0.000 | *** | 1.260 | 1.187 |
| *tetA* | log_norm | 24 | 24 | 12 | 0.002 | 0.006 | ** | 2.025 | 0.736 |
| *tetM* | log_norm | 24 | 24 | 14 | 0.000 | 0.000 | *** | 1.021 | 1.435 |

The second GLMM run to more closely examine the impact of collection site on detection probability corroborated the results of the first GLMM by indicating a highly significant increase in detection probability for ARGs following the 2024 NFL Draft (Estimate = 5.82, SE = 0.79, z = 7.34, $p < 0.001$). DS1 was set as the reference site and modelling revealed that PO had a much higher detection probability than DS1 (Estimate =12.52, $p<0.001$) with all other sites having decreased detection probability in relation to DS1 (Table S30). *DHARMa* simulated residual diagnostics supported the adequacy of model fit (Table S31). Post-hoc pairwise comparisons of sites (including pooled data from before and after the 2024 NFL Draft) indicate that PO had the highest detection probability and DS2 the lowest. ARG detection probability at PO was significantly higher than all other sites, consistent with PO as a point source of ARG contamination (Table S32). To determine if the high concentration and saturated detection probability for ARGs at PO was masking temporal trends for the remaining sites, a sensitivity analysis (Wilcoxon signed-rank test excluding PO) was run. The results of the Wilcoxon signed-rank test show that constitutively high ARG concentration at PO was masking significant increases in ARG concentration at other sites. An additional 4 ARGs showed statistically significant increases in concentration when the PO site was removed from the Wilcoxon signed-rank test (Table S33).

**Table S31.** Residual diagnostics performed using simulated residuals in the DHARMa package for the logistic regression model using site as a fixed effect to determine if the CSO influenced the transect in a uniform or heterogenous manner.

| Diagnostic Test | Statistic | p-value | Result |
|---|---|---|---|
| Kolmogorov-Smirnov | D = 0.0478 | 0.1149 | Passed (Uniformity met) |
| Dispersion Test | Ratio = 0.980 | 0.966 | Passed (No over/under-dispersion) |
| Outlier Test | Freq = 0.0048 | 0.1306 | Passed (No significant outliers) |
| Quantile Test | - | 0.1316 | Passed (No significant deviations) |

**Table S32.** Pairwise comparisons of the likelihood of AMR gene detection between sampling sites. Estimates represent the log-odds ratios derived from the GLMM. Significant values ($p < 0.05$) indicate distinct spatial differences in the prevalence of resistance targets along the transect.

| Contrast | Estimate | SE | Odds Ratio | 95% CI | p (adj) |
|---|---|---|---|---|---|
| DS1 - DS2 | 8.179 | 1.11 | 3.57E+03 | 405 – 3.14e+04 | <0.0001 |
| DS1 - DS3 | 3.286 | 0.64 | 26.7 | 7.62 – 93.7 | <0.0001 |
| DS1 - DS4 | 2.675 | 0.599 | 14.5 | 4.48 – 46.9 | 0.0001 |
| DS1 - PO | -12.518 | 2.65 | 3.66E-06 | 2.03e-08 – 0.0006 | <0.0001 |
| DS1 - UP | 7.199 | 1.05 | 1.34E+03 | 171 – 1.05e+04 | <0.0001 |
| DS2 - DS3 | -4.893 | 0.922 | 0.0075 | 0.00123 – 0.0457 | <0.0001 |
| DS2 - DS4 | -5.504 | 0.946 | 0.00407 | 0.000637 – 0.026 | <0.0001 |
| DS2 - PO | -20.697 | 3.21 | 1.03E-09 | 1.92e-12 – 5.49e-07 | <0.0001 |
| DS2 - UP | -0.981 | 0.713 | 0.375 | 0.0927 – 1.52 | 0.7425 |
| DS3 - DS4 | -0.611 | 0.559 | 0.543 | 0.182 – 1.62 | 0.884 |
| DS3 - PO | -15.803 | 2.83 | 1.37E-07 | 5.33e-10 – 3.52e-05 | <0.0001 |
| DS3 - UP | 3.913 | 0.868 | 50 | 9.14 – 274 | <0.0001 |
| DS4 - PO | -15.192 | 2.798 | 2.52e-07 | 1.05e-09 – 6.07e-05 | <0.0001 |
| DS4 - UP | 4.524 | 0.891 | 92.2 | 16.1 – 528 | <0.0001 |
| PO - UP | 19.716 | 3.17 | 3.65E+08 | 7.31e+05 – 1.82e+11 | <0.0001 |

**Table S33.** Sensitivity analysis of gene abundance changes excluding the suspected point source (PO). Results reflect repeated paired Wilcoxon Signed-Rank Tests after removal of the PO site.

| target | .y. | n1 | n2 | statistic | p | p.adj | significance | diff_median | diff_mean |
|---|---|---|---|---|---|---|---|---|---|
| *aac(6')-Ib-cr* | log_norm | 20 | 20 | 15 | 0.001 | 0.003 | ** | 1.725 | 1.056 |
| *aph(3')-Ia* | log_norm | 20 | 20 | 0 | 0.009 | 0.015 | * | 0.000 | 0.564 |
| $bla_{CMY-2}$ | log_norm | 20 | 20 | 0 | 0.371 | 0.393 | ns | 0.000 | 0.031 |
| $bla_{KPC}$ | log_norm | 20 | 20 | 3 | 0.005 | 0.010 | ** | 1.350 | 0.726 |
| $bla_{OXA-48}$ | log_norm | 20 | 20 | 3 | 1.000 | 1.000 | ns | 0.000 | 0.053 |
| $bla_{SHV}$ | log_norm | 20 | 20 | 0 | 0.014 | 0.021 | * | 0.000 | 0.356 |
| $bla_{TEM}$ | log_norm | 20 | 20 | 13 | 0.083 | 0.100 | ns | 0.000 | 0.373 |
| crAssphage | log_norm | 20 | 20 | 1 | 0.000 | 0.001 | *** | 1.232 | 0.938 |
| $bla_{CTX-M1}$ | log_norm | 20 | 20 | 0 | 0.036 | 0.046 | * | 0.000 | 0.364 |
| $bla_{CTX-M9}$ | log_norm | 20 | 20 | 0 | NaN | NaN | ns | 0.000 | 0.000 |
| *dfrA1* | log_norm | 20 | 20 | 0 | 0.036 | 0.046 | * | 0.000 | 0.509 |
| *gyrA83L-Esh* | log_norm | 20 | 20 | 0 | NaN | NaN | ns | 0.000 | 0.000 |
| *int1* | log_norm | 20 | 20 | 0 | 0.000 | 0.000 | *** | 0.754 | 0.739 |
| *mecA* | log_norm | 20 | 20 | 0 | NaN | NaN | ns | 0.000 | 0.000 |
| *mphA* | log_norm | 20 | 20 | 0 | 0.000 | 0.001 | ** | 2.423 | 1.203 |
| *QnrB1* | log_norm | 20 | 20 | 0 | 0.181 | 0.204 | ns | 0.000 | 0.122 |
| *qnrS* | log_norm | 20 | 20 | 7 | 0.001 | 0.002 | ** | 2.487 | 1.257 |
| *sul1* | log_norm | 20 | 20 | 0 | 0.000 | 0.000 | *** | 1.354 | 1.060 |
| *sul2* | log_norm | 20 | 20 | 1 | 0.000 | 0.000 | *** | 1.107 | 1.455 |
| *tetA* | log_norm | 20 | 20 | 0 | 0.002 | 0.003 | ** | 1.822 | 0.904 |
| *tetM* | log_norm | 20 | 20 | 0 | 0.000 | 0.000 | *** | 2.014 | 1.760 |

#### S5.6 Comparison of nanofluidic qPCR card using TaqMan® assays in the OpenArray ® platform and RT-qPCR

Detection agreement differed between OpenArray ® and RT-qPCR methods for wastewater and river water matrices. For wastewater samples, with generally higher viral concentrations, both OpenArray ® and RT-qPCR demonstrated perfect detection for all targets tested (100% detection frequency). However, for lower viral concentration environmental samples detection frequency for the OpenArray ® was greatly reduced. RT-qPCR detected $bla_{KPC}$ in 100% of environmental samples (24/24) but the OpenArray ® detected $bla_{KPC}$ 54% of the time in the same sample set. OpenArray ® failed to detect $bla_{NDM}$ in any of the environmental samples while RT-qPCR exhibited a 67% detection frequency (13/24). Of the assessed gene targets, $bla_{OXA\text{-}48}$ displayed the greatest degree of concordance with 33% detection frequency for RT-qPCR and 29.2% detection frequency for OpenArray ®. Detection frequency was also calculated for the global data set (wastewater and environmental samples combined (Table 5, main manuscript).

Outlier detection using the 1.5x IQR criterion for each ARG target identified five pairings (3 $bla_{NDM}$ and 2 $bla_{KPC}$) which were removed from the data set. Subsequent quantitative analysis was carried out on a sample set with n =16 for $bla_{KPC}$, n=15 for $bla_{NDM}$, and n=18 for $bla_{OXA\text{-}48}$. Some data remained non-normally distributed following transformation and outlier removal and thus, non-parametric analysis was prioritized in the method comparison pipeline.

Bland Altman analysis found that $bla_{KPC}$ had the narrowest limits of agreement, while $bla_{NDM}$ and $bla_{OXA\text{-}48}$ had wider LoAs (Figure S19). Both $bla_{KPC}$ and $bla_{NDM}$ showed positive bias while $bla_{OXA\text{-}48}$ showed negative bias. Fold change analysis corroborated this finding by demonstrating that the OpenArray ® generally had a higher recovery rate in the high concentration wastewater matrix (Figure S20). Both $bla_{KPC}$ and $bla_{NDM}$ had median fold changes greater than 1. Again, the exception to this was $bla_{OXA\text{-}48}$ which had reduced recovery in the OpenArray ® when compared to RT-qPCR.

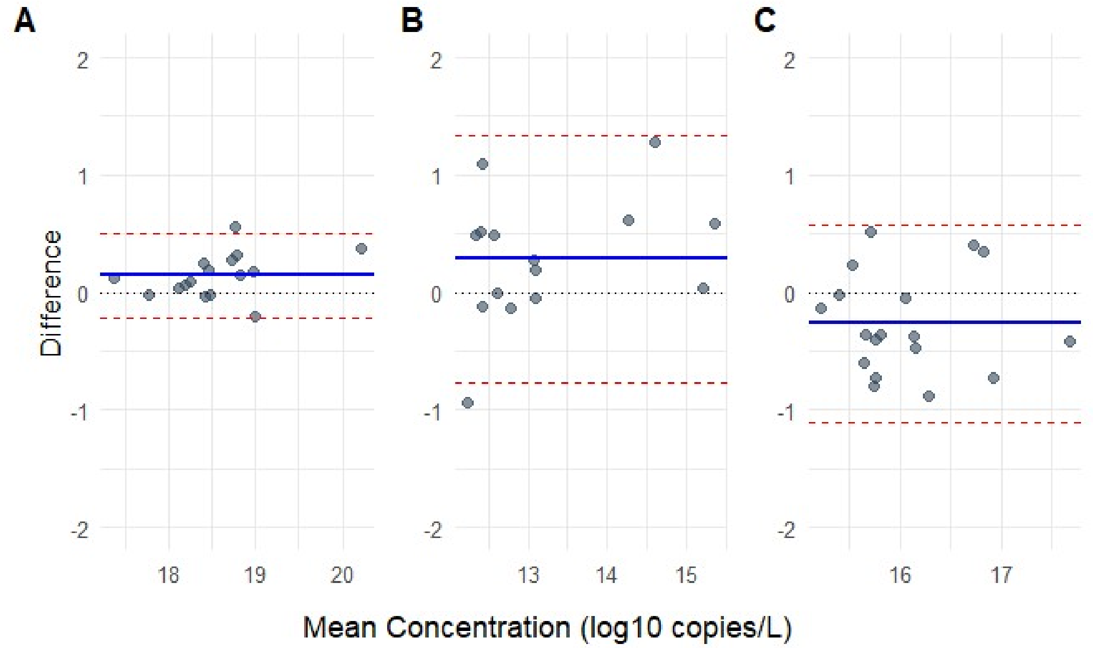


**Figure S19.** Bland-Altman plots visualzing method agreement for carbapenemase gene quantification between qRT-PCR and OpenArray® platforms. Plots represent agreement between measurement methods in log-transformed space for $bla_{KPC}$ (A), $bla_{NDM}$ (B), $bla_{OXA\text{-}48}$ (C). The solid blue line inidcates the average system diffence between quantification methods and the red dashed lines represent the range within which 95% of the differences fall.

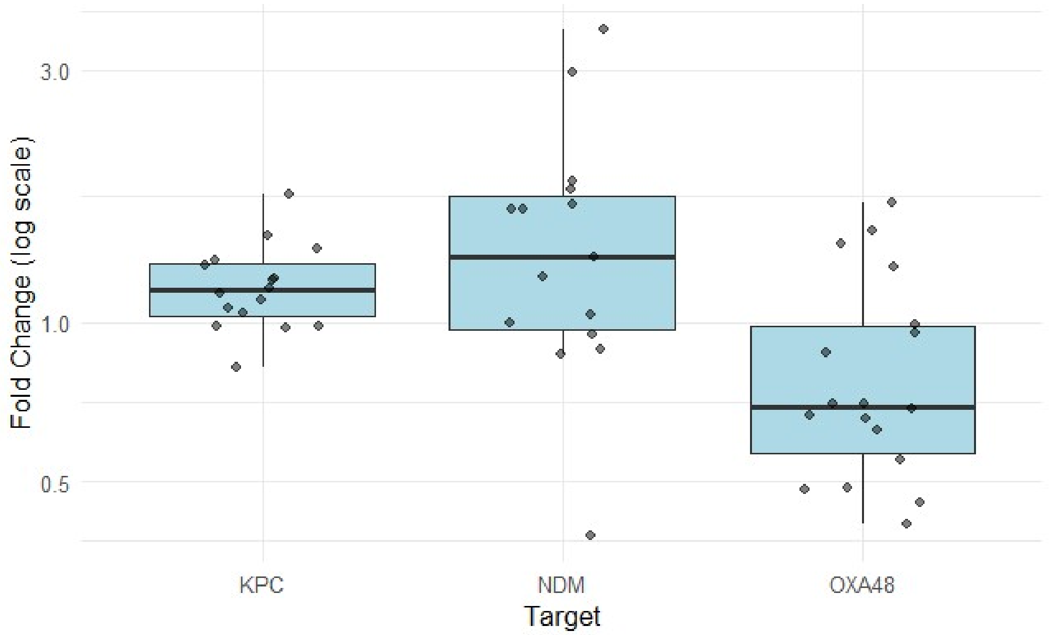


**Figure S20.** Fold change in carbapenemase gene concentrations between RT-qPCR and OpenArray® measurements. Box plots represent the ratio between concentration measurements for the OpenArray® and conventional RT-qPCR for $bla_{KPC}$, $bla_{NDM}$, and $bla_{OXA\text{-}48}$. Fold change values of 1 represent agreement between measurement techniques. Values greater than 1 indicate that the OpenArray® had a higher measured concentration than RT-qPCR.

Spearman's correlations between the results of the OpenArray ® analysis and the RT-qPCR analysis varied by target. Agreement was strongest for $bla_{KPC}$ and weakest for $bla_{OXA\text{-}48}$ (tablex). Linear relationships between the OpenArray ® and RT-qPCR quantification methods were generally strong with $bla_{KPC}$ having a Pearson's R of 0.97 and and $bla_{NDM}$ having a Pearson's R of 0.93 (Table 6, main manuscript).

Pasing-Pablok and Demming regression showed consistent proportional bias across targets (slopes >1.0), with large negative intercepts (Table S34, Figure S21). This pattern is consistent with relative under estimation of ARG concentration by the OpenArray ® at low concentrations, progressing to relative over estimation of ARG concentration by the OpenArray ® at higher analyte concentrations. Concordance correlation coefficients were highest for $bla_{KPC}$ (CCC=0.94) and lower for $bla_{NDM}$ (CCC = 0.88) and $bla_{OXA\text{-}48}$ (CCC=0.75) respectively (Table 6, main manuscript). These correspond to substantial methodological agreement across platforms for $bla_{KPC}$ and poor methodological agreement across platforms for both $bla_{NDM}$ and $bla_{OXA\text{-}48}$ based on previously proposed criteria [41].

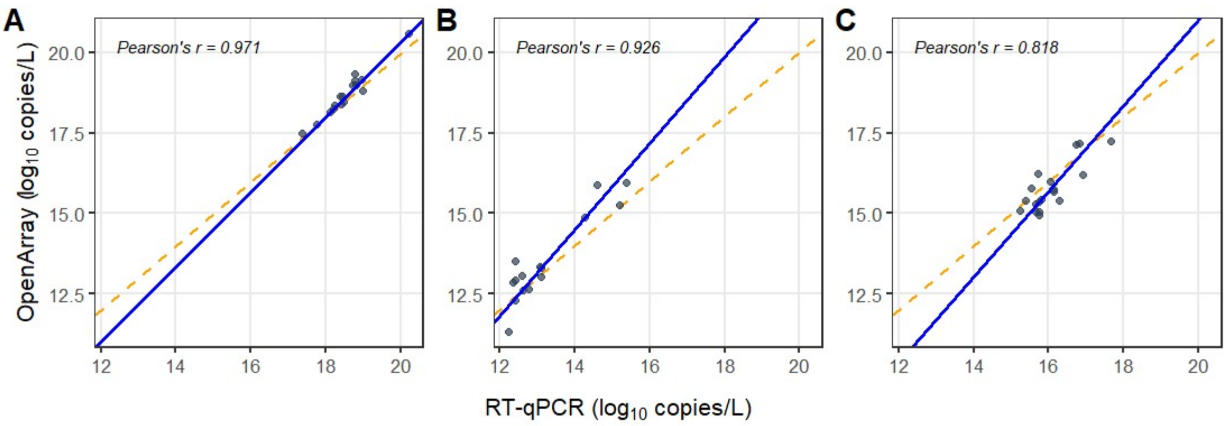


**Figure S21.** Passing-Bablok regression comparing OpenArray® and conventional qRT-PCR for quantification of carbapenemase genes;(A) $bla_{KPC}$, (B) $bla_{NDM}$, and (C) $bla_{OXA\text{-}48}$

**Table S34.** Method comparison regression statistics for the quantification of carbapenemase gene targets. Passing-Bablok and Deming regression analyses were performed for *blaKPC*, *blaNDM*, and *blaOXA48* to evaluate agreement between measurement methods. Intercept values (indicating constant bias) and Slope values (indicating proportional bias) are presented with their respective 95% confidence intervals (CI). Interpretation of bias is based on whether the 95% CI includes 0 for the intercept or 1 for the slope.

| Target | Method | Intercept (95% CI) | Slope (95% CI) | Interpretation |
|---|---|---|---|---|
| $bla_{KPC}$ | Passing-Bablok | -2.95 (-7.80, -0.32) | 1.17 (1.02, 1.43) | Proportional Bias |
| | Deming | -2.68 (-7.35, -0.19) | 1.15 (1.02, 1.41) | |
| $bla_{NDM}$ | Passing-Bablok | -4.31 (-12.90, 0.40) | 1.34 (0.99, 2.03) | Proportional Bias |
| | Deming | -3.51 (-9.41, 0.42) | 1.29 (1.00, 1.74) | |
| $bla_{OXA\text{-}48}$ | Passing-Bablok | -5.62 (-19.69, 0.45) | 1.33 (0.95, 2.21) | High systemic error |
| | Deming | -4.43 (-17.09, 1.29) | 1.26 (0.90, 2.06) | |

**S6. Supplemental note**

Comparison of the OpenArray ® and RT-qPCR quantification of three carbapenemase genes across two sample matrices was conducted to ascertain the validity of the OpenArray ® platform for ARG surveillance. We implemented a variety of methods to find that OpenArray ® can be used for ARG trend assessment in concentrated wastewater matrices but that it is not directly quantitatively interchangeable with RT-qPCR, likely due to low sample volumes.

Overall, detection agreement between the two assays was associated with ARG abundance in the sample measured. Concordance between methods was 100% in high concentration wastewater samples but was extremely poor (Cohen's Kappa 0 for $bla_{NDM}$ and $bla_{KPC}$) for environmental samples. Disagreement arose from non-detects by the OpenArray ® rather than disagreement between methods. This observation is consistent with the approximately 600-fold smaller reaction volume (33nL vs 20 µL) for the OpenArray ®, which lowers detection probability at low copy numbers. Thus, the OpenArray ® has low utility for environmental monitoring without upstream concentration [42–44].

Assessment of quantitative agreement between OpenArray ® in high concentration matrices also revealed significant discrepancies between methods. Passing-Bablok and Demming regression both revealed systematic proportional and constant bias between methods. Slopes greater than 1 and large negative intercepts are consistent with underestimation of samples at low concentrations and progressive over estimation at higher concentrations (consistent with more a less sensitive, due to nanolitre scale inputs, but more efficient assay). The magnitude of the negative intercept for $bla_{OXA\text{-}48}$ explains an apparent discrepancy between the Passing-Pablok/Demming slope being greater than 1 with a negative fold change and Bland-Altman mean bias. The negative intercept bias overwhelms the proportional bias, yielding the reduced recovery despite the greater than 1 slopes.

Agreement between the OpenArray ® and QRT-PCR platforms was target dependent with $bla_{KPC}$ showing the strongest quantitative agreement (CCC=0.94), indicating methodological interchangeability. However, for $bla_{NDM}$ and $bla_{OXA\text{-}48}$, overall agreement between methods was much lower (CCC=0.88 and CCC =0.75, respectively). For these targets OpenArray ® and RT-qPCR methods are not quantitatively interchangeable. However, strong correlations were found between OpenArray ® data and RT-qPCR data for all targets (Table 6, main manuscript). These correlations support reliable/reproducible trend tracking for OpenArray ® and RT-qPCR despite differences in the absolute concentration measured. Tracking of trends over time is an important aspect of both wastewater-based surveillance and of environmental monitoring and despite the apparent quantitative issues with the OpenArray ® platform, trend agreement is encouraging for the future development of platform. The multitude of ARG targets to be monitored and high relative abundance in wastewater matrices makes OpenArray ® a promising platform of ARG monitoring in high concentration matrices with potential future applications for environmental monitoring with the implementation of pre-amplification and further methodological confirmations.

1. Chatham-Kent Public Utilites Commission. *Water and Wastewater Master Plan (Approach 1) DRAFT*.
2. Government of Canada, S. C. Focus on Geography Series, 2021 Census - Leamington (Census subdivision). https://www12.statcan.gc.ca/census-recensement/2021/as-sa/fogs-spg/page.cfm?lang=E&topic=1&dguid=2021A00053537003 (2022).
3. Community Profile. https://www.leamington.ca/en/business/demographics.aspx (2023).
4. Government of Canada, S. C. Focus on Geography Series, 2021 Census - Thunder Bay (Census metropolitan area). https://www12.statcan.gc.ca/census-recensement/2021/as-sa/fogs-spg/page.cfm?lang=E&topic=1&dguid=2021S0503595 (2022).
5. Lu, X. *et al.* US CDC Real-Time Reverse Transcription PCR Panel for Detection of Severe Acute Respiratory Syndrome Coronavirus 2. *Emerg Infect Dis* **26**, (2020).
6. CDC. CDC's Influenza SARS-CoV-2 Multiplex Assay. *Centers for Disease Control and Prevention* https://www.cdc.gov/coronavirus/2019-ncov/lab/multiplex.html (2020).
7. Hughes, B. *et al. Respiratory Syncytial Virus (RSV) RNA in Wastewater Settled Solids Reflects RSV Clinical Positivity Rates*. http://medrxiv.org/lookup/doi/10.1101/2021.12.01.21267014 (2021) doi:10.1101/2021.12.01.21267014.
8. Klymus, K. E. *et al.* Reporting the limits of detection and quantification for environmental DNA assays. *Environmental DNA* **2**, 271–282 (2020).

18. BLAST: Basic Local Alignment Search Tool. https://blast.ncbi.nlm.nih.gov/Blast.cgi.

26. R Core Team. *R: A Language and Environment for Statistical Computing*. (R Foundation for Statistical Computing, Vienna, Austria, 2025).

32. CSO/SSO List - MiEnviro Portal. https://mienviro.michigan.gov/ncore/external/overflow/list.

33. Signorell, A. *DescTools: Tools for Descriptive Statistics*. (2025). doi:10.32614/CRAN.package.DescTools.

34. Potapov, S. *et al. Mcr: Method Comparison Regression*. (2024). doi:10.32614/CRAN.package.mcr.

35. A. C. Davison & D. V. Hinkley. *Bootstrap Methods and Their Applications*. (Cambridge University Press, Cambridge, 1997).
